# Multiclass risk models for ovarian malignancy: an illustration of prediction uncertainty due to the choice of algorithm

**DOI:** 10.1101/2023.07.25.23293141

**Authors:** Ledger Ashleigh, Ceusters Jolien, Valentin Lil, Testa Antonia, Caroline VAN Holsbeke, Franchi Dorella, Bourne Tom, Froyman Wouter, Timmerman Dirk, Ben VAN Calster

## Abstract

**OBJECTIVE:** To compare performance and probability estimates of six algorithms to estimate the probabilities that an ovarian tumor is benign, borderline malignant, stage I primary invasive, stage II-IV primary invasive, or secondary metastatic.

**MATERIALS AND METHODS:** Models were developed on 5909 patients (recruited 1999-2012) and validated on 3199 patients (2012-2015). Nine clinical and ultrasound predictors were used. Outcome was based on histology following surgery within 120 days after the ultrasound examination. We developed models using multinomial logistic regression (MLR), Ridge MLR, random forest (RF), XGBoost, neural networks (NN), and support vector machines (SVM).

**RESULTS:** Benign tumors were most common (62%), secondary metastatic tumors least common (5%). XGBoost, RF, NN and MLR had similar performance: c-statistics for benign versus any type of malignant tumors were 0.92, multiclass c-statistics 0.54-0.55, average Estimated Calibration Indexes 0.03-0.07, and Net Benefits at the 10% malignancy risk threshold 0.33-0.34. Despite poorer discrimination and calibration performance for Ridge MLR and in particular SVM, Net Benefits were similar for all models. The estimated probabilities often differed strongly between models. For example, the probability of a benign tumor differed by more than 20 percentage points in 29% of the patients, and by more than 30 percentage points in 16% of the patients.

**DISCUSSION:** Several regression and machine learning models had very good and similar performance in terms of discrimination, calibration and clinical utility. Nevertheless, individual probabilities often varied substantially.

**CONCLUSION:** Machine learning did not outperform MLR. The choice of algorithm can strongly affect probabilities given to a patient.

## BACKGROUND AND SIGNIFICANCE

Patients with an ovarian tumor should be managed appropriately. There is evidence that treatment in oncology centers improves ovarian cancer prognosis.[1, 2] However, benign ovarian cysts are frequent and can be managed conservatively (i.e. non-surgically with clinical and ultrasound follow-up) or with surgery in a general hospital.[3] Risk prediction models can support optimal patient triage by estimating a patient’s risk of malignancy based on a set of predictors.[4, 5] ADNEX is a multinomial logistic regression (MLR) model that uses nine clinical and ultrasound predictors to estimate the probabilities that a tumor is benign, borderline, stage I primary invasive, stage II-IV primary invasive, or secondary metastatic.[6–8] ADNEX differentiates between four types of malignancies because these tumor types require different management.[7, 9]

There is an increasing interest in the use of flexible machine learning algorithms to develop prediction models.[10–12] Contrary to regression models, flexible machine learning algorithms do not require the user to specify the model structure: these algorithms automatically search for nonlinear associations and potential interactions between predictors.[10] This may result in better performing models, but poor design and methodology may yield misleading and overfitted results.[10, 11] A recent systematic review observed better performance for flexible machine learning algorithms versus logistic regression when comparisons were at high risk of bias, but not when comparisons were at low risk of bias.[10] Few of the included studies addressed the accuracy of the risk estimates (calibration), none assessed clinical utility.

In addition, there is increased awareness for uncertainty of predictions.[13, 14] It is known that probability estimates for individuals are unstable, in the sense that a fitting the model on different sample from the same population may lead to very different probability estimates for individual patients.[15, 16] This instability decreases with the sample size for model development, but is considerable even when models are based on currently recommended sample sizes.[16, 17] Apart from instability, ‘model uncertainty’ reflects the impact of various decisions made during model development on the estimated probabilities for individual patients. Modeling decision may relate to issues such as the choice of predictors or the method to handle missing data.[18, 19] All other modeling decision being equal, the choice of modeling algorithm (e.g. logistic regression versus random forest) may also play a role.

## OBJECTIVE

In this study, we (1) compare the performance of multiclass risk models for ovarian cancer diagnosis based on regression and flexible machine learning algorithms in terms of discrimination, calibration, and clinical utility, and (2) assess differences between the models regarding the estimated probabilities for individual patients to study model uncertainty caused by choosing a particular algorithm.

## MATERIALS AND METHODS

### Study design, setting and participants

This is a secondary analysis of prospectively collected data from multicenter cohort studies that were conducted by the International Ovarian Tumor Analysis (IOTA) group. For model training, we used data from 5909 consecutively recruited patients at 24 centers across four consecutive cohort studies between 1999 and 2012.[6, 20-23] All patients had at least one adnexal (ovarian, para-ovarian, or tubal) mass that was judged not to be a physiological cyst, provided consent for transvaginal ultrasound examination, were not pregnant, and underwent surgical removal of the adnexal mass within 120 days after the ultrasound examination. This dataset was also used to develop the ADNEX model.[6] For external validation, we used data from 3199 consecutively recruited patients at 25 centers between 2012 and 2015.[8] All patients had at least one adnexal mass that was judged not to be a physiological cyst with a largest diameter below 3 cm and provided consent for transvaginal ultrasound examination. Although this study recruited patients that subsequently underwent surgery or were managed conservatively, the current work only used data from patients that were operated within 120 days after the ultrasound examination without additional preoperative ultrasound visits. The external validation dataset was therefore comparable to the training dataset.

Participating centers were ultrasound units in a gynecological oncology center (labeled oncology centers), or gynecological ultrasound units not linked to an oncology center. All mother studies received ethics approval from the Research Ethics Committee of the University Hospitals Leuven and from each local ethics committee. All participants provided informed consent. We obtained approval from the Ethics Committee in Leuven (S64709) for secondary use of the data for methodological purposes. We report this study using the TRIPOD checklist.[4, 24]

### Data collection

A standardized history from each patient was taken at the inclusion visit to obtain clinical information, and all patients underwent a standardized transvaginal ultrasound examination.[25] Transabdominal sonography was added if necessary, e.g. for large masses. Information on a set of predefined gray scale and color or power Doppler ultrasound variables was collected following the research protocol. When more than one mass was present, examiners included the mass with the most complex ultrasound morphology. If multiple masses with similar morphology were found, the largest mass or the mass best seen on ultrasound was included. Measurement of CA125 was neither mandatory nor standardized but was done according to local protocols regarding kits and timing.

### Outcome

The outcome was the classification of the mass into one of five outcome categories based on the histological diagnosis of the mass following laparotomy or laparoscopic surgery and on staging of malignant tumors using the classification of the International Federation of Gynecology and Obstetrics (FIGO): benign, borderline, stage I primary invasive, stage II-IV primary invasive, or, secondary metastasis.[26, 27] The histological assessment was performed without knowing the detailed results of the ultrasound examination, but pathologists might have received clinically relevant information as per local procedures.

### Statistical analysis

#### Predictors and sample size

We used the following nine clinical and ultrasound predictors: type of center (oncology center vs other), patient age (years), serum CA125 level (U/ml), proportion of solid tissue (maximum diameter of the largest solid component divided by the maximum diameter of the lesion), maximum diameter of the lesion (mm), presence of shadows (yes/no), presence of ascites (yes/no), presence of more than ten cyst locules (yes/no), and number of papillary projections (0, 1, 2, 3, >3). These predictors were selected for the ADNEX model based on expert domain knowledge regarding likely diagnostic importance, objectivity, and measurement difficulty, and based on stability between centers (see Supplementary Material 1 for more information).[6] We also developed models without CA125: not all centers routinely measure CA125, and including CA125 implies that predictions can only be made when the laboratory result becomes available. We discuss the adequacy of our study sample size in Supplementary Material 2.

#### Algorithms

We developed models using standard MLR, ridge MLR, random forest (RF), extreme gradient boosting (XGBoost), neural networks (NN), and support vector machines (SVM).[28–31] For the MLR models, continuous variables were modeled with restricted cubic splines (using 3 knots) to allow for nonlinear associations.[32] The hyperparameters were tuned with 10-fold cross-validation on the development data (Supplementary Material 3). Using the selected hyperparameters, the full development data was used to train the model.

#### Model performance on external validation data

Discrimination was assessed with the Polytomous Discrimination Index (PDI), a multiclass extension of the binary c-statistic (or area under the receiver operating characteristic curve, AUROC).[33] In this study, PDI equals 0.2 (one divided by five outcome categories) for useless models, and 1 for perfect discrimination: PDI estimates the probability that the model can correctly identify a patient from a randomly chosen category from a set of five patients (one from each outcome category). We also calculated pairwise c-statistics for each pair of outcome categories using the conditional risk method.[34] Finally, we calculated the binary c-statistic to discriminate benign from any type of malignant tumor. The estimated risk of any type of malignancy equals one minus the estimated probability of a benign tumor. PDI and c-statistics were analyzed through meta-analysis of center-specific results. We calculated 95% prediction intervals (PI) from the meta-analysis to indicate what performance to expect in a new center.

Calibration was assessed using flexible (loess-based) calibration curves per outcome; center-specific curves were averaged and weighted by the square root of sample size.[35] Calibration curves were summarized by the rescaled Estimated Calibration Index (ECI).[36] The rescaled ECI equals 0 if the calibration curve fully coincides with the diagonal line, and 1 if the calibration curve is horizontal (i.e. the model has no predictive ability).

We calculated the Net Benefit to assess the utility of the model to select patients for referral to a gynecologic oncology center.[37, 38] A consensus statement suggests to refer patients when the risk of malignancy is ≥10%.[9] We plotted Net Benefit for malignancy risk thresholds between 5% and 40% in a decision curve, but we focus on the 10% risk threshold. At each threshold, Net Benefit of the models is compared with default strategies: select everyone (‘treat all’) or select no-one (‘treat none’) for referral.[37, 38] Net Benefit was calculated using meta-analysis of center-specific results.[39] We calculated decision reversal for each pair of models by calculating the percentage of patients for which one model had an estimated risk ≥10% and the other <10%.

#### Missing values for CA125

CA125 was missing for 1805 (31%) patients in the development data and for 966 (30%) patients in the validation data. Patients with tumors that looked suspicious for malignancy more often had CA125 measured. We used ‘multiple imputation by chained equations’ to deal with missing CA125 values. Imputation results, done separately for the development and validation data, were available from the original publications, see Supplementary Material 4 for details.[6, 8]

#### Modelling procedure and software

Supplementary Material 5 presents the modelling and validation procedure for models with CA125 and models without CA125. The analysis was performed with R version 4.1.2, using packages nnet (MLR), and caret together with packages glmnet (Ridge MLR), ranger (RF), xgboost, nnet (NN), and kernlab (SVM).[29] Meta-analysis for Net Benefit was performed using Winbugs.

## RESULTS

Descriptive statistics for the development and validation datasets are shown in Tables 1 and S1. A list of centers with distribution of the five tumor types is shown in Table S2. The median age of the patients was 47 years (interquartile range 35-60) in the development dataset and 49 years in the validation dataset (interquartile range 36-62). Most tumors were benign: 3980 (67%) in the development dataset and 1988 (62%) in the validation dataset. Secondary metastatic tumors were least common: 246 (4%) in the development dataset and 172 (5%) in the validation dataset.

**Table 1.** Descriptive statistics of predictors and outcome in the development and validation datasets.

| Characteristic | Development | External validation |
| --- | --- | --- |
| Number of patients | 5909 | 3199 |
| Number of centers | 24 | 25 |
| Number of oncology centers | 13<sup>a</sup> | 15 |
| Age (years) | 47 (35-60) | 49 (36-62) |
| Serum CA125 (U/mL) | 26 (13-109) | 25 (11-109) |
| Missing CA-125 | 1805 (31%) | 966 (30%) |
| Maximal diameter of lesion (mm) | 69 (48-100) | 71 (50-105) |
| Proportion of solid tissue | 0.11 (0-0.66) | 0.06 (0-0.67) |
| Papillary projections |  |  |
| 0 | 4771 (81%) | 2711 (85%) |
| 1 | 495 (8%) | 200 (6%) |
| 2 | 148 (3%) | 76 (2%) |
| 3 | 137 (2%) | 51 (2%) |
| >3 | 358 (6%) | 161 (5%) |
| >10 cyst locules | 471 (8%) | 311 (12%) |
| Ascites | 720 (12%) | 321 (10%) |
| Shadows | 742 (13%) | 464 (14%) |
| Tumor outcome |  |  |
| Benign | 3980 (67%) | 1988 (62%) |
| Borderline | 339 (6%) | 259 (8%) |
| Stage I invasive | 356 (6%) | 219 (7%) |
| Stage II-IV invasive | 988 (17%) | 561 (18%) |
| Secondary metastasis | 246 (4%) | 172 (5%) |

Results are shown as median (interquartile range) for continuous variables, and as n (%) for categorical variables.
<sup>a</sup> One center changed from a non-oncology center to an oncology center during the study period (Bologna, Italy). As a result, there were 24 centers yet 13 oncology and 12 non-oncology centers.

### Discrimination performance

For models with CA125, PDI ranged from 0.41 (95% CI 0.39-0.43) for SVM to 0.55 (0.51-0.60) for XGBoost (Table 2, Figure S1). In line with these results, the pairwise c-statistics were generally lower for SVM than for other models (Table S3). For the best models, pairwise c-statistics were above 0.90 for benign versus stage II-IV tumors, benign versus secondary metastatic tumors, benign versus stage I tumors, and borderline versus stage II-IV tumors. For all models, pairwise c-statistics were below 0.80 for borderline versus stage I tumors, stage I versus secondary metastatic tumors, and stage II-IV versus secondary metastatic tumors. The binary c-statistics (or AUROC) for any malignancy was 0.92 for all algorithms except Ridge MLR (0.90) and SVM (0.89) (Figure S2).

**Table 2.** Overview of discrimination, calibration, and utility performance on external validation data.

| Model | PDI<br>(95% CI) | Benign | BOT | ECI<br>Stage<br>I | Stage<br>II-IV | Sec.<br>meta | Mean | NB at<br>10%<br>(referrals<br>avoided) |
| --- | --- | --- | --- | --- | --- | --- | --- | --- |
| Models with CA125 |  |  |  |  |  |  |  |  |
| MLR | 0.54 (0.50-0.59) | 0.060 | 0.098 | 0.023 | 0.013 | 0.035 | 0.046 | 0.33 (0.23) |
| Ridge MLR | 0.49 (0.46-0.53) | 0.113 | 0.240 | 0.089 | 0.038 | 0.158 | 0.128 | 0.33 (0.21) |
| RF | 0.54 (0.50-0.59) | 0.014 | 0.083 | 0.020 | 0.0002 | 0.037 | 0.031 | 0.34 (0.28) |
| XGBoost | 0.55 (0.51-0.60) | 0.017 | 0.055 | 0.009 | 0.007 | 0.041 | 0.026 | 0.34 (0.27) |
| NN | 0.54 (0.50-0.58) | 0.042 | 0.114 | 0.058 | 0.005 | 0.155 | 0.075 | 0.33 (0.23) |
| SVM | 0.41 (0.39-0.43) | 0.161 | 0.179 | 0.271 | 0.279 | 0.069 | 0.192 | 0.33 (0.23) |
| Models without CA125 |  |  |  |  |  |  |  |  |
| MLR | 0.51 (0.47-0.54) | 0.065 | 0.131 | 0.043 | 0.029 | 0.013 | 0.056 | 0.34 (0.24) |
| Ridge MLR | 0.47 (0.44-0.49) | 0.038 | 0.206 | 0.073 | 0.004 | 0.111 | 0.086 | 0.34 (0.25) |
| RF | 0.50 (0.46-0.54) | 0.013 | 0.076 | 0.009 | 0.001 | 0.086 | 0.037 | 0.34 (0.24) |
| XGBoost | 0.50 (0.46-0.54) | 0.016 | 0.047 | 0.012 | 0.005 | 0.068 | 0.030 | 0.34 (0.25) |
| NN | 0.50 (0.46-0.54) | 0.048 | 0.049 | 0.048 | 0.008 | 0.089 | 0.048 | 0.33 (0.19) |
| SVM | 0.42 (0.39-0.45) | 0.215 | 0.254 | 0.414 | 0.228 | 0.022 | 0.227 | 0.33 (0.16) |

PDI, polytomous discrimination index; ECI, estimated calibration index; NB, net benefit; CI, confidence interval; Sec, secondary; MLR, multinomial logistic regression; RF, random forest; NN, neural network; SVM, support vector machine. Net Benefit: the Net Benefit of treat all is 0.31.
Referrals avoided: the net proportion of patients where an unnecessary referral (i.e. a false positive) was avoided relative to treat all (referring everyone).

For models without CA125, PDI ranged from 0.42 (95% CI 0.39-0.45) for SVM to 0.51 (0.47-0.54) for standard MLR (Table 2, Figure S3). Including CA125 mainly improved c-statistics for stage II-IV primary invasive vs secondary metastatic tumors, and stage I vs stage II-IV primary invasive tumors (Table S4). The binary c-statistics for any malignancy was less affected by excluding CA125, with values up to 0.91 (Figure S4).

### Calibration performance

For models with CA125, the probability of a benign tumor was too high on average for all algorithms, in particular for SVM (Figure 1). The risks of a stage I tumor and a secondary metastatic tumor were fairly well calibrated. The risk of a borderline tumor was slightly too low on average for all algorithms. The risk of a stage II-IV tumor was too low on average for standard MLR, Ridge MLR, and in particular for SVM. Based on the ECI, RF and XGBoost had the best calibration performance, SVM the worst (Table 2). Box plots of the estimated probabilities for each algorithm are presented in Figures S5-S10. For models without CA125, calibration results were roughly similar (Table 2, Figures S11-17).

**Figure 1.**
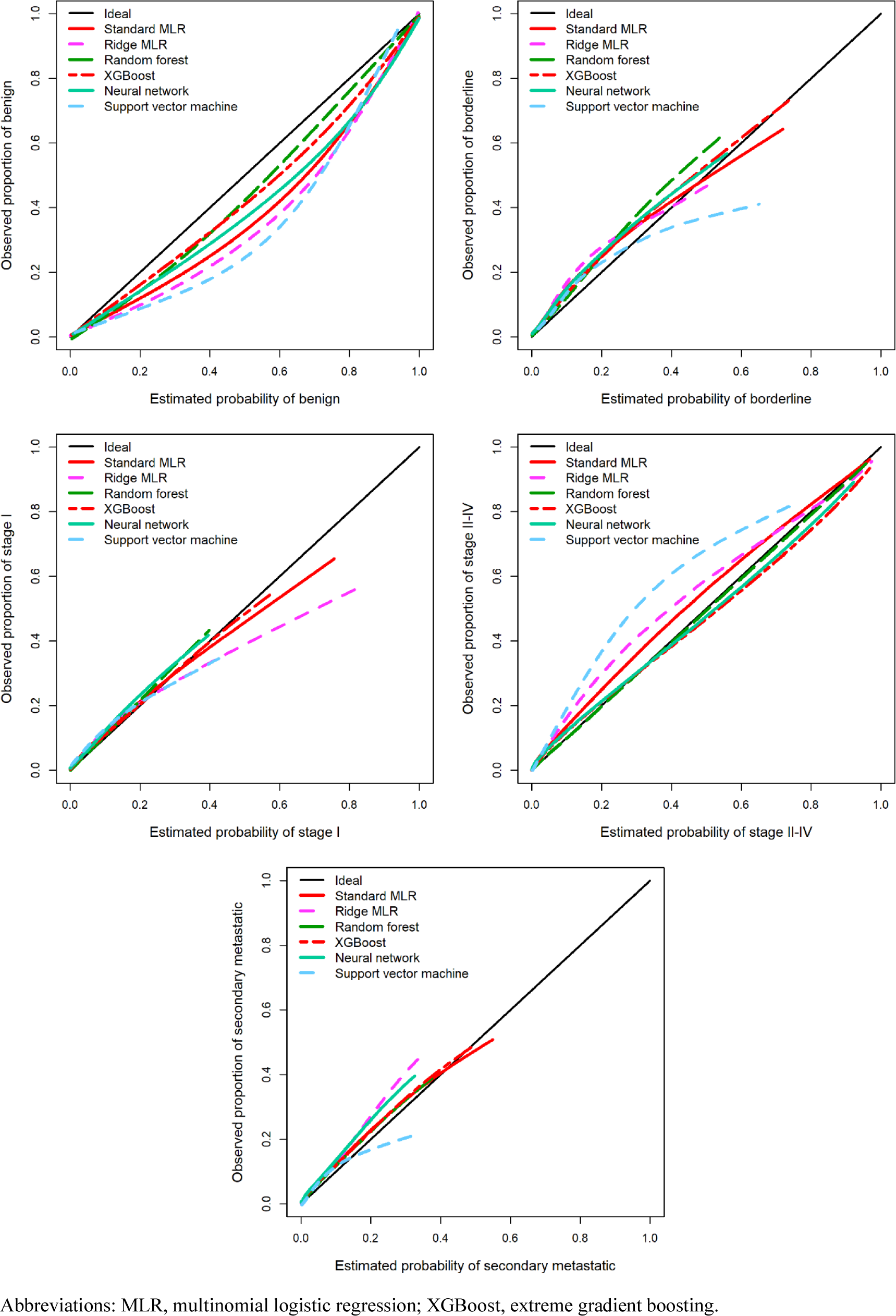
Flexible calibration curves for models with CA125 on external validation data.

### Clinical utility

All models with CA125 were superior to the default strategies (treat all, treat none) at any threshold (Figure S18). At the 10% threshold for the risk of any malignancy, all algorithms had similar Net Benefit (Table 2). At higher thresholds, RF and XGBoost had the best results, SVM the worst. For models without CA125, results were roughly similar (Table 2, Figure S19, Table S4).

### Comparing estimated probabilities between algorithms

For an individual patient, the six models could generate very different probabilities. For example, depending on the model, the estimated probability of a benign tumor differed at least 0.2 (20 percentage points) for 29% (models with CA125) and 31% (models without CA125) of the validation patients (Table 3, Figure S20). Scatter plots of estimated probabilities for each pair of models are provided in Figures 2-5 for models with CA125 and in Figures S21-S25 for models without. When comparing two models at the 10% threshold for the estimated risk of any malignancy, between 3% (XGBoost vs NN, with CA125) and 30% (NN vs SVM, without CA125) of patients fell on opposite sides of the threshold (Table S5).

**Figure 2.**
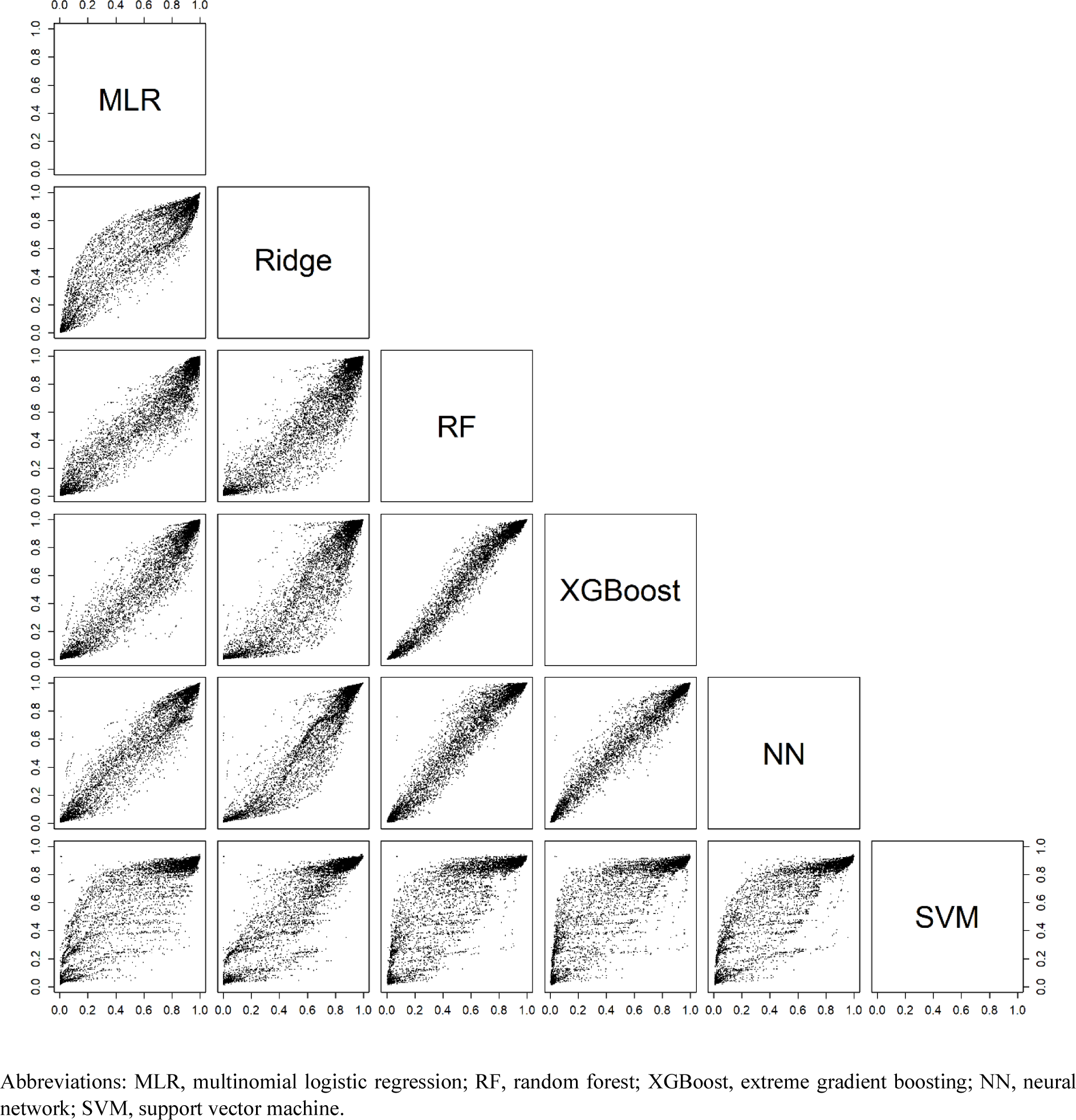
Scatter plots for the estimated probability of a benign tumor for each pair of models with CA125 on validation data.

**Figure 3.**
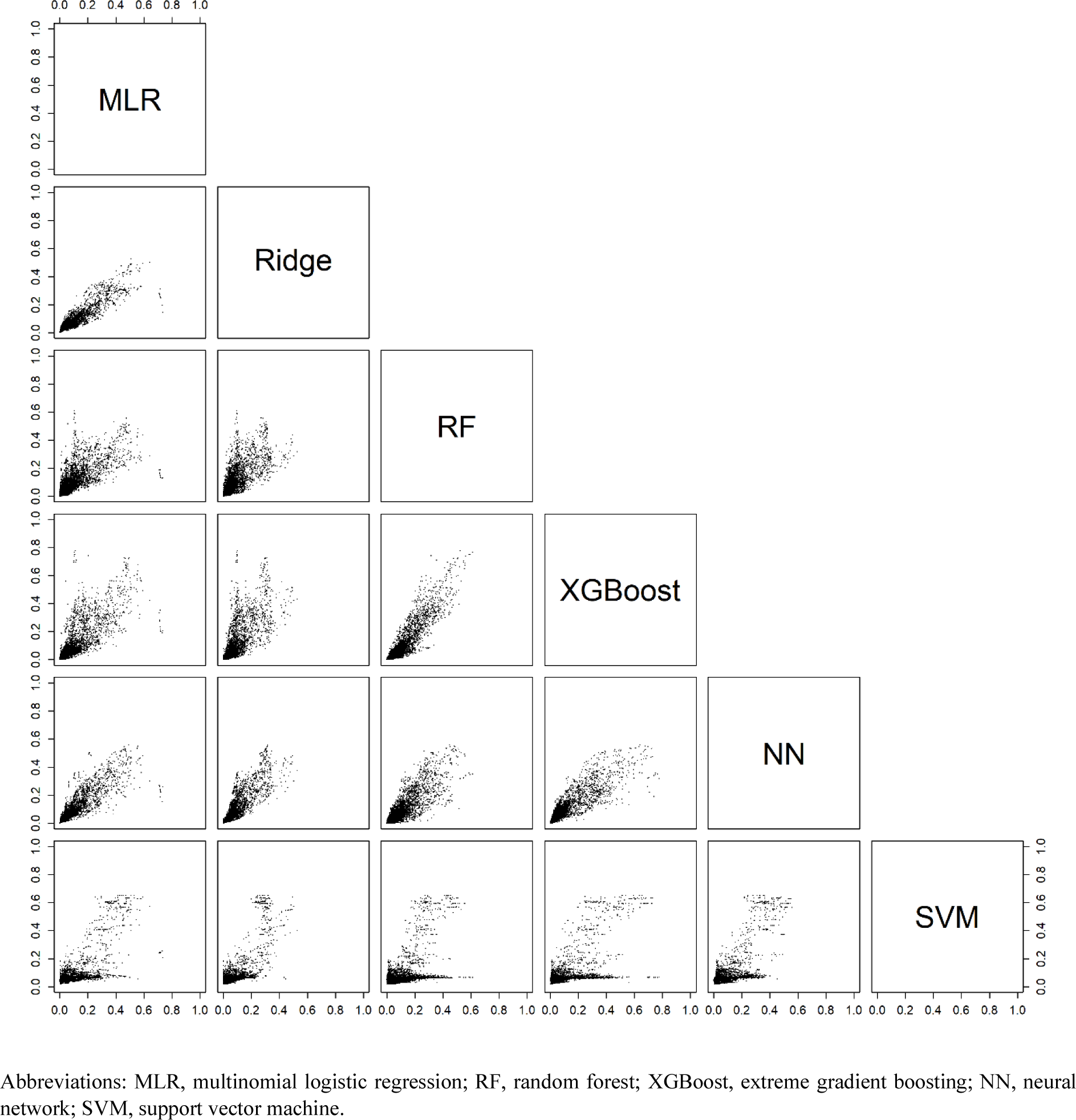
Scatter plots of the estimated risk of a borderline tumor for each pair of models with CA125 on validation data.

**Figure 4.**
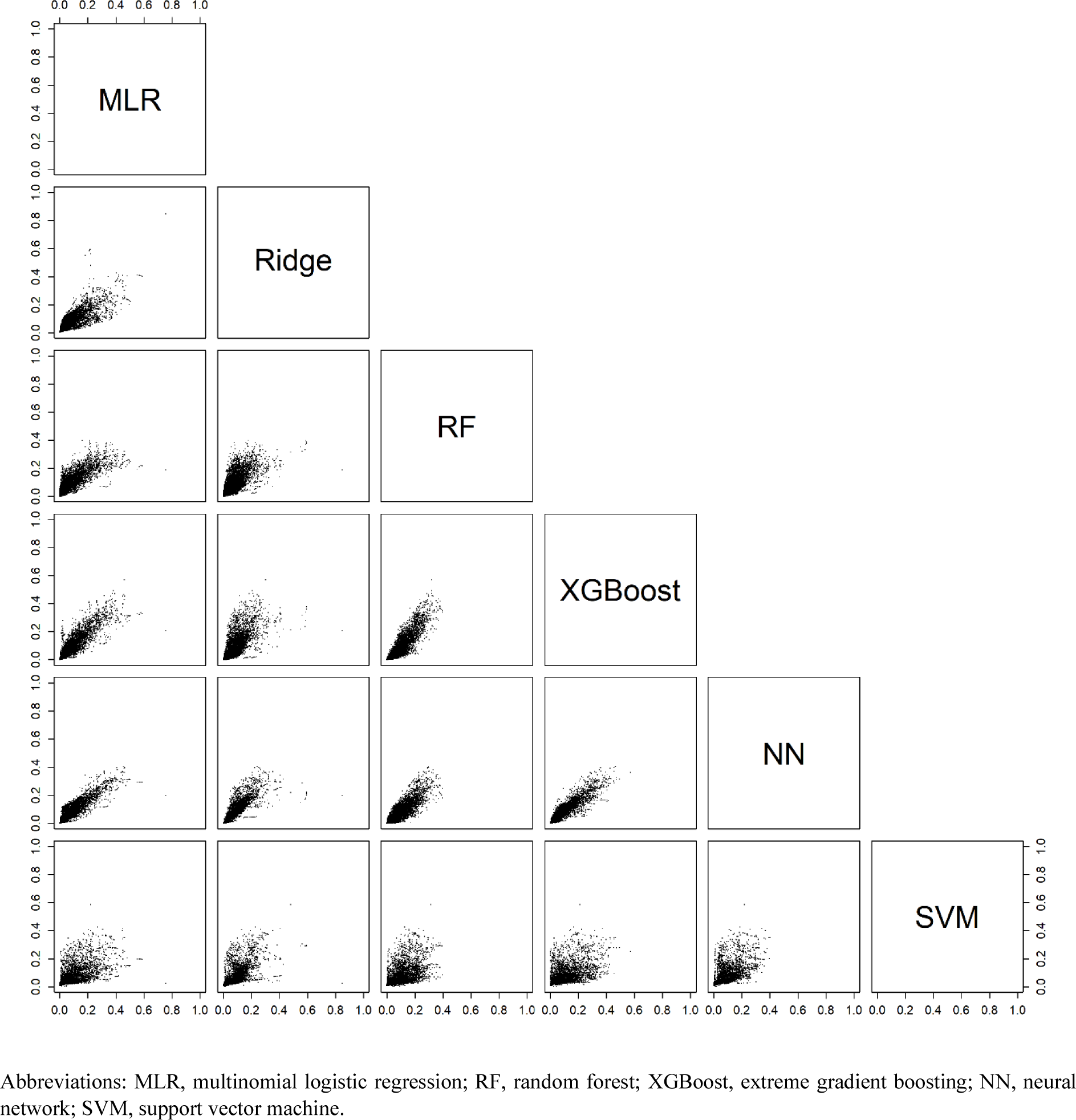
Scatter plots of the estimated risk of a stage I primary invasive tumor for each pair of models with CA125 on validation data.

**Figure 5.**
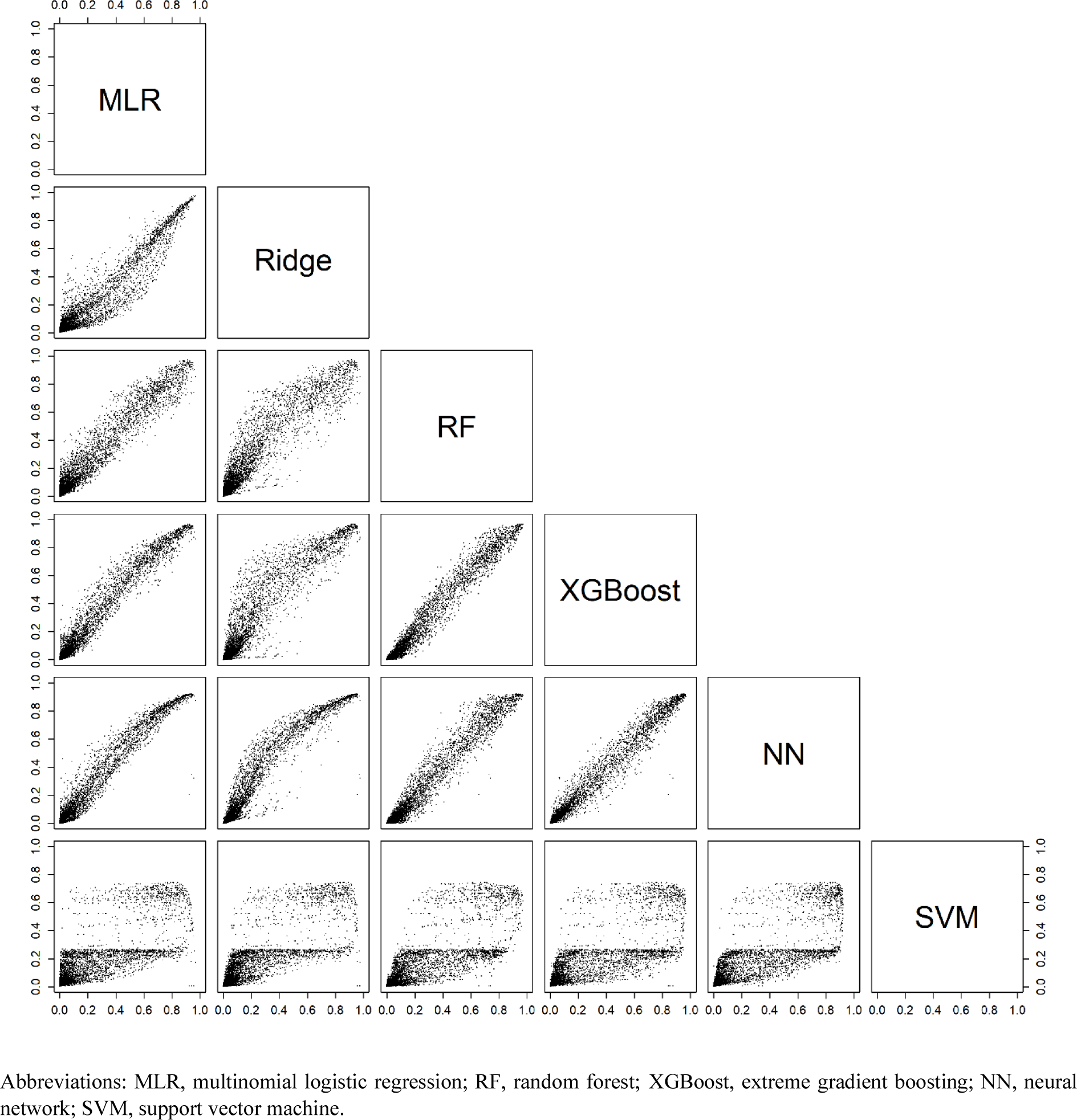
Scatter plots of the estimated risk of a stage II-IV primary invasive tumor for each pair of models with CA125 on validation data.

**Figure 6.**
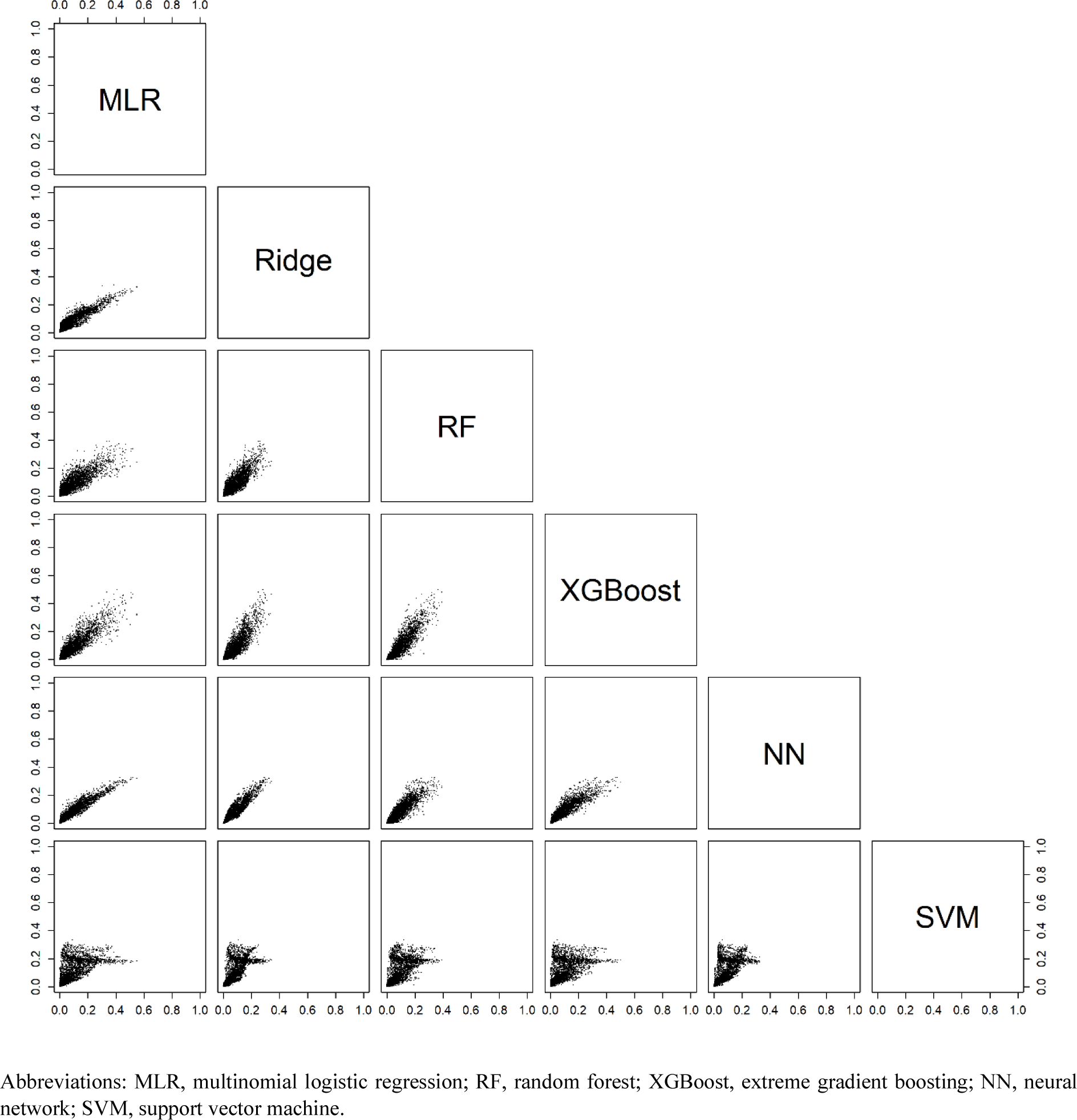
Scatter plots of the estimated risk of a secondary metastatic tumor for each pair of models with CA125 on validation data.

**Table 3.** Descriptive statistics of the probability range (highest minus lowest estimated probability) across the six models for each outcome on the external validation data.

| Range | Benign | Borderline | Stage I | Stage II-IV | Sec. meta |
| --- | --- | --- | --- | --- | --- |
| Models with CA125 |  |  |  |  |  |
| ≥0.05, n (%) | 3043 (95) | 1762 (55) | 1324 (41) | 1347 (42) | 919 (29) |
| ≥0.10, n (%) | 1467 (46) | 537 (17) | 569 (18) | 1033 (32) | 467 (15) |
| ≥0.20, n (%) | 927 (29) | 245 (8) | 116 (4) | 665 (21) | 87 (3) |
| ≥0.30, n (%) | 514 (16) | 95 (3) | 14 (0.4) | 430 (13) | 4 (0.1) |
| Models without CA125 |  |  |  |  |  |
| ≥0.05, n (%) | 3100 (97) | 1889 (59) | 1221 (38) | 1655 (52) | 924 (29) |
| ≥0.10, n (%) | 1673 (52) | 532 (17) | 550 (17) | 1301 (41) | 388 (12) |
| ≥0.20, n (%) | 990 (31) | 260 (8) | 106 (3) | 697 (22) | 12 (0.4) |
| ≥0.30, n (%) | 523 (16) | 121 (4) | 13 (0.4) | 229 (7) | 0 |

## DISCUSSION

We compared six algorithms to develop multinomial risk prediction models for ovarian cancer diagnosis. There was no algorithm that clearly outperformed the others. XGBoost, RF, NN and MLR had similar performance, SVM had the worst performance. CA125 mainly increased discrimination between stage II-IV primary invasive tumors and the other two types of invasive tumors. Despite similar performance for several algorithms, the choice of algorithm had a clear impact on the estimated probabilities for individual patients. Choosing a different algorithm could lead to different clinical decisions in a substantial percentage of patients.

Strengths of the study include (1) the use of large international multicenter datasets, (2) data collection according to a standardized ultrasound examination technique, measurement technique, and terminology,[24] (3) evaluation of risk calibration and clinical utility, and (4) appropriate modeling practices by addressing nonlinearity for continuous predictors in regression models and hyperparameter tuning for the machine learning algorithms. Such modeling practices are often lacking in comparative studies.[10] A limitation could be that we included only patients that received surgery, thereby excluding patients managed conservatively. This limitation affects most studies on ovarian malignancy diagnosis, because surgery allows using histopathology to determine outcome. The use of a fixed set of predictors could also be perceived as a limitation. However, these predictors were carefully selected based largely on expert domain knowledge for development of the ADNEX model, which is perhaps the best performing ultrasound-based prediction model to date.[6,8,40] Including more predictors, or using a data-driven selection procedure per algorithm, would likely increase the observed differences in estimated probabilities between algorithms.

Previous studies developed machine learning models using sonographic and clinical variables to estimate the risk of malignancy in adnexal masses on smaller datasets (median sample size 357, range 35-3004).[41–52] Calibration was not assessed, and the outcome was binary (usually benign vs malignant) in all but two studies. One study distinguished between benign, borderline, and invasive tumors,[45] another study distinguished between benign, borderline, primary invasive, and secondary metastatic tumors.[44] However, sample size was small in these two studies (the smallest outcome category had 16 and 30 cases, respectively, in the development set). All studies focused exclusively on neural networks, support vector machines, or related kernel-based methods. All but one of these studies implicitly or explicitly supported the use of machine learning algorithms over logistic regression.

Our results illustrate that the probability estimates for individual patients can vary substantially by algorithm. There are different types of uncertainty of individual predictions.[53] ‘Aleatory uncertainty’ implies that two patients with the same predictor measurements (same age, same maximum lesion diameter, etcetera) may have a different outcome. ‘Epistemic uncertainty’ refers to lack of knowledge about the best final model and is divided into ‘approximation uncertainty’ and ‘model uncertainty’.[53] ‘Approximation uncertainty’ reflects sample size: the smaller the sample size, the more uncertain the developed model. This means that very different models can be obtained when fitting the same algorithm to different training datasets of the same size, and that these differences become smaller with increasing sample size. ‘Model uncertainty’ reflects the impact of various decisions made during model development. Our study illustrates that model certainty strongly relates to the decision of which algorithm to use for model development.

A first implication of our work is that there is no important advantage of using flexible machine learning over multinomial logistic regression for developing ultrasound-based risk models for ovarian cancer diagnosis to support clinical decisions. An MLR-based model is easier to implement, update, and explain than a flexible machine learning model. We would like to emphasize that the ADNEX model that was mentioned in the introduction, although based on MLR, includes random intercepts by center.[6] This is an advantage because it acknowledges that prevalences of the outcome categories vary between centers.[58] We did not use random intercepts in the current study, because they do not generalize directly to flexible algorithms. A second implication is that the choice of algorithm matters for individual predictions, even when discrimination, calibration, and clinical utility are similar. Different models with equal clinical utility in the population may yield very different risk estimates for an individual patient, and this may lead to different management decisions for the same individual. Although, in our opinion, the crux of clinical risk prediction models is that their use should lead to improved clinical decisions for a specific population as a whole, differences in risk estimates for the same individual are an important finding. More research is needed to better understand uncertainty in predictions caused by the choice of algorithm, or other decisions made by the modeler such as the predictor selection method. The observation that different algorithms may make different predictions emphasizes the need of sufficiently large databases when developing prediction models. The recently established guidance for minimum sample size to develop a regression-based prediction model is a crucial step forward.[59,60] However, it is based on general performance measures related to discrimination and calibration, and does not cover uncertainty of risk estimates for individual patients. Hence, if possible, the sample size should be larger than what the guidance would suggest. Flexible machine learning algorithms may require even more data than regression algorithms.[61] We should consider providing an indication of the uncertainty around a risk estimate. Confidence intervals around the estimated probabilities may be provided, although this may be confusing for patients.[62] Moreover, standard confidence intervals do not capture all sources of uncertainty. Related options may be explored, such as models that abstain from making predictions when uncertainty is too large.[14]

## CONCLUSION

Several algorithms had similar performance and good clinical utility to estimate the probability of five tumor types in women with an adnexal (ovarian, para-ovarian, or tubal) mass treated with surgery. However, different algorithms could yield very different probabilities for individual patients.

## AUTHOR CONTRIBUTIONS

Contributions were based on the CRediT taxonomy. Conceptualization: AL, JC, BVC. Data curation: JC, WF. Formal analysis: AL, JC, BVC. Funding acquisition: LV, WF, DT, BVC. Investigation: LV, AT, CVH, DF, TB, WF, DT. Methodology: AL, BVC. Project administration: WF, DT, BVC. Resources: LV, AT, CVH, DF, TB, WF, DT. Software: AL, JC. Supervision: DT, BVC. Validation: BVC. Visualization: AL, BVC. Writing – original draft: AL, JC, BVC. Writing – review & editing: all authors. AL, JC, WF, DT, BVC directly accessed and verified the raw data, and no authors were precluded from accessing the data. All authors have read, share final responsibility for the decision to submit for publication, and agree to be accountable for all aspects of the work in ensuring that questions related to the accuracy or integrity of any part of the work are appropriately investigated and resolved.

## DATA AVAILABILITY

The analysis code and statistical analysis plan are available on GitHub (https://github.com/AshleighLedger/Paper-IOTA-ML). The datasets that we analysed during the current study are not publicly available because this was not part of the informed consent at the time (the last patient was recruited in 2015). However, the dataset may be obtained following permission of prof. Dirk Timmerman and after fulfilling all requirements such as data transfer agreements or ethics approval from the leading ethics committee during data collection (Research Ethics Committee of the University Hospitals Leuven).

