## Supplementary material for "Multiclass risk models for ovarian malignancy: an illustration of prediction uncertainty due to the choice of algorithm"

### Table of contents

Supplementary Material 1. Predictor selection

Supplementary Material 2. Sample size argumentation

Supplementary Material 3. Hyperparameter tuning

Supplementary Material 4. Multiple imputation for CA125

Supplementary Material 5. Flowcharts for modeling and validation procedure

Table S1. Descriptive statistics by reference standard (final diagnosis).

Table S2. List of centers in the development and validation data.

Table S3. Pairwise area under the receiver operating characteristic curve (AUROC) values (with 95% CI) for models with CA125 on external validation data.

Table S4. Pairwise area under the receiver operating characteristic curve (AUROC) values for models without CA125.

Table S5. Percentage of patients on validation data falling on opposite sides of the 10% risk of malignancy threshold when comparing two models.

Figure S1. Polytomous discrimination index for models with CA125 on external validation data.

Figure S2. AUROC for benign tumors vs any malignancy for models with CA125.

Figure S3. Polytomous Discrimination Index (PDI) for models without CA125.

Figure S4. AUROC for benign tumors vs any malignancy for models without CA125.

Figure S5. Box plots of estimated probabilities for standard MLR with CA125.

Figure S6. Box plots of estimated probabilities for ridge MLR with CA125.

Figure S7. Box plots of estimated probabilities for random forest with CA125.

Figure S8. Box plots of estimated probabilities for extreme gradient boosting (XGBoost) with CA125.

Figure S9. Box plots of estimated probabilities for neural network with CA125.

Figure S10. Box plots of estimated probabilities for support vector machine with CA125.

Figure S11. Flexible calibration curves for models without CA125.

Figure S12. Box plots of estimated probabilities for standard MLR without CA125.

Figure S13. Box plots of estimated probabilities for ridge MLR without CA125.

Figure S14. Box plots of estimated probabilities for random forest without CA125.

Figure S15. Box plots of estimated probabilities for extreme gradient boosting (XGBoost) without CA125.

Figure S16. Box plots of estimated probabilities for neural network without CA125.

Figure S17. Box plots of estimated probabilities for support vector machine without CA125.

Figure S18. Decision curves for models with CA125 on external validation data.

Figure S19. Decision curves for models without CA125 on external validation data.

Figure S20. Differences between the highest and lowest estimated probability for each outcome across the six models with CA125 (panel A) and the six models without CA125 (panel B) for patients in the external validation dataset. Each dot denotes the difference between the highest and the lowest estimated probability for one patient. This means that each patient is shown five times in each panel, once for each outcome category. For example, at the far left, the difference between the highest and lowest estimated probability for a benign tumor is shown for all 3199 patients in the dataset. The box represents the interquartile range which contains the middle 50% of the differences. The line inside the box indicates the median. Whiskers correspond to the 5th and 95th percentile.

Figure S21. Scatter plots of the estimated risk of a benign tumor for each pair of models without CA125.

Figure S22. Scatter plots of the estimated risk of a borderline tumor for each pair of models without CA125.

Figure S23. Scatter plots of the estimated risk of a stage I primary invasive tumor for each pair of models wi without th CA125.

Figure S24. Scatter plots of the estimated risk of a stage II-IV primary invasive tumor for each pair of models without CA125.

Figure S25. Scatter plots of the estimated risk of a secondary metastatic tumor for each pair of models without CA125.

References for supplementary material

**Supplementary Material 1. Predictor selection for the ADNEX model.**

As described in the original publication from 2014, the variable selection for the development of the ADNEX model was done in two stages.[1] First, 10 variables were selected based on expert domain knowledge and stability of the values across centers.[1,2] To select variables a priori, clinicians (DT, TB, CVH, LV) with substantial experience in evaluating and diagnosing adnexal masses assessed variables in terms of their likely value to differentiate between the five outcome categories, subjectivity, and measurement cost. Stability across centers was studied by studying heterogeneity of values between centers after correcting for tumor outcome.[2] We favored predictors with limited heterogeneity, irrespective of the cause of the heterogeneity. Predictor values may differ between centers due to subjectivity of the predictor, dependence on experience, differences regarding local procedures or equipment, or case-mix differences.

Second, data-driven selection was performed using multivariate fractional polynomials.[3] This method combines backward variable elimination with selection of optimal transformations of numerical/continuous variables. To accommodate the multiclass nature of the outcome, we applied this procedure in the context of four binary logistic regression models (using random intercepts per center): borderline vs benign, stage I invasive vs benign, stage II-IV invasive vs benign, and secondary metastasis vs benign. To eliminate variables as well as to select the optimal transformation, we used an alpha level that corresponded to the Bayesian Information Criterion.[4] The final set of predictors included predictors that were selected in at least one of the four submodels. If different transformations were selected for a predictor in the four submodels, one final transformation was chosen manually. Two predictors could not be eliminated because we wanted to include them anyway: age and type of center. This means that the alpha level for these variables in the backward elimination process was set to 1.

The multivariable fractional polynomial procedure selected nine variables. Only personal history of ovarian cancer (yes/no) was excluded. The following transformations were selected for numerical predictors: maximum lesion diameter and CA125 were log-transformed, proportion solid tissue was modeled using a linear and quadratic term, and age and number of papillations were modeled linearly (i.e. no transformation was selected).[1]

### Supplementary Material 2. Sample size argumentation

1. Model development (N=5909)

This retrospective study develops models on the same data as were used to develop the ADNEX model.[1] This involves 5909 patients, of which 3980 with a benign, 339 with a borderline, 356 with a stage I invasive, 988 with a stage II-IV invasive, and 246 with a secondary metastatic tumor. Recently, sample size calculation methods have been published for the development of logistic regression models for binary as well as multinomial outcomes.[5,6]

To develop the ADNEX model, data-driven selection was performed on 10 predictors, using 17 parameters in total in order to address nonlinearity for continuous covariates.[1] To develop a logistic regression model for the binary outcome benign vs malignant, we further require the outcome event fraction and an estimate of the Cox-Snell R-squared.[5] The event fraction of malignant tumors is 33% (1929/5909). We can assume that the area under the receiver operating characteristic curve (AUROC) is at least 0.90,[7] yielding a Cox-Snell R-squared of approximately 0.412.[8] Based on the four criteria from Riley et al,[5] this leads to a minimum sample size of 789, when targeting a minimum expected calibration slope of 0.9 (to minimize overfitting in the predictor effects), a maximum expected margin of error of 5% for the estimate of the overall event fraction, a maximum expected optimism in the apparent R-squared of 0.05, and a mean absolute prediction error of 0.05.

A novel methodology for determining sample size for multinomial logistic regression models focuses on three criteria: (i) minimizing overfitting in the predictor effects for every pair of outcome categories (i.e. minimum calibration slope of 0.9 recommended), (ii) limiting the optimism in the apparent R-squared (maximum 0.05 recommended), and (iii) estimating the overall event fractions of all outcome categories with small margin of error (maximum 5% recommended).[6] Criterion ii leads to a required sample size of 1416, and criterion iii to 587. Criterion i is rather strict. We can use estimates for the pairwise Cox-Snell R-squared values based on the pairwise AUROC values reported for the validation data in the ADNEX study.[1] Using these estimates, we can target calibration slopes of 0.9 for eight outcome pairs, 0.89 for one pair (borderline vs stage I invasive) and 0.82 for one pair (stage I invasive vs secondary metastasis).

In sum, sample size is sufficient for robust model development, although allowing some flexibility for the comparison between stage I invasive tumors and secondary metastatic tumors. Note that these sample size calculations focus on regression models only.

2. External validation (n=3199)

The common rule of thumb for externally validating a model for a binary outcome is to have 100, but preferably 200, observations in the smallest outcome groups.[9,10] The rule of thumb to have at least 200 in each category is mainly based on having sufficiently stable calibration curves. Recently, a dedicated sample size calculation method for the external validation of a clinical prediction model with binary outcome was published.[11] Riley et al (2021) proposed iterative and closed form solutions to compute the minimum sample size required to precisely estimate the ratio between the observed and expected number of event outcomes (O/E ratio) and calibration slope (calibration), the AUROC (discrimination) and net benefit (clinical utility).[11] These calculations require the specification of the anticipated outcome event proportion, the confidence intervals width, the anticipated value for the estimates and the anticipated distribution of the linear predictor values.

Calculations suggest that at least 628 participants (and 239 malignant cases) are required to precisely estimate an O/E ratio with a 95% CI width of 0.20 assuming that the expected O/E ratio is 1. At least 1479 participants (562 malignant cases) are required to precisely estimate a calibration slope with a CI width of 0.20 assuming that the expected slope is 1, and that the expected distribution of linear predictor is skew normal (mean= 1.5, sd=2.5, skewness parameter=-0.9 for patients with a malignant tumor, and mean=-3.3, sd=1.1, skewness parameter=1.2, for patients with a benign mass). For the AUROC, at least 168 participants (including 64 malignant cases) are required to estimate an expected AUROC of 0.90 with a 95% CI width of 0.10. For the net benefit, the minimum number of participants required was the highest at 50% decision threshold (on the risk of malignancy) with a CI width of 0.20 and an anticipated sensitivity of 71% and specificity of 93%. To estimate net benefit with reasonable precision, at least 337 participants and 129 events are required. Therefore, our sample size of 3199 including 1211 malignant outcomes is sufficient to estimate the previously mentioned estimates with reasonable precision.

There is no sample size methodology for the external validation of multiclass models. However, we can address adequacy of the sample size for the current study based on a validation study of ADNEX that used 2489 of the 3199 patients in the current validation dataset.[7] In this validation study of ADNEX, the reported AUROCs between each pair of outcomes had 95% confidence intervals with a width between 0.01 and 0.10, which suggests sufficient precision. It is difficult to make claims about the multinomial calibration curves. We can only state that four outcome categories had more than 200 cases (1988 benign, 561 stage II-IV primary invasive, 259 borderline, 219 stage I primary invasive), and secondary metastasis had 172 cases. This corresponds to the abovementioned rule of thumb to have at least 100 but preferably 200 cases in each outcome category.[9,10]

### Supplementary Material 3. Hyperparameter tuning

Machine learning algorithms have hyperparameters for which a value has to be selected. Instead of choosing default levels, the hyperparameters can be tuned to optimize the performance of the algorithm. The hyperparameter set $\lambda$ is estimated using the following optimization problem:[12]

$$\hat{\lambda} \equiv \underset{\lambda \in \{\lambda^{(1)} \ldots \lambda^{(S)}\}}{\operatorname{argmin}} \Psi(\lambda),$$

where $\Psi(\cdot)$ is the loss function, and $\lambda^{(1)}$ to $\lambda^{(S)}$ the set of trials over which to evaluate the loss function.[12] An important step is the selection of the set of trials, and there exists several methods to do so, such as grid, manual and random search.[12] Default values provided by software can be used as well, however hyperparameters differ between datasets and should be adapted to the data.[13]

We trained ridge, random forest, XGBoost, neural network and support vector machine with the R package caret (function “train”).[14] We used the function “glmnet” for ridge, “ranger” for random forests, “xgbTree” for XGBoost, “nnet” for neural networks, and “svmRadial” for support vector machines with radial basis function kernel respectively (requiring packages glmnet, ranger, xgboost, nnet, and kernlab). To tune the hyperparameters, we used the random or grid search method with 30 trials (i.e. 30 randomly chosen combinations of hyperparameters). The best combination of hyperparameters was selected based on the logloss (also known as entropy or log-likelihood) through a 10-fold cross-validation procedure on the development dataset. The average test set logloss across the 10 folds was calculated to determine the best combination of hyperparameters. Using the best combination of hyperparameters for a given algorithm, a model was then developed on the complete development dataset.

Bergstra and Bengio (2012) define the random search method as “independent draws from a uniform density from the same configuration space as would be spanned by a regular grid”.[12] At each instance, random values of hyperparameters are selected over the parameter space. The random draws with replacement ensure that the trials are statistically independent. The search space over which the draws are randomly made for each hyperparameter in the package caret is shown in Table SM3.1. The range of values was determined by caret.

Tables SM3.2-6 present the 30 trials for each model, and the obtained average logloss for the models that use CA125 as a predictor. Tables SM3.7-11 present the 30 trials for each model, and the obtained average logloss for the models that do not use CA125 as a predictor.

**Table SM3.1: Hyperparameters used together with their range to determine the parameter space for the random and grid search tuning procedure.**

| Algorithm | Hyperparameter | Hyperparameter meaning | Range of values |
|---|---|---|---|
| Ridge | lambda | Regularization parameter | 2^Uniform[-10,3] |
| Random forest | min.node.size | minimum size of terminal nodes | 1 to 20 |
| | mtry | number of variables randomly sampled as candidates at each split | 1 to 11[a] |
| | splitrule | How splits within each tree are chosen | "gini", "extratrees"[b] |
| XGBoost | eta | step size shrinkage used in update to prevent overfitting | Uniform[0.001, 0.6] |
| | max_depth | maximum depth of a tree | 1 to 10 |
| | gamma | minimum loss reduction required to make a further partition on a leaf node | Uniform[0, 10] |
| | colsample_bytree | subsample ratio of columns when constructing each tree | Uniform[0.7, 7] |
| | min_child_weight | minimum sum of instance weight needed in a child | 0 to 20 |
| | subsample | subsample ratio of the training instances | Uniform[0.25, 1] |
| | nrounds | number of decision trees in the final model | 1 to 1000 |
| Neural network | size | number of units in the hidden layer | 1 to 10 |
| | decay | weight decay | 10^Uniform[-5, 1] |
| Support vector machine | sigma | inverse kernel width for the radial basis kernel function | Exp(Uniform[-6.1, 0.21]) |
| | C | regularisation parameter | 2^Uniform[-5, 10] |

[a] mtry goes up to 11 because the number of papillations is represented through 4 dummy variables.

[b] gini refers to the standard approach, extratrees refers to the approach of extremely randomized trees in which splits are chosen randomly.[15]

**Table SM3.2. Grid search parameter values and average logloss for Ridge MLR with CA125 as a predictor.** Missing data for CA125 were addressed using multiple imputation, hence the 10-fold cross-validation procedure was applied in each of the 10 imputed datasets. The best hyperparameter value is marked in bold.

| | **Average logloss for the 10 imputed datasets** | | | | | | | | | |
|---|---|---|---|---|---|---|---|---|---|---|
| **Lambda** | **1** | **2** | **3** | **4** | **5** | **6** | **7** | **8** | **9** | **10** |
| 0.00099 | 0.60 | 0.60 | 0.60 | 0.60 | 0.60 | 0.60 | 0.60 | 0.60 | 0.60 | 0.60 |
| 0.00117 | 0.60 | 0.60 | 0.60 | 0.60 | 0.60 | 0.60 | 0.60 | 0.60 | 0.60 | 0.60 |
| 0.00178 | 0.60 | 0.60 | 0.60 | 0.60 | 0.60 | 0.60 | 0.60 | 0.60 | 0.60 | 0.60 |
| 0.00340 | 0.60 | 0.60 | 0.60 | 0.60 | 0.60 | 0.60 | 0.60 | 0.60 | 0.60 | 0.60 |
| 0.00549 | 0.60 | 0.60 | 0.60 | 0.60 | 0.60 | 0.60 | 0.60 | 0.60 | 0.60 | 0.60 |
| 0.01265 | 0.60 | 0.60 | 0.60 | 0.60 | 0.60 | 0.60 | 0.60 | 0.60 | 0.60 | 0.60 |
| **0.01641** | **0.60** | **0.60** | **0.60** | **0.60** | **0.60** | **0.60** | **0.60** | **0.60** | **0.60** | **0.60** |
| 0.02860 | 0.60 | 0.60 | 0.60 | 0.60 | 0.60 | 0.60 | 0.60 | 0.60 | 0.60 | 0.60 |
| 0.0520 | 0.61 | 0.61 | 0.61 | 0.61 | 0.61 | 0.61 | 0.61 | 0.61 | 0.61 | 0.61 |
| 0.08061 | 0.62 | 0.62 | 0.62 | 0.62 | 0.62 | 0.62 | 0.62 | 0.62 | 0.62 | 0.62 |
| 0.10881 | 0.63 | 0.64 | 0.63 | 0.64 | 0.64 | 0.63 | 0.63 | 0.63 | 0.63 | 0.63 |
| 0.13585 | 0.64 | 0.64 | 0.64 | 0.64 | 0.64 | 0.64 | 0.64 | 0.64 | 0.64 | 0.64 |
| 0.14604 | 0.65 | 0.65 | 0.65 | 0.65 | 0.65 | 0.65 | 0.65 | 00.65 | 0.64 | 0.64 |
| 0.18028 | 0.66 | 0.66 | 0.66 | 0.66 | 0.66 | 0.66 | 0.66 | 0.66 | 0.64 | 0.65 |
| 0.18647 | 0.66 | 0.66 | 0.66 | 0.66 | 0.66 | 0.66 | 0.66 | 0.66 | 0.66 | 0.66 |
| 0.18671 | 0.66 | 0.66 | 0.66 | 0.66 | 0.66 | 0.66 | 0.66 | 0.66 | 0.66 | 0.66 |
| 0.25463 | 0.68 | 0.68 | 0.68 | 0.68 | 0.68 | 0.68 | 0.68 | 0.68 | 0.67 | 0.67 |
| 0.32586 | 0.69 | 0.69 | 0.69 | 0.69 | 0.69 | 0.69 | 0.69 | 0.69 | 0.69 | 0.69 |
| 0.52 | 0.73 | 0.73 | 0.73 | 0.73 | 0.73 | 0.73 | 0.73 | 0.73 | 0.72 | 0.73 |
| 0.6282 | 0.74 | 0.74 | 0.74 | 0.74 | 0.74 | 0.74 | 0.74 | 0.74 | 0.74 | 0.74 |
| 0.76854 | 0.76 | 0.76 | 0.76 | 0.76 | 0.76 | 0.76 | 0.76 | 0.76 | 0.76 | 0.76 |
| 0.80835 | 0.79 | 0.76 | 0.76 | 0.76 | 0.76 | 0.76 | 0.76 | 0.76 | 0.76 | 0.76 |
| 1.06 | 0.79 | 0.79 | 0.79 | 0.79 | 0.79 | 0.79 | 0.79 | 0.79 | 0.79 | 0.79 |
| 1.12 | 0.84 | 0.79 | 0.79 | 0.79 | 0.79 | 0.79 | 0.79 | 0.79 | 0.79 | 0.79 |
| 1.87 | 0.87 | 0.84 | 0.84 | 0.84 | 0.84 | 0.84 | 0.84 | 0.84 | 0.84 | 0.84 |
| 2.37 | 0.87 | 0.87 | 0.87 | 0.87 | 0.87 | 0.87 | 0.87 | 0.87 | 0.86 | 0.86 |
| 2.52 | 0.88 | 0.87 | 0.87 | 0.87 | 0.87 | 0.87 | 0.87 | 0.87 | 0.87 | 0.87 |
| 2.78 | 0.91 | 0.88 | 0.88 | 0.88 | 0.88 | 0.88 | 0.88 | 0.88 | 0.88 | 0.88 |
| 4.17 | 0.91 | 0.92 | 0.92 | 0.91 | 0.92 | 0.91 | 0.91 | 0.91 | 0.91 | 0.91 |
| 4.23 | 0.92 | 0.92 | 0.92 | 0.92 | 0.92 | 0.92 | 0.92 | 0.92 | 0.92 | 0.92 |

**Table SM3.3. Randomly drawn hyperparameter combinations and average logloss for the random forest with CA125 as a predictor.** Missing data for CA125 were addressed using multiple imputation, hence the 10-fold cross-validation procedure was applied in each of the 10 imputed datasets. The best hyperparameter combination is marked in bold.

| | | | **Average logloss for the 10 imputed datasets** | | | | | | | | | |
|---|---|---|---|---|---|---|---|---|---|---|---|---|
| **mtry** | **min.node.size** | **splitrule** | **1** | **2** | **3** | **4** | **5** | **6** | **7** | **8** | **9** | **10** |
| 1 | 16 | extratrees | 0.78 | 0.78 | 0.78 | 0.78 | 0.78 | 0.78 | 0.78 | 0.78 | 0.78 | 0.78 |
| 1 | 4 | gini | 0.71 | 0.71 | 0.71 | 0.71 | 0.71 | 0.71 | 0.71 | 0.71 | 0.71 | 0.71 |
| 1 | 14 | gini | 0.71 | 0.71 | 0.71 | 0.71 | 0.71 | 0.71 | 0.71 | 0.71 | 0.71 | 0.71 |
| 1 | 15 | gini | 0.71 | 0.71 | 0.71 | 0.71 | 0.71 | 0.71 | 0.71 | 0.71 | 0.71 | 0.71 |
| 2 | 9 | extratrees | 0.65 | 0.65 | 0.65 | 0.65 | 0.65 | 0.65 | 0.65 | 0.65 | 0.65 | 0.65 |
| 3 | 5 | extratrees | 0.61 | 0.61 | 0.61 | 0.61 | 0.61 | 0.6 | 0.61 | 0.6 | 0.6 | 0.6 |
| 3 | 4 | gini | 0.58 | 0.58 | 0.58 | 0.58 | 0.58 | 0.58 | 0.58 | 0.58 | 0.57 | 0.58 |
| 3 | 7 | gini | 0.58 | 0.58 | 0.58 | 0.58 | 0.58 | 0.58 | 0.58 | 0.58 | 0.57 | 0.58 |
| **3** | **15** | **gini** | **0.58** | **0.58** | **0.58** | **0.58** | **0.58** | **0.57** | **0.58** | **0.58** | **0.57** | **0.57** |
| 5 | 5 | extratrees | 0.6 | 0.6 | 0.59 | 0.59 | 0.6 | 0.59 | 0.59 | 0.59 | 0.59 | 0.59 |
| 6 | 4 | extratrees | 0.61 | 0.61 | 0.61 | 0.61 | 0.6 | 0.61 | 0.6 | 0.61 | 0.6 | 0.6 |
| 6 | 8 | extratrees | 0.6 | 0.6 | 0.59 | 0.59 | 0.6 | 0.59 | 0.59 | 0.59 | 0.59 | 0.59 |
| 6 | 3 | gini | 0.66 | 0.64 | 0.65 | 0.65 | 0.66 | 0.65 | 0.63 | 0.65 | 0.64 | 0.64 |
| 6 | 6 | gini | 0.63 | 0.64 | 0.63 | 0.63 | 0.62 | 0.63 | 0.63 | 0.65 | 0.63 | 0.63 |
| 7 | 12 | gini | 0.66 | 0.64 | 0.65 | 0.64 | 0.64 | 0.66 | 0.64 | 0.64 | 0.64 | 0.65 |
| 8 | 2 | extratrees | 0.66 | 0.65 | 0.65 | 0.66 | 0.66 | 0.64 | 0.65 | 0.65 | 0.63 | 0.65 |
| 8 | 9 | extratrees | 0.61 | 0.61 | 0.61 | 0.62 | 0.61 | 0.6 | 0.61 | 0.6 | 0.6 | 0.6 |
| 8 | 5 | gini | 0.72 | 0.73 | 0.72 | 0.73 | 0.72 | 0.71 | 0.71 | 0.72 | 0.71 | 0.72 |
| 8 | 14 | gini | 0.67 | 0.66 | 0.66 | 0.66 | 0.65 | 0.66 | 0.66 | 0.66 | 0.66 | 0.64 |
| 8 | 20 | gini | 0.64 | 0.64 | 0.65 | 0.63 | 0.63 | 0.65 | 0.64 | 0.63 | 0.62 | 0.63 |
| 9 | 2 | extratrees | 0.7 | 0.71 | 0.72 | 0.69 | 0.68 | 0.67 | 0.7 | 0.7 | 0.69 | 0.67 |
| 9 | 19 | extratrees | 0.61 | 0.61 | 0.61 | 0.6 | 0.61 | 0.6 | 0.6 | 0.6 | 0.59 | 0.59 |
| 9 | 20 | extratrees | 0.6 | 0.6 | 0.6 | 0.61 | 0.61 | 0.6 | 0.61 | 0.61 | 0.6 | 0.6 |
| 9 | 20 | gini | 0.66 | 0.64 | 0.66 | 0.64 | 0.65 | 0.66 | 0.67 | 0.65 | 0.64 | 0.65 |
| 10 | 3 | extratrees | 0.75 | 0.73 | 0.75 | 0.72 | 0.71 | 0.74 | 0.73 | 0.73 | 0.73 | 0.75 |
| 10 | 4 | extratrees | 0.73 | 0.71 | 0.73 | 0.7 | 0.72 | 0.72 | 0.74 | 0.71 | 0.71 | 0.71 |
| 10 | 12 | extratrees | 0.63 | 0.63 | 0.63 | 0.63 | 0.63 | 0.64 | 0.64 | 0.63 | 0.63 | 0.63 |
| 10 | 16 | extratrees | 0.63 | 0.62 | 0.62 | 0.63 | 0.63 | 0.62 | 0.62 | 0.64 | 0.61 | 0.61 |
| 11 | 8 | gini | 0.73 | 0.71 | 0.71 | 0.73 | 0.72 | 0.72 | 0.73 | 0.73 | 0.72 | 0.73 |
| 11 | 15 | gini | 0.68 | 0.67 | 0.68 | 0.7 | 0.68 | 0.68 | 0.67 | 0.67 | 0.69 | 0.67 |

**Table SM3.4. Randomly drawn hyperparameters combinations and average logloss for XGBoost with CA125 as a predictor.** Missing data for CA125 were addressed using multiple imputation, hence the 10-fold cross-validation procedure was applied in each of the 10 imputed datasets. The best hyperparameter combination is marked in bold.

| | | | | | | | Average logloss for the 10 imputed datasets | | | | | | | | | |
|---|---|---|---|---|---|---|---|---|---|---|---|---|---|---|---|---|
| **eta** | **max_depth** | **gamma** | **ColSample_bytree** | **Min_child_weight** | **SubSample** | **Nrounds** | **1** | **2** | **3** | **4** | **5** | **6** | **7** | **8** | **9** | **10** |
| **0.04** | **3** | **1.68** | **0.59** | **1** | **0.65** | **440** | **0.56** | **0.56** | **0.57** | **0.56** | **0.56** | **0.57** | **0.57** | **0.57** | **0.56** | **0.56** |
| 0.06 | 3 | 2.51 | 0.43 | 5 | 1 | 272 | 0.57 | 0.57 | 0.57 | 0.57 | 0.57 | 0.57 | 0.57 | 0.57 | 0.56 | 0.56 |
| 0.10 | 10 | 2.13 | 0.7 | 5 | 0.83 | 30 | 0.6 | 0.6 | 0.61 | 0.6 | 0.6 | 0.61 | 0.61 | 0.61 | 0.6 | 0.6 |
| 0.16 | 9 | 3.94 | 0.32 | 9 | 0.32 | 484 | 0.57 | 0.57 | 0.57 | 0.57 | 0.57 | 0.57 | 0.57 | 0.57 | 0.56 | 0.56 |
| 0.22 | 6 | 9.78 | 0.4 | 5 | 0.86 | 422 | 0.58 | 0.58 | 0.58 | 0.58 | 0.58 | 0.58 | 0.58 | 0.58 | 0.58 | 0.58 |
| 0.22 | 10 | 0.05 | 0.37 | 2 | 0.44 | 551 | 0.87 | 0.87 | 0.88 | 0.88 | 0.87 | 0.88 | 0.88 | 0.88 | 0.86 | 0.88 |
| 0.25 | 1 | 7.63 | 0.48 | 14 | 0.26 | 832 | 0.58 | 0.58 | 0.58 | 0.58 | 0.58 | 0.58 | 0.58 | 0.58 | 0.58 | 0.58 |
| 0.25 | 2 | 7.3 | 0.6 | 5 | 0.46 | 595 | 0.57 | 0.57 | 0.57 | 0.57 | 0.57 | 0.57 | 0.57 | 0.57 | 0.57 | 0.57 |
| 0.26 | 9 | 7.17 | 0.58 | 17 | 0.94 | 580 | 0.57 | 0.58 | 0.58 | 0.58 | 0.57 | 0.58 | 0.58 | 0.58 | 0.57 | 0.57 |
| 0.28 | 3 | 8.48 | 0.55 | 4 | 0.67 | 79 | 0.58 | 0.58 | 0.58 | 0.58 | 0.58 | 0.58 | 0.58 | 0.58 | 0.58 | 0.58 |
| 0.31 | 4 | 0.56 | 0.44 | 6 | 0.45 | 926 | 0.77 | 0.77 | 0.78 | 0.78 | 0.77 | 0.78 | 0.78 | 0.78 | 0.76 | 0.77 |
| 0.33 | 6 | 3.08 | 0.36 | 0 | 0.29 | 387 | 0.62 | 0.62 | 0.62 | 0.62 | 0.62 | 0.62 | 0.62 | 0.61 | 0.6 | 0.61 |
| 0.34 | 5 | 4.71 | 0.65 | 0 | 0.64 | 347 | 0.57 | 0.57 | 0.57 | 0.57 | 0.57 | 0.57 | 0.58 | 0.57 | 0.57 | 0.57 |
| 0.35 | 5 | 9.02 | 0.66 | 9 | 0.97 | 224 | 0.58 | 0.58 | 0.58 | 0.58 | 0.58 | 0.58 | 0.58 | 0.58 | 0.58 | 0.58 |
| 0.35 | 10 | 1.66 | 0.31 | 6 | 0.84 | 484 | 0.62 | 0.63 | 0.63 | 0.62 | 0.63 | 0.63 | 0.63 | 0.63 | 0.62 | 0.62 |
| 0.38 | 4 | 7.88 | 0.42 | 12 | 0.66 | 862 | 0.57 | 0.57 | 0.57 | 0.57 | 0.57 | 0.57 | 0.57 | 0.57 | 0.57 | 0.57 |
| 0.41 | 10 | 6.47 | 0.64 | 3 | 0.74 | 58 | 0.58 | 0.58 | 0.57 | 0.58 | 0.58 | 0.58 | 0.58 | 0.58 | 0.57 | 0.57 |
| 0.44 | 2 | 5.8 | 0.52 | 7 | 1 | 430 | 0.58 | 0.58 | 0.58 | 0.58 | 0.58 | 0.58 | 0.58 | 0.58 | 0.57 | 0.57 |
| 0.45 | 2 | 7.42 | 0.37 | 8 | 0.26 | 624 | 0.58 | 0.58 | 0.58 | 0.58 | 0.58 | 0.58 | 0.58 | 0.58 | 0.57 | 0.58 |
| 0.46 | 5 | 2.07 | 0.53 | 1 | 0.31 | 523 | 0.74 | 0.73 | 0.74 | 0.73 | 0.72 | 0.73 | 0.74 | 0.74 | 0.72 | 0.74 |
| 0.48 | 8 | 7.64 | 0.54 | 19 | 0.72 | 733 | 0.57 | 0.57 | 0.57 | 0.58 | 0.57 | 0.57 | 0.58 | 0.57 | 0.57 | 0.57 |
| 0.49 | 1 | 8.73 | 0.46 | 4 | 0.96 | 181 | 0.59 | 0.59 | 0.59 | 0.59 | 0.59 | 0.59 | 0.59 | 0.59 | 0.59 | 0.59 |
| 0.52 | 2 | 4.8 | 0.34 | 11 | 0.62 | 284 | 0.57 | 0.57 | 0.57 | 0.57 | 0.57 | 0.57 | 0.57 | 0.57 | 0.57 | 0.57 |
| 0.52 | 8 | 6.25 | 0.6 | 7 | 0.65 | 231 | 0.58 | 0.58 | 0.58 | 0.58 | 0.58 | 0.58 | 0.58 | 0.57 | 0.57 | 0.57 |
| 0.53 | 4 | 0.15 | 0.64 | 2 | 0.71 | 359 | 0.88 | 0.88 | 0.89 | 0.88 | 0.88 | 0.89 | 0.9 | 0.88 | 0.87 | 0.89 |
| 0.53 | 6 | 5.88 | 0.43 | 4 | 0.66 | 764 | 0.58 | 0.58 | 0.58 | 0.58 | 0.58 | 0.58 | 0.58 | 0.58 | 0.58 | 0.58 |
| 0.54 | 9 | 2.62 | 0.68 | 4 | 0.76 | 573 | 0.64 | 0.64 | 0.65 | 0.64 | 0.64 | 0.64 | 0.64 | 0.64 | 0.64 | 0.64 |
| 0.55 | 7 | 3.9 | 0.31 | 18 | 0.66 | 81 | 0.57 | 0.57 | 0.57 | 0.57 | 0.57 | 0.57 | 0.57 | 0.57 | 0.57 | 0.57 |
| 0.56 | 9 | 6.53 | 0.33 | 20 | 0.46 | 873 | 0.57 | 0.57 | 0.57 | 0.57 | 0.57 | 0.57 | 0.57 | 0.57 | 0.57 | 0.57 |
| 0.59 | 8 | 6.46 | 0.7 | 19 | 0.65 | 119 | 0.57 | 0.57 | 0.58 | 0.58 | 0.58 | 0.57 | 0.58 | 0.58 | 0.57 | 0.57 |

**Table SM3.5. Randomly drawn hyperparameter combinations and average logloss for the neural network with CA125 as a predictor.** Missing data for CA125 were addressed using multiple imputation, hence the 10-fold cross-validation procedure was applied in each of the 10 imputed datasets. The best hyperparameter combination is marked in bold.

| | | **Average logloss for the 10 imputed datasets** | | | | | | | | | |
|---|---|---|---|---|---|---|---|---|---|---|---|
| **Size** | **Decay** | **1** | **2** | **3** | **4** | **5** | **6** | **7** | **8** | **9** | **10** |
| 1 | 0.004 | 0.72 | 0.68 | 0.72 | 0.68 | 0.72 | 0.75 | 0.68 | 0.71 | 0.67 | 0.68 |
| 2 | 0.45 | 0.61 | 0.61 | 0.61 | 0.61 | 0.61 | 0.61 | 0.61 | 0.61 | 0.6 | 0.61 |
| 3 | 0.001 | 0.61 | 0.6 | 0.61 | 0.61 | 0.59 | 0.6 | 0.6 | 0.61 | 0.59 | 0.59 |
| 3 | 3.32 | 0.61 | 0.6 | 0.61 | 0.61 | 0.61 | 0.61 | 0.61 | 0.6 | 0.6 | 0.6 |
| 4 | 0.02 | 0.58 | 0.58 | 0.58 | 0.59 | 0.58 | 0.58 | 0.58 | 0.58 | 0.58 | 0.57 |
| 5 | 0.67 | 0.58 | 0.58 | 0.57 | 0.57 | 0.58 | 0.58 | 0.58 | 0.58 | 0.57 | 0.57 |
| 6 | 0.17 | 0.57 | 0.57 | 0.57 | 0.57 | 0.57 | 0.57 | 0.57 | 0.58 | 0.57 | 0.57 |
| 6 | 0.19 | 0.57 | 0.57 | 0.57 | 0.57 | 0.57 | 0.57 | 0.57 | 0.57 | 0.57 | 0.56 |
| 7 | 0.0004 | 0.65 | 0.62 | 0.65 | 0.65 | 0.62 | 0.67 | 0.63 | 0.64 | 0.64 | 0.63 |
| 8 | 0.002 | 0.61 | 0.61 | 0.61 | 0.6 | 0.61 | 0.62 | 0.62 | 0.61 | 0.6 | 0.6 |
| 10 | 0.01 | 0.6 | 0.6 | 0.61 | 0.61 | 0.6 | 0.61 | 0.6 | 0.62 | 0.61 | 0.59 |
| 10 | 3.72 | 0.57 | 0.57 | 0.57 | 0.57 | 0.57 | 0.57 | 0.57 | 0.57 | 0.57 | 0.57 |
| 11 | 0.001 | 0.68 | 0.69 | 0.67 | 0.73 | 0.66 | 0.67 | 0.68 | 0.67 | 0.68 | 0.68 |
| 11 | 0.004 | 0.63 | 0.61 | 0.64 | 0.63 | 0.63 | 0.64 | 0.64 | 0.62 | 0.62 | 0.63 |
| 12 | 0.0002 | 0.73 | 0.75 | 0.75 | 0.76 | 0.73 | 0.72 | 0.68 | 0.71 | 0.69 | 0.73 |
| 12 | 0.001 | 0.71 | 0.72 | 0.77 | 0.76 | 0.7 | 0.77 | 0.72 | 0.69 | 0.71 | 0.71 |
| **12** | **2.04** | **0.56** | **0.56** | **0.56** | **0.56** | **0.56** | **0.56** | **0.56** | **0.56** | **0.56** | **0.56** |
| 15 | 0.0004 | 0.7 | 0.73 | 0.75 | 0.74 | 0.7 | 0.77 | 0.7 | 0.7 | 0.73 | 0.76 |
| 15 | 0.003 | 0.67 | 0.68 | 0.68 | 0.7 | 0.66 | 0.68 | 0.67 | 0.68 | 0.67 | 0.7 |
| 15 | 0.24 | 0.58 | 0.57 | 0.57 | 0.57 | 0.58 | 0.58 | 0.58 | 0.57 | 0.57 | 0.57 |
| 16 | 0.0001 | 0.79 | 0.79 | 0.83 | 0.85 | 0.82 | 0.8 | 0.79 | 0.78 | 0.77 | 0.73 |
| 16 | 0.003 | 0.68 | 0.7 | 0.66 | 0.68 | 0.67 | 0.68 | 0.69 | 0.68 | 0.67 | 0.67 |
| 16 | 0.3 | 0.58 | 0.58 | 0.57 | 0.57 | 0.57 | 0.58 | 0.58 | 0.57 | 0.57 | 0.57 |
| 16 | 0.32 | 0.57 | 0.57 | 0.57 | 0.57 | 0.57 | 0.58 | 0.57 | 0.57 | 0.57 | 0.57 |
| 17 | 0.1 | 0.6 | 0.59 | 0.59 | 0.6 | 0.58 | 0.6 | 0.59 | 0.6 | 0.59 | 0.59 |
| 18 | 0.0002 | 0.75 | 0.83 | 0.82 | 0.76 | 0.81 | 0.83 | 0.79 | 0.86 | 0.78 | 0.82 |
| 18 | 0.001 | 0.75 | 0.79 | 0.78 | 0.79 | 0.72 | 0.76 | 0.75 | 0.79 | 0.79 | 0.74 |
| 18 | 0.38 | 0.57 | 0.58 | 0.57 | 0.57 | 0.57 | 0.57 | 0.57 | 0.57 | 0.57 | 0.57 |
| 20 | 0.001 | 0.98 | 0.82 | 0.97 | 0.94 | 0.88 | 0.92 | 0.82 | 0.88 | 0.84 | 1.02 |
| 20 | 0.002 | 0.77 | 0.75 | 0.76 | 0.76 | 0.77 | 0.77 | 0.73 | 0.75 | 0.74 | 0.74 |

**Table SM3.6. Randomly drawn hyperparameter combinations and average logloss for SVM with CA125 as a predictor.** Missing data for CA125 were addressed using multiple imputation, hence the 10-fold cross-validation procedure was applied in each of the 10 imputed datasets. The best hyperparameter combination is marked in bold.

| | | **Average logloss for the 10 imputed datasets** | | | | | | | | | |
|---|---|---|---|---|---|---|---|---|---|---|---|
| **sigma** | **C** | **1** | **2** | **3** | **4** | **5** | **6** | **7** | **8** | **9** | **10** |
| 0.002 | 0.34 | 0.73 | 0.73 | 0.73 | 0.73 | 0.73 | 0.73 | 0.73 | 0.73 | 0.73 | 0.73 |
| 0.003 | 0.05 | 0.75 | 0.75 | 0.75 | 0.75 | 0.75 | 0.75 | 0.75 | 0.75 | 0.75 | 0.75 |
| **0.004** | **4.41** | **0.69** | **0.7** | **0.7** | **0.69** | **0.69** | **0.69** | **0.69** | **0.69** | **0.69** | **0.69** |
| 0.01 | 0.36 | 0.71 | 0.71 | 0.71 | 0.71 | 0.71 | 0.71 | 0.71 | 0.71 | 0.71 | 0.71 |
| 0.01 | 1.06 | 0.71 | 0.71 | 0.71 | 0.71 | 0.71 | 0.71 | 0.7 | 0.71 | 0.7 | 0.7 |
| 0.01 | 2.42 | 0.7 | 0.7 | 0.7 | 0.7 | 0.7 | 0.7 | 0.7 | 0.7 | 0.7 | 0.7 |
| 0.01 | 0.03 | 0.75 | 0.76 | 0.76 | 0.75 | 0.75 | 0.75 | 0.75 | 0.75 | 0.75 | 0.75 |
| 0.02 | 156.38 | 0.72 | 0.72 | 0.72 | 0.72 | 0.72 | 0.72 | 0.72 | 0.72 | 0.72 | 0.72 |
| 0.03 | 24.38 | 0.71 | 0.71 | 0.71 | 0.71 | 0.71 | 0.71 | 0.71 | 0.71 | 0.71 | 0.71 |
| 0.05 | 0.86 | 0.73 | 0.73 | 0.73 | 0.73 | 0.73 | 0.73 | 0.73 | 0.73 | 0.72 | 0.72 |
| 0.05 | 6.24 | 0.72 | 0.72 | 0.72 | 0.72 | 0.71 | 0.72 | 0.71 | 0.72 | 0.71 | 0.71 |
| 0.06 | 14.95 | 0.72 | 0.72 | 0.72 | 0.72 | 0.72 | 0.72 | 0.72 | 0.72 | 0.71 | 0.71 |
| 0.08 | 20.38 | 0.72 | 0.72 | 0.73 | 0.73 | 0.72 | 0.72 | 0.72 | 0.72 | 0.72 | 0.72 |
| 0.09 | 71.38 | 0.74 | 0.74 | 0.75 | 0.74 | 0.74 | 0.74 | 0.74 | 0.74 | 0.74 | 0.74 |
| 0.1 | 0.05 | 0.78 | 0.78 | 0.78 | 0.78 | 0.78 | 0.78 | 0.78 | 0.78 | 0.78 | 0.78 |
| 0.11 | 29.63 | 0.73 | 0.73 | 0.74 | 0.74 | 0.73 | 0.73 | 0.73 | 0.73 | 0.73 | 0.73 |
| 0.13 | 0.05 | 0.79 | 0.79 | 0.79 | 0.79 | 0.79 | 0.79 | 0.79 | 0.79 | 0.79 | 0.79 |
| 0.13 | 86.24 | 0.75 | 0.76 | 0.77 | 0.77 | 0.76 | 0.76 | 0.76 | 0.76 | 0.76 | 0.76 |
| 0.15 | 759.25 | 0.82 | 0.82 | 0.83 | 0.83 | 0.81 | 0.82 | 0.82 | 0.83 | 0.81 | 0.81 |
| 0.16 | 127.85 | 0.77 | 0.78 | 0.79 | 0.78 | 0.78 | 0.78 | 0.78 | 0.79 | 0.77 | 0.78 |
| 0.19 | 0.81 | 0.74 | 0.74 | 0.74 | 0.74 | 0.74 | 0.74 | 0.74 | 0.74 | 0.74 | 0.74 |
| 0.2 | 3.25 | 0.73 | 0.73 | 0.73 | 0.73 | 0.73 | 0.73 | 0.73 | 0.73 | 0.73 | 0.73 |
| 0.23 | 11.05 | 0.74 | 0.74 | 0.74 | 0.75 | 0.74 | 0.74 | 0.74 | 0.74 | 0.73 | 0.74 |
| 0.39 | 0.17 | 0.77 | 0.78 | 0.78 | 0.78 | 0.78 | 0.77 | 0.78 | 0.78 | 0.77 | 0.78 |
| 0.44 | 0.04 | 0.8 | 0.81 | 0.81 | 0.81 | 0.81 | 0.8 | 0.8 | 0.8 | 0.8 | 0.8 |
| 0.66 | 301.52 | 0.81 | 0.81 | 0.81 | 0.82 | 0.81 | 0.82 | 0.82 | 0.82 | 0.81 | 0.81 |
| 0.79 | 218.64 | 0.81 | 0.81 | 0.81 | 0.82 | 0.81 | 0.82 | 0.82 | 0.82 | 0.81 | 0.81 |
| 0.96 | 179.12 | 0.81 | 0.81 | 0.82 | 0.82 | 0.82 | 0.82 | 0.82 | 0.82 | 0.81 | 0.81 |
| 1.02 | 4.66 | 0.77 | 0.77 | 0.77 | 0.77 | 0.77 | 0.77 | 0.77 | 0.77 | 0.76 | 0.77 |
| 1.06 | 20.42 | 0.79 | 0.79 | 0.8 | 0.79 | 0.79 | 0.8 | 0.8 | 0.8 | 0.79 | 0.79 |

**Table SM3.7. Grid search parameter and average logloss for ridge MLR without CA125.** The best hyperparameter combination is marked in bold.

| Lambda | Average logloss |
|---|---|
| 0.00099 | 0.66 |
| 0.00117 | 0.66 |
| 0.00178 | 0.66 |
| 0.00340 | 0.66 |
| 0.00549 | 0.66 |
| 0.01265 | 0.66 |
| **0.01641** | **0.66** |
| 0.02860 | 0.66 |
| 0.0520 | 0.67 |
| 0.08061 | 0.68 |
| 0.10881 | 0.69 |
| 0.13585 | 0.70 |
| 0.14604 | 0.70 |
| 0.18028 | 0.71 |
| 0.18647 | 0.72 |
| 0.18671 | 0.72 |
| 0.25463 | 0.73 |
| 0.32586 | 0.75 |
| 0.52 | 0.78 |
| 0.6282 | 0.80 |
| 0.76854 | 0.81 |
| 0.80835 | 0.82 |
| 1.06 | 0.84 |
| 1.12 | 0.85 |
| 1.87 | 0.89 |
| 2.37 | 0.91 |
| 2.52 | 0.91 |
| 2.78 | 0.92 |
| 4.17 | 0.95 |
| 4.23 | 0.95 |

**Table SM3.8. Randomly drawn hyperparameter combinations and average logloss for the random forest without CA125.** The best hyperparameter combination is marked in bold.

| mtry | min.node.size | splitrule | Average logloss |
|---|---|---|---|
| 1 | 16 | extratrees | 0.79 |
| 1 | 4 | gini | 0.76 |
| 1 | 14 | gini | 0.76 |
| 1 | 15 | gini | 0.76 |
| 2 | 9 | extratrees | 0.67 |
| 3 | 5 | extratrees | 0.63 |
| 3 | 4 | gini | 0.63 |
| 3 | 7 | gini | 0.63 |
| **3** | **15** | **gini** | **0.62** |
| 5 | 5 | extratrees | 0.64 |
| 6 | 4 | extratrees | 0.65 |
| 6 | 8 | extratrees | 0.64 |
| 6 | 3 | gini | 0.70 |
| 6 | 6 | gini | 0.68 |
| 7 | 12 | gini | 0.69 |
| 8 | 2 | extratrees | 0.73 |
| 8 | 9 | extratrees | 0.66 |
| 8 | 5 | gini | 0.79 |
| 8 | 14 | gini | 0.73 |
| 8 | 20 | gini | 0.70 |
| 9 | 2 | extratrees | 0.79 |
| 9 | 19 | extratrees | 0.65 |
| 9 | 20 | extratrees | 0.65 |
| 9 | 20 | gini | 0.74 |
| 10 | 3 | extratrees | 0.91 |
| 10 | 4 | extratrees | 0.91 |
| 10 | 12 | extratrees | 0.73 |
| 10 | 16 | extratrees | 0.70 |
| 11 | 8 | gini | 0.86 |
| 11 | 15 | gini | 0.77 |

**Table SM3.9. Randomly drawn hyperparameters combinations and average logloss for XGBoost without CA125.** The best hyperparameter combination is marked in bold.

| eta | max_depth | gamma | ColSample_ by tree | Min_child_weight | SubSample | Nrounds | Average logloss |
|---|---|---|---|---|---|---|---|
| **0.04** | **3** | **1.68** | **0.59** | **1** | **0.65** | **440** | **0.615** |
| 0.06 | 3 | 2.51 | 0.43 | 5 | 1 | 272 | 0.618 |
| 0.10 | 10 | 2.13 | 0.70 | 5 | 0.83 | 30 | 0.651 |
| 0.16 | 9 | 3.9 | 0.32 | 9 | 0.32 | 484 | 0.617 |
| 0.22 | 6 | 9.78 | 0.40 | 5 | 0.86 | 422 | 0.634 |
| 0.22 | 10 | 0.05 | 0.37 | 2 | 0.44 | 551 | 0.917 |
| 0.25 | 1 | 7.63 | 0.48 | 14 | 0.26 | 832 | 0.635 |
| 0.25 | 2 | 7.3 | 0.60 | 5 | 0.46 | 595 | 0.624 |
| 0.26 | 9 | 7.17 | 0.58 | 17 | 0.94 | 580 | 0.627 |
| 0.28 | 3 | 8.48 | 0.55 | 4 | 0.67 | 79 | 0.632 |
| 0.31 | 4 | 0.56 | 0.44 | 6 | 0.45 | 926 | 0.797 |
| 0.33 | 6 | 3.08 | 0.36 | 0 | 0.29 | 387 | 0.649 |
| 0.34 | 5 | 4.71 | 0.65 | 0 | 0.64 | 347 | 0.621 |
| 0.35 | 5 | 9.02 | 0.66 | 9 | 0.97 | 224 | 0.633 |
| 0.35 | 10 | 1.66 | 0.31 | 6 | 0.84 | 484 | 0.661 |
| 0.38 | 4 | 7.88 | 0.42 | 12 | 0.66 | 862 | 0.623 |
| 0.41 | 10 | 6.47 | 0.64 | 3 | 0.74 | 58 | 0.625 |
| 0.44 | 2 | 5.80 | 0.52 | 7 | 1 | 430 | 0.626 |
| 0.45 | 2 | 7.42 | 0.37 | 8 | 0.26 | 624 | 0.629 |
| 0.46 | 5 | 2.07 | 0.53 | 1 | 0.31 | 523 | 0.768 |
| 0.48 | 8 | 7.64 | 0.54 | 19 | 0.72 | 733 | 0.625 |
| 0.49 | 1 | 8.73 | 0.46 | 4 | 0.96 | 181 | 0.643 |
| 0.52 | 2 | 4.80 | 0.34 | 11 | 0.62 | 284 | 0.618 |
| 0.52 | 8 | 6.25 | 0.60 | 7 | 0.65 | 231 | 0.622 |
| 0.53 | 4 | 0.15 | 0.64 | 2 | 0.71 | 359 | 0.925 |
| 0.53 | 6 | 5.88 | 0.43 | 4 | 0.66 | 764 | 0.626 |
| 0.54 | 9 | 2.62 | 0.68 | 4 | 0.76 | 573 | 0.677 |
| 0.55 | 7 | 3.90 | 0.31 | 18 | 0.66 | 81 | 0.619 |
| 0.56 | 9 | 6.53 | 0.33 | 20 | 0.46 | 873 | 0.623 |
| 0.59 | 8 | 6.46 | 0.7 | 19 | 0.65 | 119 | 0.625 |

**Table SM3.10. Randomly drawn hyperparameters combinations and average logloss for the neural network without CA125.** The best hyperparameter combination is marked in bold.

| Size | Decay | Average logloss |
|---|---|---|
| 1 | 0.004 | 0.67 |
| 2 | 0.45 | 0.65 |
| 3 | 0.001 | 0.63 |
| 3 | 3.32 | 0.64 |
| 4 | 0.02 | 0.61 |
| 5 | 0.67 | 0.61 |
| 6 | 0.17 | 0.62 |
| 6 | 0.19 | 0.62 |
| 7 | 0.00004 | 0.67 |
| 8 | 0.002 | 0.64 |
| 10 | 0.01 | 0.63 |
| 10 | 3.72 | 0.62 |
| 11 | 0.0006 | 0.65 |
| 11 | 0.004 | 0.65 |
| 12 | 0.00002 | 0.77 |
| 12 | 0.00007 | 0.72 |
| **12** | **2.04** | **0.61** |
| 15 | 0.0004 | 0.70 |
| 15 | 0.003 | 0.70 |
| 15 | 0.24 | 0.62 |
| 16 | 0.0001 | 0.74 |
| 16 | 0.003 | 0.68 |
| 16 | 0.30 | 0.63 |
| 16 | 0.32 | 0.62 |
| 17 | 0.10 | 0.65 |
| 18 | 0.0002 | 0.79 |
| 18 | 0.0005 | 0.76 |
| 18 | 0.38 | 0.62 |
| 20 | 0.00006 | 0.79 |
| 20 | 0.002 | 0.73 |

**Table SM3.11. Randomly drawn hyperparameters combinations and average logloss for SVM without CA125.** The best hyperparameter combination is marked in bold.

| sigma | C | logloss |
|---|---|---|
| 0.002 | 0.34 | 0.74 |
| 0.003 | 0.05 | 0.76 |
| **0.004** | **4.41** | **0.70** |
| 0.01 | 0.36 | 0.72 |
| 0.01 | 1.06 | 0.71 |
| 0.01 | 2.42 | 0.71 |
| 0.01 | 0.03 | 0.76 |
| 0.02 | 156.38 | 0.74 |
| 0.03 | 24.38 | 0.73 |
| 0.05 | 0.86 | 0.74 |
| 0.05 | 6.24 | 0.73 |
| 0.06 | 14.95 | 0.74 |
| 0.08 | 20.38 | 0.76 |
| 0.09 | 71.38 | 0.77 |
| 0.1 | 0.05 | 0.78 |
| 0.11 | 29.63 | 0.76 |
| 0.13 | 0.05 | 0.79 |
| 0.13 | 86.24 | 0.79 |
| 0.15 | 759.25 | 0.84 |
| 0.16 | 127.85 | 0.80 |
| 0.19 | 0.81 | 0.75 |
| 0.2 | 3.25 | 0.75 |
| 0.23 | 11.05 | 0.76 |
| 0.39 | 0.17 | 0.78 |
| 0.44 | 0.04 | 0.81 |
| 0.66 | 301.52 | 0.83 |
| 0.79 | 218.64 | 0.84 |
| 0.96 | 179.12 | 0.84 |
| 1.02 | 4.66 | 0.78 |
| 1.06 | 20.42 | 0.81 |

**Supplementary Material 4. Multiple imputation for CA125**

For the current study, we used the imputations that were obtained for the original studies.[1,7] CA125 was missing in 1805/5909 (31%) patients in the development data, and in 966/3199 (30%) patients in the external validation data. Missing values are probably caused by (1) varying local protocols about CA125 measurement and (2) less clinical need to measure CA125 when the ultrasound evaluation was suggestive of a benign tumor (Table SM4.1). As a result, missing values were more common in women with a benign tumor. The missingness mechanism is most likely Missing At Random (MAR), meaning that missing values are random conditional on other observed information (e.g. clinical and ultrasound information, and the tumor histology and stage of malignant tumors).

**Table SM4.1. Missing values for CA125 by subjective assessment by the ultrasound examiner.**

| Subjective assessment by ultrasound examiner | Missing values for CA125, n (%) | |
|---|---|---|
| | Development | Validation |
| Certainly benign | 1000/2591 (39%) | 453/1068 (42%) |
| Probably benign | 311/992 (31%) | 235/657 (36%) |
| Uncertain | 107/412 (26%) | 80/323 (25%) |
| Probably malignant | 144/623 (23%) | 100/463 (22%) |
| Certainly malignant | 243/1291 (19%) | 98/688 (14%) |
| All | 1805/5909 (31%) | 966/3199 (30%) |

The missing values were handled using multiple imputation. The imputation was performed separately for the development and validation data.[1,7] 100 imputed development and validation datasets were available, but we used only the first 10 datasets for the current study. We considered multiple imputation a more appropriate method than complete case analysis.[16] Because missing values are not purely random, a complete case analysis fundamentally changes the target population, which can affect model performance and generalizability. In addition, it would lead to a substantial loss of data.

1. Development dataset

In the development data, predictive mean matching regression was used based on variables that were related to CA125 or to its missingness. Log(log(CA125+1)) was imputed due to the heavy skewness of CA125 values. Variables used to impute CA125 were center (nominal), patient age (in years), personal history of ovarian cancer (yes/no), current use of hormonal therapy (yes/no), hysterectomy (yes/no), parity (count), pelvic pain during examination (yes/no), maximum diameter of the lesion (in mm), color score of intra-tumoral blood flow (ordinal: 1-4), venous blood flow only (yes/no), presence of an incomplete septum (yes/no), irregular internal cyst walls (yes/no), hemorrhagic echogenicity (yes/no), ascites (yes/no), bilaterality (yes/no), shadows (yes/no), presence of locules (yes/no), the number of locules (1, 2-10, >10, none), the presence of solid components (yes/no), the maximal diameter of the largest solid component (in mm), amount of fluid in the pouch of Douglas (in mm), pathology group (nominal with 21 levels), and log(log(CA125+1)). This was done using the regpmm function of the PROC MI procedure in SAS v9.3. Two comments are of interest. First, in the model development data, imputation was done separately for the data from IOTA phases 1, 1b, and 2 (n=3506) and for the data from IOTA phase 3 (n=2403). Second, the predictor selection procedure was conducted on 5 of the imputed datasets from the first part of the data (n=3506), with minimal differences.[1]

2. External validation dataset

The external validation data are part of the IOTA5 interim analysis dataset, which includes patients with a new mass that were operated or followed conservatively.[7] For the current study, we only use patients who were operated within 120 days following the ultrasound examination without follow-up scan. In this group, histology was missing for 8 patients. For another 15 patients, it was known that the tumor was malignant but the type of malignancy was unclear.

Multiple imputation with chained equations (mice) was used to address missing values of CA125 and the multinomial reference standard (benign, borderline, stage I primary malignancy, stage II-IV primary malignancy and secondary metastatic malignancy). Predictive mean matching regression was used to impute log(log(CA125 + 1)). Multinomial logistic regression was used to impute the multinomial outcome. Imputations were based on variables that are likely related to CA125/outcome or to the unavailability of CA125/outcome, and/or variables used in the ADNEX model. Hence, the following variables were used: patient age (in years), type of center (binary), log of the maximum diameter of the lesion (in mm), proportion of solid tissue (calculated as the maximum diameter of the largest solid component divided by the maximum diameter of the lesion), the square of the proportion of solid tissue, the number of locules (1, 2-10, >10, none), the number of papillations (ordinal variable: 0, 1, 2, 3, >3), acoustic shadows (yes/no), ascites (yes/no), presence of metastases (yes/no), bilaterality (yes/no), pelvic pain during examination (yes/no), personal history of ovarian cancer (yes/no), irregular internal cyst walls (yes/no), maximum papillary height (in mm), presence of papillary projections with blood flow (yes/no), color score of intra-tumoral blood flow (ordinal variable: 1-4), echogenicity of cyst fluid (nominal: anechoic, low-level, ground glass, hemorrhagic, mixed, no cyst fluid), log(log(CA125+1)), presumed endometrioma (yes/no), subjective assessment at inclusion (ordinal: certainly benign, probably benign, benign but uncertain, malignant but uncertain, probably malignant, certainly malignant), and the multinomial outcome. All variables (except the outcome) were based on the inclusion scan.

**Supplementary Material 5. Flowcharts for modeling and validation procedure**

The detailed flowchart for model development and validation when multiple imputation was used (i.e. models with CA125 as a predictor) was as follows:

1. Using a specific algorithm (e.g., Random Forest), fit a model in each imputed development dataset (IDD), yielding 10 models:
   a. In each IDD, 10-fold cross-validation is performed to tune hyperparameters (if any).
   b. Using the selected hyperparameters for that IDD, a model is fitted using the complete IDD.
2. Calculate external validation performance:
   a. Apply the 10 developed models on each of the 10 imputed validation datasets (IVD); for each validation patient, this yields 100 risk estimates per outcome category.
   b. Average the predictions of the 10 models within each IVD; for each patient, this reduces the 100 risk estimates per outcome category to 10 risk estimates; i.e. 1 per IVD.
   c. Calculate center-specific performance metrics for each IVD:
      i. Polytomous discrimination index (PDI), pairwise area under the curves (AUROCs), binary AUROC: center-specific estimates are obtained.
      ii. Flexible calibration curves for each of the five outcome categories: center-specific loess-based calibration models are obtained using the probability of each outcome category (i.e. 5 curves per center per IVD).
      iii. Clinical utility: Calculate center-specific net benefit, sensitivity, specificity and prevalence at each pre-specified risk threshold.
      iv. Note: centers with small numbers were grouped into a single 'other' group. For PDI, pairwise AUROCs, and flexible calibration curves, 15 centers with <7 cases for at least one outcome category were grouped, leaving 10 centers and 1 other group. For the binary AUROC and clinical utility, 6 centers with < 10 malignancies were grouped, leaving 19 centers and 1 other group.
   d. Combine the center-specific performance estimates across the IVDs to get final estimates:
      i. PDI, pairwise AUROCs, binary AUROC: center-specific estimates per IVD are combined using Rubin's rules. Then, random effects meta-analysis is used to combine center-specific estimates.
      ii. Flexible calibration curves: per IVD, the center-specific calibration models are applied to all patients in the IVD. Per patient, the estimated probabilities from the prediction model and the fitted values from the center-specific calibration models are averaged over the IVD. Then, we averaged the resulting fitted values from each center-specific calibration model per patient, using the square root of the center-specific sample size as weights. The averaged estimated probabilities and the weighted average of the fitted values from the calibration models are used to plot the flexible calibration curves per outcome.
      iii. Clinical utility: the center-specific net benefit, sensitivity and specificity are averaged over the IVD, resulting in overall center-specific net benefit, sensitivity and specificity. The software Winbugs is used to perform a Bayesian trivariate random-effects meta-analysis of sensitivity, specificity and prevalence to obtain an overall net benefit per threshold.
3. Repeat for every algorithm.

When no imputation was needed (i.e. models without CA125 as a predictor), the flowchart was the following:

1. Using a specific algorithm (e.g., Random Forest), fit a model in the development dataset:
    a. 10-fold cross-validation is performed to tune hyperparameters (if any).
    b. Using the selected hyperparameters, a model is fitted using the complete development dataset
2. Calculate external validation performance:
    a. Apply the developed model to the external validation dataset.
    b. Calculate the center-specific performance metrics:
        i. PDI, pairwise AUROCs, binary AUROC: center-specific estimates are obtained.
        ii. Flexible calibration curves for each of the five outcome categories: center-specific loess-based calibration models are obtained using the probability of each outcome category (i.e. 5 curves per center).
        iii. Clinical utility: Calculate center-specific net benefit, sensitivity, specificity and prevalence at each pre-specified risk threshold.
        iv. Note: centers with small numbers were grouped into a single 'other' group. For PDI, pairwise AUROCs, and flexible calibration curves, 15 centers with fewer than 7 cases for at least one outcome category were grouped, leaving 10 centers and 1 other group. For the binary AUROC and clinical utility, 6 centers with $< 10$ malignancies were grouped, leaving 19 centers and 1 other group.
    c. Combine the center-specific performance estimates to get final estimates:
        i. PDI, pairwise AUROCs, binary AUROC: random effects meta-analysis is used to combine center-specific estimates.
        ii. Flexible calibration curves: we averaged the fitted values from each center-specific calibration model per patient, using the square root of the center-specific sample size as weights. The estimated probability from the prediction model and the weighted average of the fitted values from the calibration models are used to plot the flexible calibration curves per outcome.
        iii. Clinical utility: the software Winbugs is used to perform a Bayesian trivariate random-effects meta-analysis of sensitivity, specificity and prevalence to obtain an overall net benefit per threshold.
3. Repeat this for every algorithm.

**Table S1. Descriptive statistics by reference standard (final diagnosis).**

| Variables | Tumor outcome<br>Benign | Borderline | Stage I primary invasive | Stage II-IV primary invasive | Secondary metastasis |
|---|---|---|---|---|---|
| Development data | | | | | |
| N | 3980 | 339 | 356 | 988 | 246 |
| Age (years) | 42 (32-54) | 49 (36-62) | 54 (44-64) | 59 (50-67) | 57 (47-68) |
| Serum CA125 (U/mL) | 18 (11-39) | 30 (16-88) | 51 (20-199) | 436 (147-1217) | 89 (30-270) |
| Missing CA125 result | 1447 (36%) | 62 (18%) | 71 (20%) | 163 (16%) | 62 (25%) |
| Max. diameter of lesion (mm) | 63 (45-87) | 86 (52-150) | 106 (71-153) | 85 (56-123) | 86 (56-124) |
| Proportion of solid tissue | 0 (0-0.20) | 0.30 (0.08-0.54) | 0.58 (0.30-1) | 1 (0.55-1) | 1 (0.59-1) |
| Papillary projections | | | | | |
| 0 | 3424 (86%) | 135 (40%) | 227 (64%) | 772 (78%) | 213 (87%) |
| 1 | 333 (8%) | 69 (20%) | 25 (7%) | 56 (6%) | 12 (5%) |
| 2 | 80 (2%) | 21 (6%) | 17 (5%) | 30 (3%) | 0 (0%) |
| 3 | 66 (2%) | 24 (7%) | 17 (5%) | 28 (3%) | 2 (1%) |
| >3 | 77 (2%) | 90 (27%) | 70 (20%) | 102 (10%) | 19 (8%) |
| >10 cyst locules | 199 (5%) | 74 (22%) | 69 (19%) | 93 (9%) | 36 (15%) |
| Ascites | 64 (2%) | 28 (8%) | 65 (18%) | 473 (48%) | 90 (37%) |
| Shadows | 676 (17%) | 8 (2.4%) | 18 (5%) | 30 (3%) | 10 (4%) |
| Validation data | | | | | |
| N | 1988 | 259 | 219 | 561 | 172 |
| Age (years) | 44 (32-58) | 47 (35-60) | 54 (44-64) | 60 (50-69) | 55 (46-67) |
| Serum CA125 (U/mL) | 16 (9-33) | 32 (14-104) | 52 (19-229) | 389 (107-1157) | 78 (22-251) |
| Missing CA125 result | 764 (38%) | 50 (19%) | 41 (19%) | 76 (14%) | 35 (20%) |
| Max. diameter of lesion (mm) | 65 (47-91) | 79 (48-144) | 100 (64-140) | 86 (61-118) | 82 (56-120) |
| Proportion of solid tissue | 0 (0-0.14) | 0.26 (0-0.50) | 0.50 (0.36-0.83) | 0.80 (0.52-1) | 1 (0.44-1) |
| Papillary projections | | | | | |
| 0 | 1798 (90%) | 137 (53%) | 146 (67%) | 476 (85%) | 154 (90%) |
| 1 | 104 (5%) | 37 (14%) | 26 (12%) | 29 (5%) | 4 (2%) |
| 2 | 36 (2%) | 17 (7%) | 9 (4%) | 10 (2%) | 4 (2%) |
| 3 | 18 (1%) | 19 (7%) | 6 (3%) | 8 (1%) | 0 (0%) |
| >3 | 32 (2%) | 49 (19%) | 32 (15%) | 38 (7%) | 10 (6%) |
| >10 cyst locules | 158 (8%) | 63 (24%) | 48 (22%) | 71 (13%) | 34 (20%) |
| Ascites | 21 (1%) | 16 (6%) | 22 (10%) | 206 (37%) | 56 (33%) |
| Shadows | 414 (21%) | 16 (6%) | 12 (5%) | 15 (3%) | 7 (4%) |

Results are shown as median (interquartile range) for continuous variables, and as n (%) for categorical variables.

**Table S2. List of centers in the development and validation data.**

| Center | Country | N | Final diagnosis (reference standard) | | | | |
|---|---|---|---|---|---|---|---|
| | | | **Ben** | **Bot** | **S1** | **S2-4** | **Met** |
| | | DEVELOPMENT DATA | | | | | |
| *University Hospitals Leuven* | *Belgium* | *930* | *596* | *64* | *48* | *171* | *51* |
| *Universita Cattolica del Sacro Cuore, Rome* | *Italy* | *787* | *377* | *44* | *79* | *213* | *74* |
| Skåne University Hospital Malmö | Sweden | 776 | 608 | 35 | 38 | 77 | 18 |
| Ziekenhuis Oost-Limburg, Genk | Belgium | 428 | 367 | 14 | 17 | 28 | 2 |
| *Ospedale San Gerardo, Monza* | *Italy* | *401* | *308* | *30* | *17* | *40* | *6* |
| *General Faculty Hospital, Prague* | *Czech Rep.* | *354* | *120* | *46* | *31* | *133* | *24* |
| *Istituto Europeo di Oncologia, Milan* | *Italy* | *311* | *135* | *21* | *27* | *109* | *19* |
| *Medical University Lublin* | *Poland* | *285* | *183* | *8* | *25* | *61* | *8* |
| Ospedale San Giovanni di Dio, Cagliari | Italy | 261 | 224 | 8 | 8 | 13 | 8 |
| DCS Sacco University of Milan | Italy | 223 | 195 | 4 | 8 | 13 | 3 |
| *University of Bologna* | *Italy* | *213* | *148* | *19* | *10* | *31* | *5* |
| University of Bologna | Italy | 135 | 124 | 3 | 3 | 3 | 2 |
| *Karolinska University Hospital, Stockholm* | *Sweden* | *120* | *67* | *12* | *7* | *26* | *8* |
| *King's College Hospital, London* | *UK* | *119* | *78* | *13* | *8* | *15* | *5* |
| Universita degli Studi di Napoli, Naples | Italy | 103 | 82 | 2 | 3 | 13 | 3 |
| Hôpital Boucicaut, Paris | France | 80 | 71 | 2 | 2 | 5 | 0 |
| *Skåne University Hospital Lund* | *Sweden* | *77* | *57* | *2* | *4* | *11* | *3* |
| *Chinese PLA General Hospital, Beijing* | *China* | *73* | *57* | *1* | *0* | *12* | *3* |
| *Universita degli Studi di Udine* | *Italy* | *64* | *45* | *1* | *10* | *6* | *2* |
| Centre Medical des Pyramides, Maurepas | France | 64 | 57 | 1 | 4 | 2 | 0 |
| Institut Universitari Dexeus, Barcelona | Spain | 37 | 26 | 8 | 2 | 1 | 0 |
| Macedonio Melloni Hospital | Italy | 21 | 17 | 1 | 2 | 1 | 0 |
| Ospedale dei Bambini Vittore Buzzi, Milan | Italy | 21 | 21 | 0 | 0 | 0 | 0 |
| *Istituto Nazionale dei Tumori, Naples* | *Italy* | *15* | *7* | *0* | *2* | *4* | *2* |
| St Joseph's Hospital, Hamilton | Canada | 11 | 10 | 0 | 1 | 0 | 0 |
| | | VALIDATION DATA | | | | | |
| *Alexandra Hospital, Athens* | *Greece* | *360* | *294* | *14* | *12* | *27* | *13* |
| *Universita Cattolica del Sacro Cuore, Rome* | *Italy* | *354* | *185* | *25* | *36* | *84* | *24* |
| *Istituto Europeo di Oncologia, Milan* | *Italy* | *286* | *125* | *27* | *28* | *99* | *7* |
| Skåne University Hospital Malmö | Sweden | 278 | 205 | 19 | 14 | 31 | 9 |
| *Karolinska University Hospital, Stockholm* | *Sweden* | *239* | *106* | *26* | *26* | *59* | *22* |
| *University of Bologna* | *Italy* | *231* | *144* | *29* | *15* | *32* | *11* |
| Ziekenhuis Oost-Limburg, Genk | Belgium | 211 | 167 | 14 | 10 | 11 | 9 |
| *University Hospitals Leuven* | *Belgium* | *202* | *110* | *22* | *17* | *39* | *14* |
| *Ospedale San Gerardo, Monza* | *Italy* | *145* | *73* | *14* | *18* | *33* | *7* |
| Ospedale San Giovanni di Dio, Cagliari | Italy | 121 | 96 | 8 | 0 | 11 | 6 |
| *Universita degli Studi di Udine* | *Italy* | *116* | *87* | *6* | *3* | *16* | *4* |
| *Di Venere Hospital, Bari* | *Italy* | *106* | *82* | *2* | *8* | *12* | *2* |
| *Medical University Lublin* | *Poland* | *92* | *60* | *5* | *6* | *16* | *5* |
| *General Faculty Hospital, Prague* | *Czech Rep.* | *90* | *20* | *20* | *11* | *26* | *13* |
| *National Cancer Institute, Milan* | *Italy* | *58* | *16* | *5* | *4* | *23* | *10* |
| *Clinica Universidad de Navarra, Pamplona* | *Spain* | *54* | *27* | *3* | *3* | *11* | *10* |
| IRCCS Materno Infantile Burlo Garofolo, Trieste | Italy | 48 | 33 | 6 | 1 | 6 | 2 |
| *Medical University of Silesia, Katowice* | *Poland* | *41* | *24* | *6* | *2* | *9* | *0* |
| *Chinese PLA General Hospital, Beijing* | *China* | *33* | *17* | *3* | *0* | *10* | *3* |
| Cairo University | Egypt | 29 | 22 | 1 | 3 | 3 | 0 |
| University of Florence | Italy | 28 | 26 | 1 | 1 | 0 | 0 |
| Whipps Cross Hospital, London (*) | UK | 25 | 24 | 1 | 0 | 0 | 0 |
| DCS Sacco University of Milan | Italy | 26 | 25 | 0 | 0 | 0 | 1 |
| Queen Charlotte's and Chelsea Hospital, London | UK | 13 | 8 | 1 | 1 | 3 | 0 |
| Ospedale dei Bambini Vittore Buzzi, Milan | Italy | 13 | 12 | 1 | 0 | 0 | 0 |

Ben, benign; Bot, borderline; S1, Stage I primary invasive; S2-4, Stage II-IV primary invasive; Met, Secondary Metastasis.
Oncology centers are in italic.
(*) In previous publications, it was incorrectly stated that this center was located in Nottingham.

**Table S3. Pairwise area under the receiver operating characteristic curve (AUROC) values (with 95% CI) for models with CA125 on external validation data.**

| Pair | Standard MLR | Ridge MLR | RF | XGBoost | NN | SVM |
|---|---|---|---|---|---|---|
| Ben vs Bot | 0.86 (0.80; 0.90) | 0.84 (0.80; 0.89) | 0.88 (0.81; 0.92) | 0.88 (0.82; 0.92) | 0.87 (0.80; 0.91) | 0.76 (0.68; 0.82) |
| Ben vs S1 | 0.91 (0.88; 0.94) | 0.89 (0.85; 0.92) | 0.91 (0.88; 0.94) | 0.92 (0.89; 0.94) | 0.91 (0.88; 0.94) | 0.89 (0.85; 0.92) |
| Ben vs S2-4 | 0.97 (0.96; 0.98) | 0.96 (0.94; 0.97) | 0.97 (0.96; 0.98) | 0.97 (0.96; 0.98) | 0.97 (0.96; 0.98) | 0.95 (0.94; 0.97) |
| Ben vs Met | 0.92 (0.89; 0.95) | 0.91 (0.88; 0.94) | 0.92 (0.87; 0.94) | 0.92 (0.88; 0.95) | 0.92 (0.89; 0.95) | 0.91 (0.88; 0.94) |
| Bot vs S1 | 0.77 (0.71, 0.82) | 0.72 (0.65; 0.77) | 0.78 (0.72; 0.84) | 0.78 (0.72; 0.82) | 0.77 (0.71; 0.82) | 0.75 (0.69; 0.80) |
| Bot vs S2-4 | 0.92 (0.88; 0.94) | 0.91 (0.87; 0.93) | 0.92 (0.89; 0.94) | 0.91 (0.88; 0.94) | 0.92 (0.89; 0.94) | 0.89 (0.86; 0.92) |
| Bot vs Met | 0.85 (0.80; 0.89) | 0.86 (0.80; 0.90) | 0.86 (0.79; 0.90) | 0.87 (0.81; 0.91) | 0.88 (0.83; 0.92) | 0.85 (0.80; 0.89) |
| S1 vs S2-4 | 0.80 (0.75; 0.85) | 0.80 (0.75; 0.84) | 0.81 (0.76; 0.85) | 0.81 (0.76; 0.85) | 0.81 (0.76; 0.85) | 0.69 (0.63; 0.73) |
| S1 vs Met | 0.75 (0.67; 0.82) | 0.72 (0.64; 0.78) | 0.71 (0.64; 0.78) | 0.75 (0.69; 0.81) | 0.73 (0.66; 0.79) | 0.68 (0.60; 0.74) |
| S2-4 vs Met | 0.79 (0.71; 0.85) | 0.77 (0.70; 0.84) | 0.76 (0.69; 0.81) | 0.77 (0.70; 0.83) | 0.77 (0.70; 0.83) | 0.53 (0.45; 0.61) |

Ben, benign; Bot, borderline tumor; S1, Stage I primary invasive; S2-4, Stage II-IV primary invasive; Met, secondary metastasis; MLR, multinomial logistic regression; RF, random forest; NN, neural network; SVM, support vector machine.

**Table S4. Pairwise area under the receiver operating characteristic curve (AUROC) values for models without CA125 on validation data.**

| Pair | Standard MLR | Ridge MLR | RF | XGBoost | NN | SVM |
|---|---|---|---|---|---|---|
| Ben vs Bot | 0.85<br>(0.81; 0.89) | 0.84<br>(0.80; 0.87) | 0.87<br>(0.81; 0.91) | 0.87<br>(0.81; 0.91) | 0.87<br>(0.80; 0.91) | 0.76<br>(0.70; 0.81) |
| Ben vs S1 | 0.91<br>(0.88; 0.94) | 0.89<br>(0.85; 0.92) | 0.91<br>(0.88; 0.94) | 0.92<br>(0.88; 0.94) | 0.92<br>(0.89; 0.94) | 0.89<br>(0.85; 0.91) |
| Ben vs S2-4 | 0.96<br>(0.94; 0.97) | 0.94<br>(0.92; 0.95) | 0.96<br>(0.94; 0.97) | 0.96<br>(0.94; 0.97) | 0.95<br>(0.94; 0.97) | 0.95<br>(0.93; 0.96) |
| Ben vs Met | 0.91<br>(0.87; 0.94) | 0.91<br>(0.87; 0.93) | 0.91<br>(0.87; 0.94) | 0.91<br>(0.87; 0.94) | 0.92<br>(0.88; 0.94) | 0.91<br>(0.87; 0.94) |
| Bot vs S1 | 0.77<br>(0.71, 0.82) | 0.72<br>(0.66; 0.77) | 0.79<br>(0.74; 0.84) | 0.78<br>(0.72; 0.84) | 0.78<br>(0.72; 0.84) | 0.75<br>(0.69; 0.80) |
| Bot vs S2-4 | 0.91<br>(0.87; 0.94) | 0.88<br>(0.84; 0.91) | 0.90<br>(0.86; 0.93) | 0.90<br>(0.86; 0.93) | 0.91<br>(0.87; 0.94) | 0.89<br>(0.85; 0.91) |
| Bot vs Met | 0.86<br>(0.80; 0.90) | 0.85<br>(0.80; 0.89) | 0.85<br>(0.78; 0.90) | 0.86<br>(0.80; 0.91) | 0.86<br>(0.81; 0.90) | 0.86<br>(0.81; 0.90) |
| S1 vs S2-4 | 0.74<br>(0.69; 0.78) | 0.74<br>(0.69; 0.78) | 0.73<br>(0.67; 0.78) | 0.73<br>(0.68; 0.78) | 0.75<br>(0.70; 0.80) | 0.71<br>(0.66; 0.75) |
| S1 vs Met | 0.74<br>(0.67; 0.81) | 0.72<br>(0.64; 0.78) | 0.72<br>(0.65; 0.77) | 0.72<br>(0.66; 0.78) | 0.73<br>(0.65; 0.80) | 0.68<br>(0.60; 0.74) |
| S2-4 vs Met | 0.69<br>(0.63; 0.74) | 0.65<br>(0.59; 0.70) | 0.60<br>(0.54; 0.66) | 0.61<br>(0.55; 0.66) | 0.67<br>(0.61; 0.72) | 0.58<br>(0.50; 0.66) |

Abbreviations: AUROC, area under the receiver operating characteristic curve; MLR, multinomial logistic regression; RF, random forest; XGBoost, extreme gradient boosting; NN, neural network; SVM, support vector machine; Ben, benign; Bot, borderline; S1, Stage I; S2-4, Stage II-IV; Met, Secondary metastasis.

**Table S5. Percentage of patients on validation data falling on opposite sides of the 10% risk of malignancy threshold when comparing two models.**

| Comparison | Models with CA125 | Models without CA125 |
|---|---|---|
| Standard MLR vs Ridge MLR | 200 (6%) | 350 (11%) |
| Standard MLR vs RF | 233 (7%) | 199(6%) |
| Standard MLR vs XGBoost | 143 (5%) | 160 (5%) |
| Standard MLR vs NN | 132 (4%) | 156 (5%) |
| Standard MLR vs SVM | 566 (18%) | 905 (28%) |
| Ridge MLR vs RF | 233 (7%) | 321 (10%) |
| Ridge MLR vs XGBoost | 191 (6%) | 310 (10%) |
| Ridge MLR vs NN | 152 (5%) | 414 (13%) |
| Ridge MLR vs SVM | 464 (15%) | 587 (18%) |
| RF vs XGBoost | 142 (5%) | 145 (5%) |
| RF vs NN | 203 (6%) | 221 (7%) |
| RF vs SVM | 487 (15%) | 828 (26%) |
| XGBoost vs NN | 109 (3%) | 168 (5%) |
| XGBoost vs SVM | 503 (16%) | 855 (27%) |
| NN vs SVM | 544 (17%) | 965 (30%) |

MLR, multinomial logistic regression; RF, random forest; XGBoost, extreme gradient boosting; NN, neural network; SVM, support vector machine.

**Figure S1. Polytomous discrimination index for models with CA125 on external validation data.**

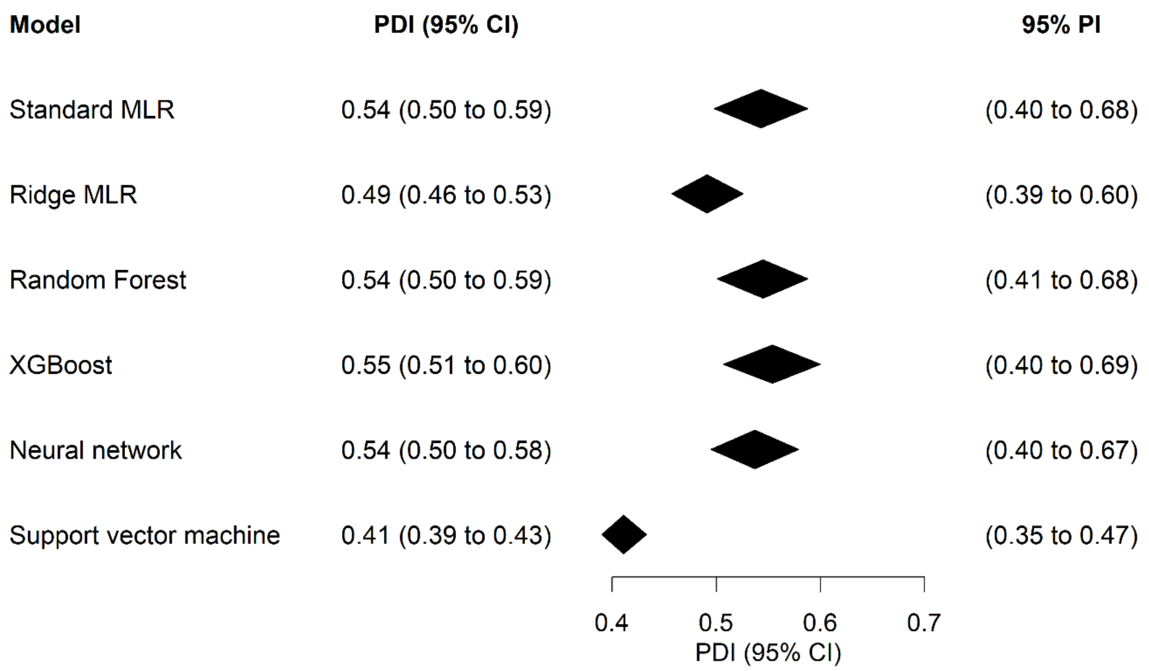

Abbreviations: PDI, polytomous discrimination index; CI, confidence interval; PI, prediction interval; MLR, multinomial logistic regression; XGBoost, extreme gradient boosting.

**Figure S2. AUROC for benign tumors vs any malignancy for models with CA125 on validation data.**

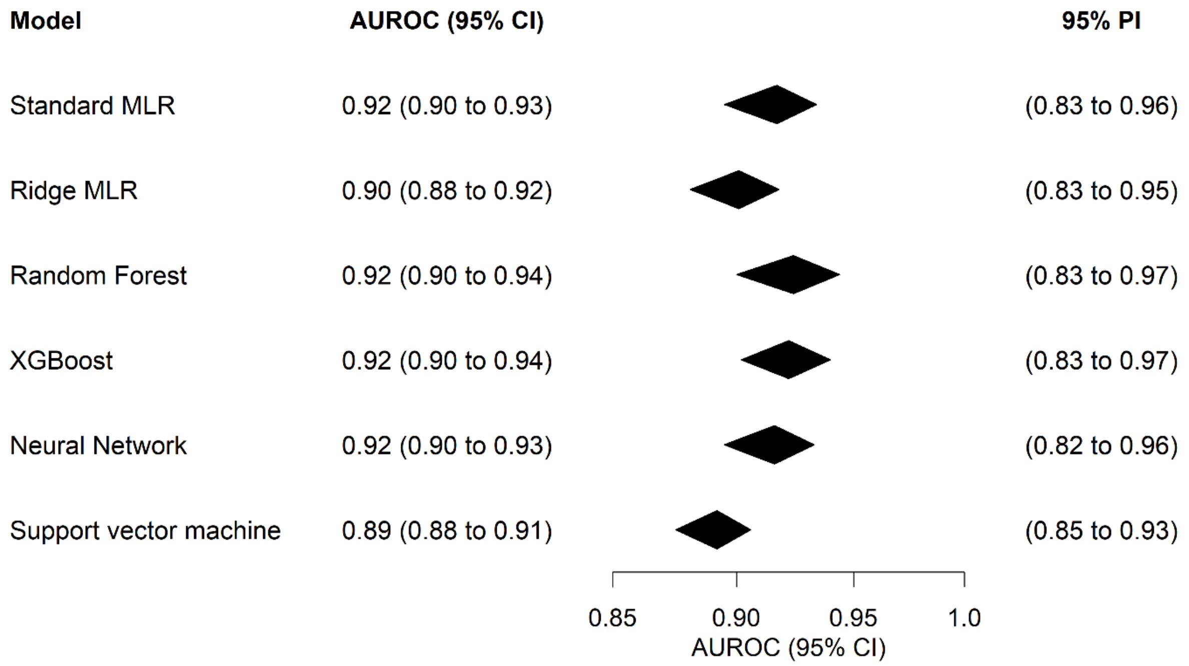

Abbreviations: AUROC, area under the receiver operating characteristic curve; CI, confidence interval; PI, prediction interval; MLR, multinomial logistic regression; XGBoost, extreme gradient boosting.

**Figure S3. Polytomous discrimination index for models without CA125 on validation data.**

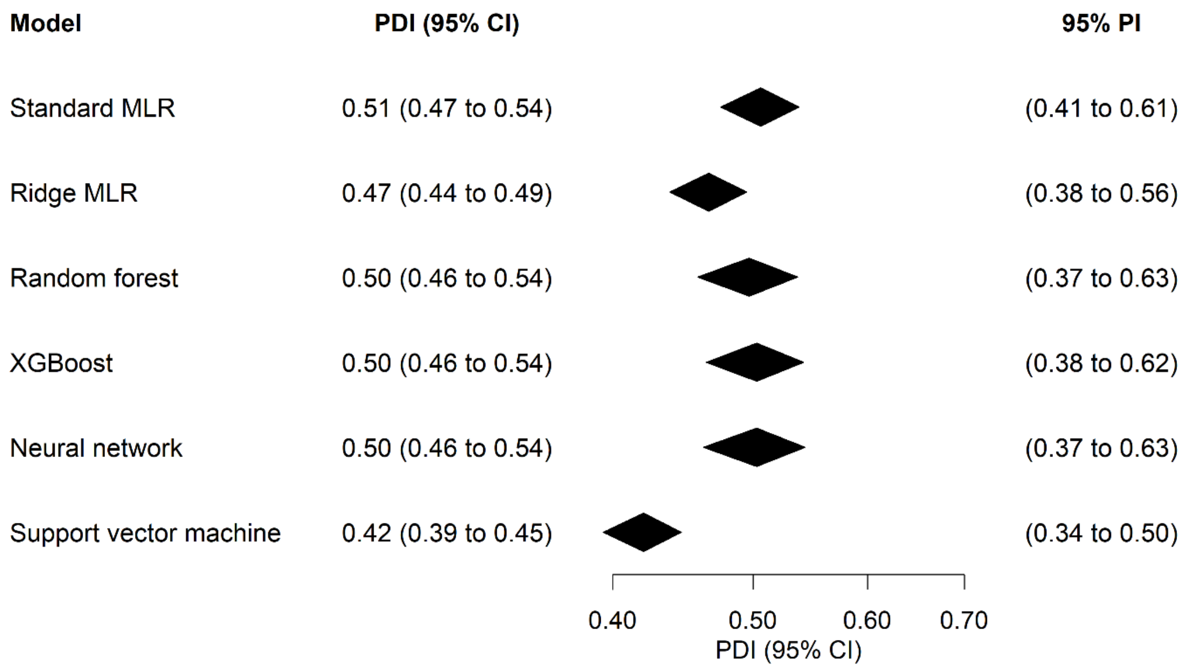

Abbreviations: PDI, polytomous discrimination index; CI, confidence interval; PI, prediction interval; MLR, multinomial logistic regression; XGBoost, extreme gradient boosting.

**Figure S4. Area under the receiver operating characteristic curve for benign tumors vs any malignancy for models without CA125 on validation data.**

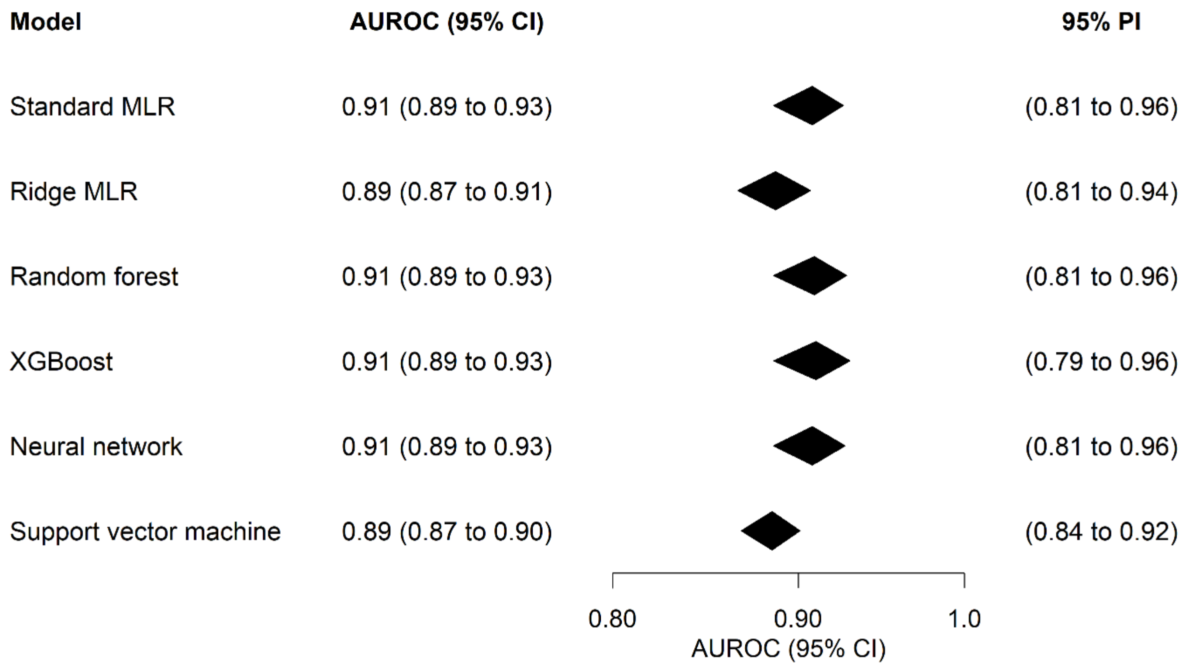

Abbreviations: AUROC, area under the receiver operating characteristic curve; CI, confidence interval; PI, prediction interval; MLR, multinomial logistic regression; XGBoost, extreme gradient boosting.

**Figure S5. Box plots of estimated probabilities for standard MLR with CA125 on validation data.**

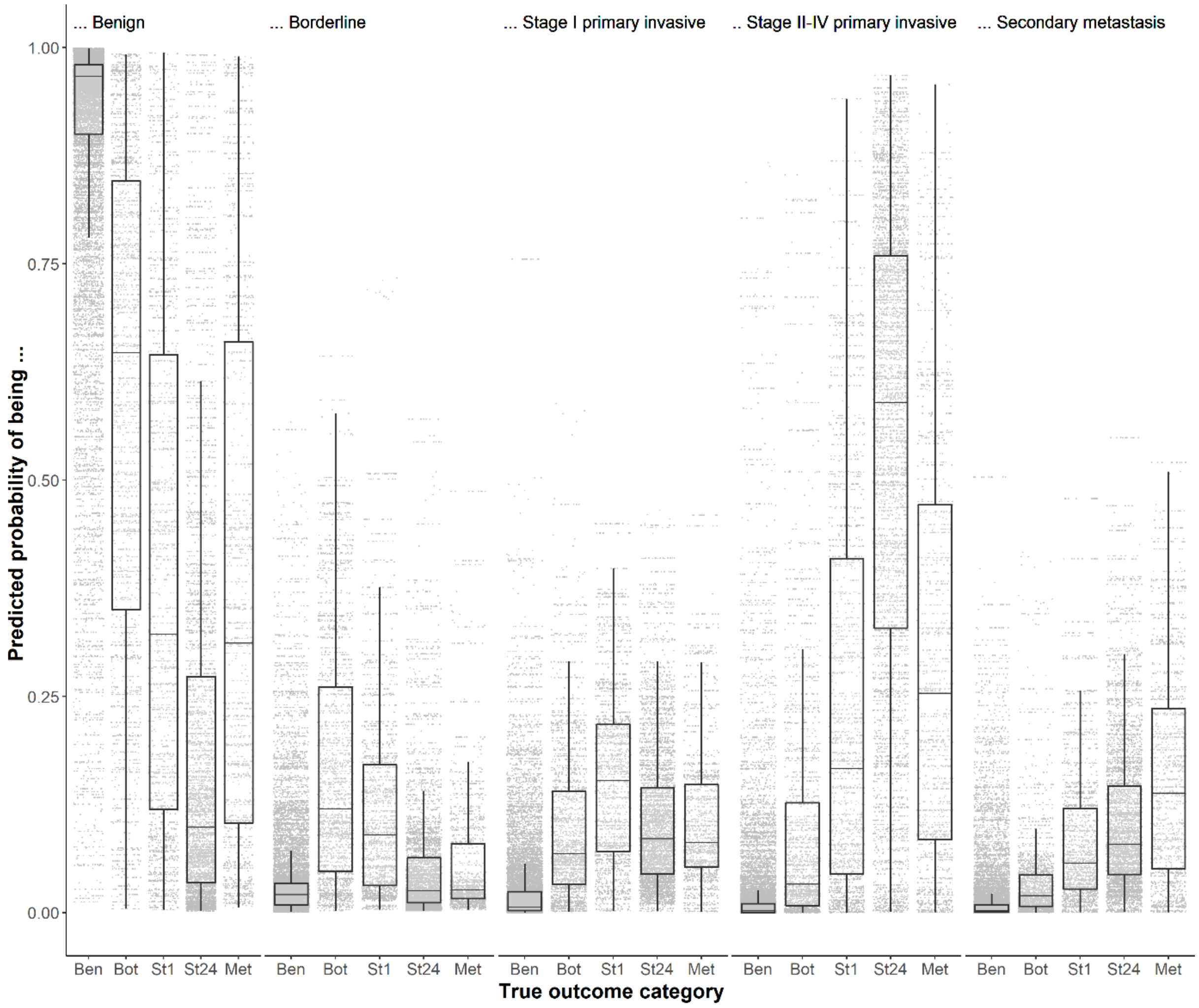

Abbreviations: MLR, multinomial logistic regression; Ben, benign; Bot, borderline; St1, stage I primary invasive; St24, stage II-IV primary invasive, Met, secondary metastatic.

**Figure S6. Box plots of estimated probabilities for ridge MLR with CA125 on validation data.**

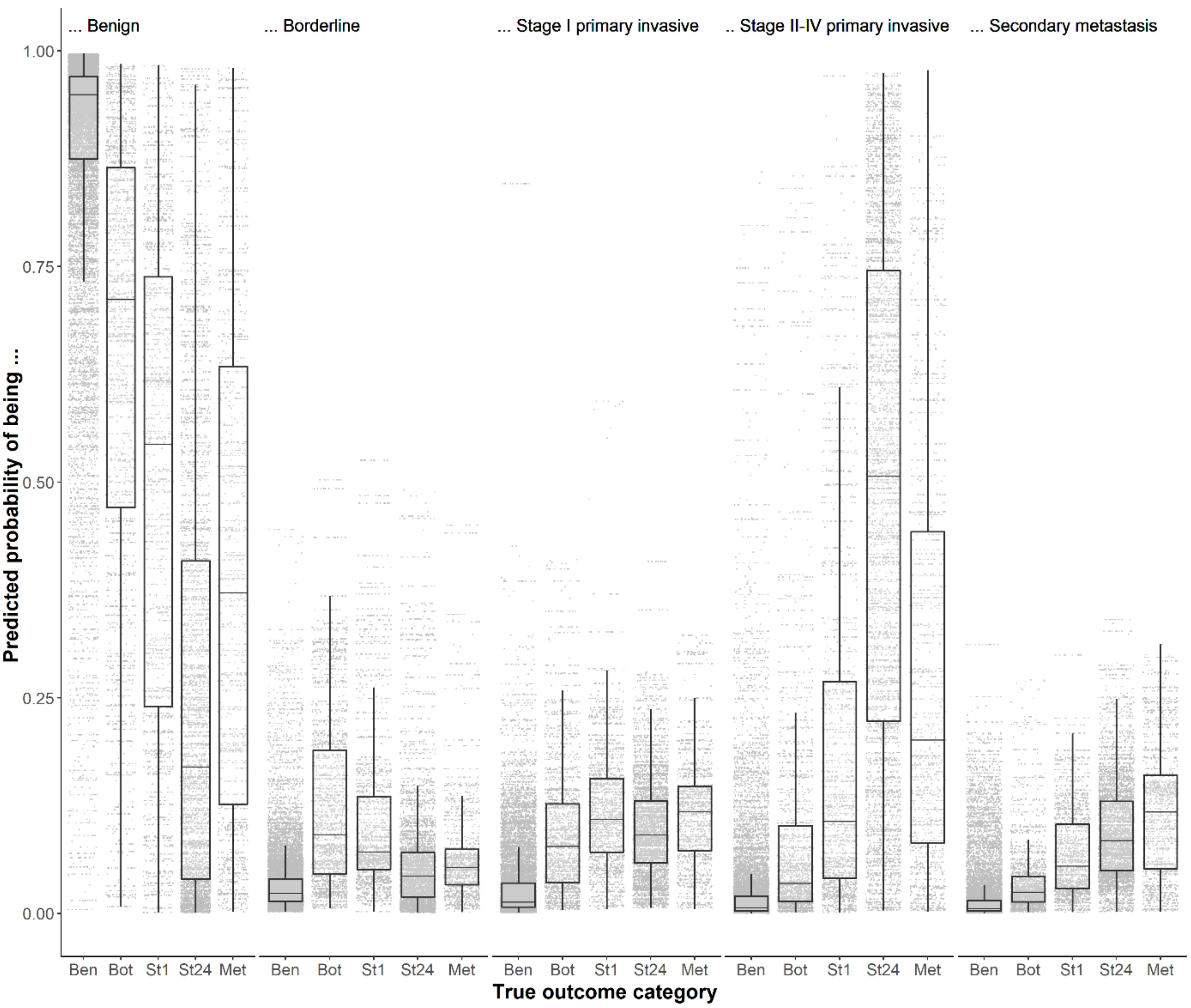

Abbreviations: MLR, multinomial logistic regression; Ben, benign; Bot, borderline; St1, stage I primary invasive; St24, stage II-IV primary invasive, Met, secondary metastatic.

**Figure S7. Box plots of estimated probabilities for random forest with CA125 on validation data.**

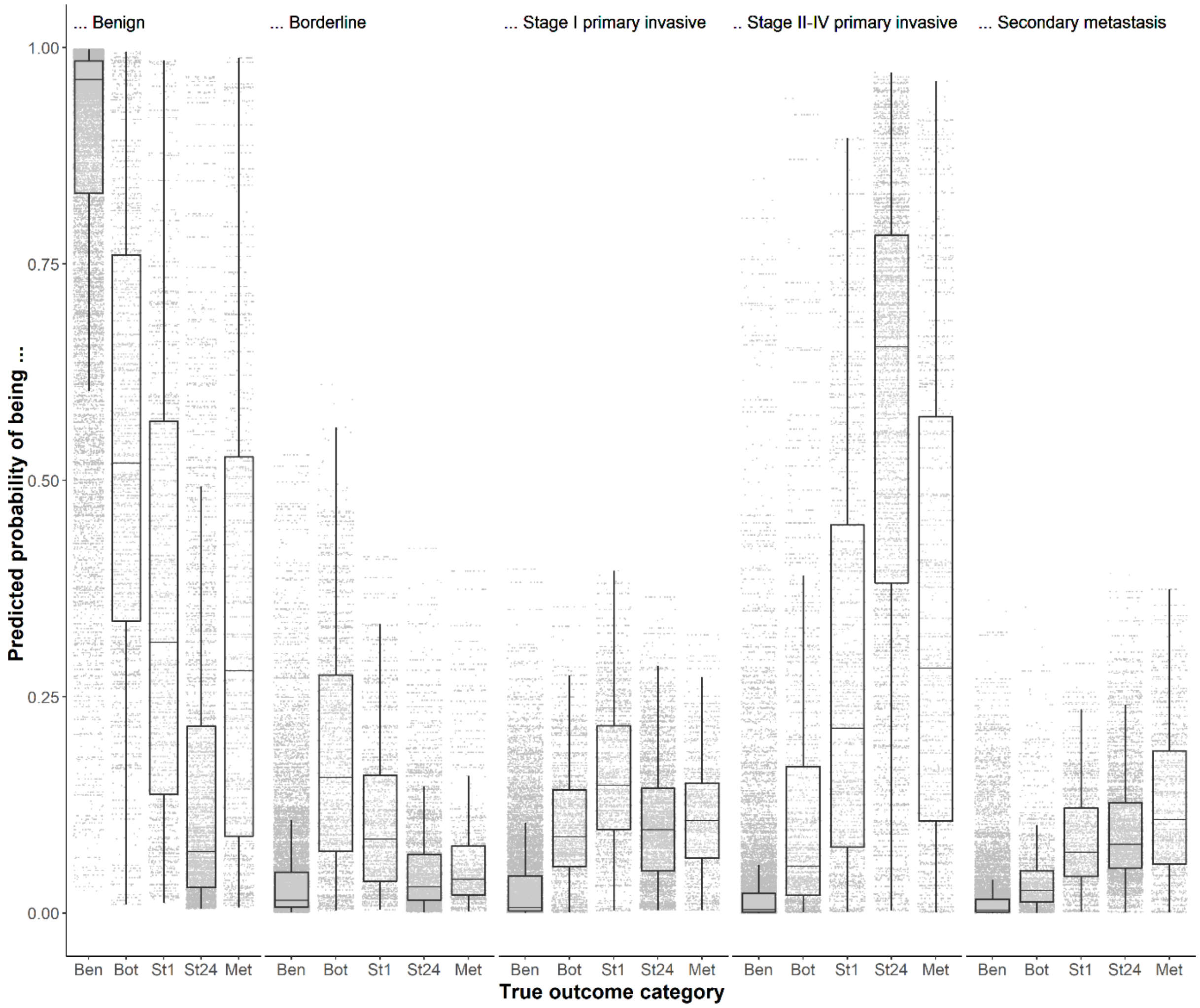

Abbreviations: Ben, benign; Bot, borderline; St1, stage I primary invasive; St24, stage II-IV primary invasive, Met, secondary metastatic.

**Figure S8. Box plots of estimated probabilities for extreme gradient boosting (XGBoost) with CA125 on validation data.**

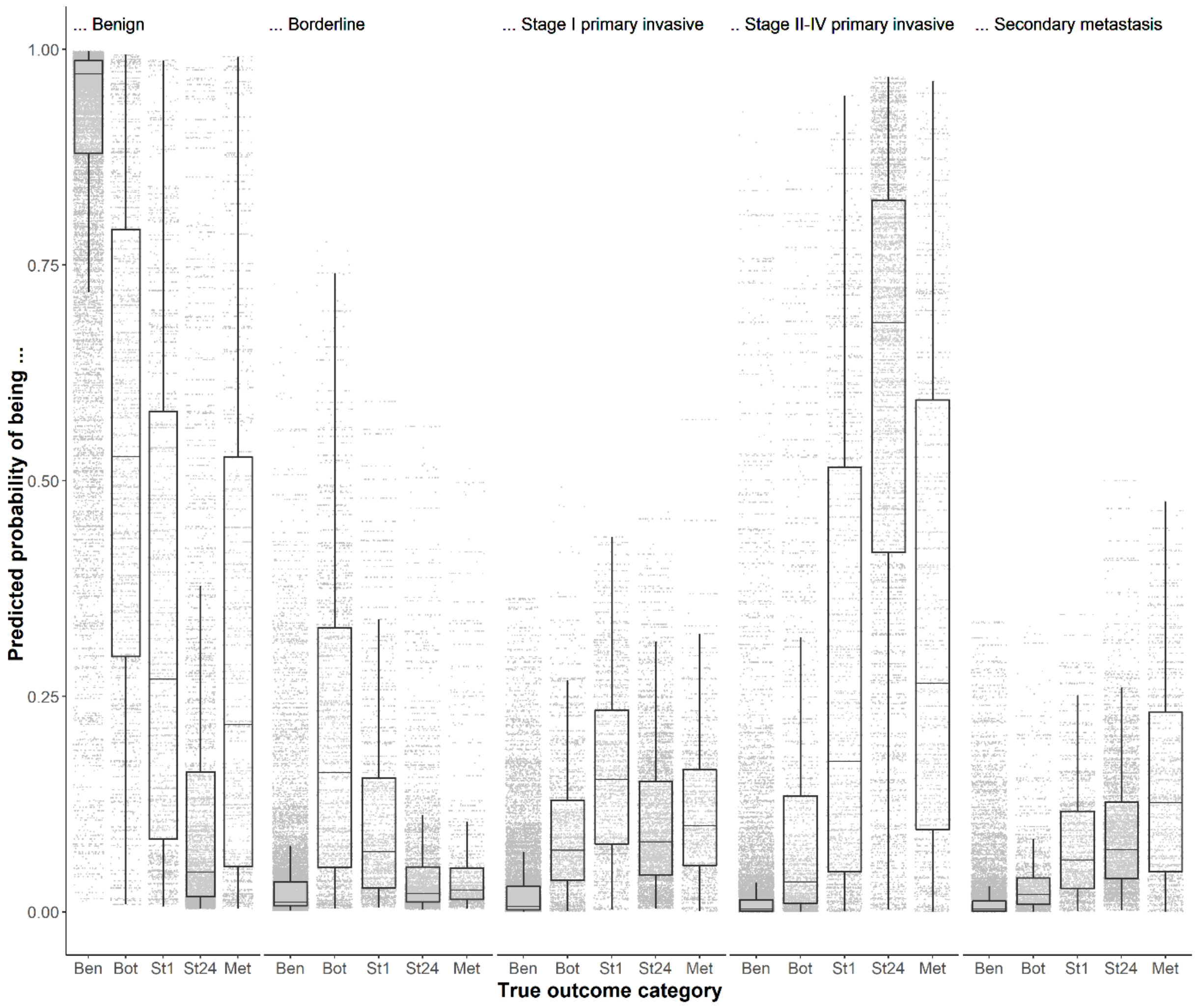

Abbreviations: Ben, benign; Bot, borderline; St1, stage I primary invasive; St24, stage II-IV primary invasive, Met, secondary metastatic.

**Figure S9. Box plots of estimated probabilities for neural network with CA125 on validation data.**

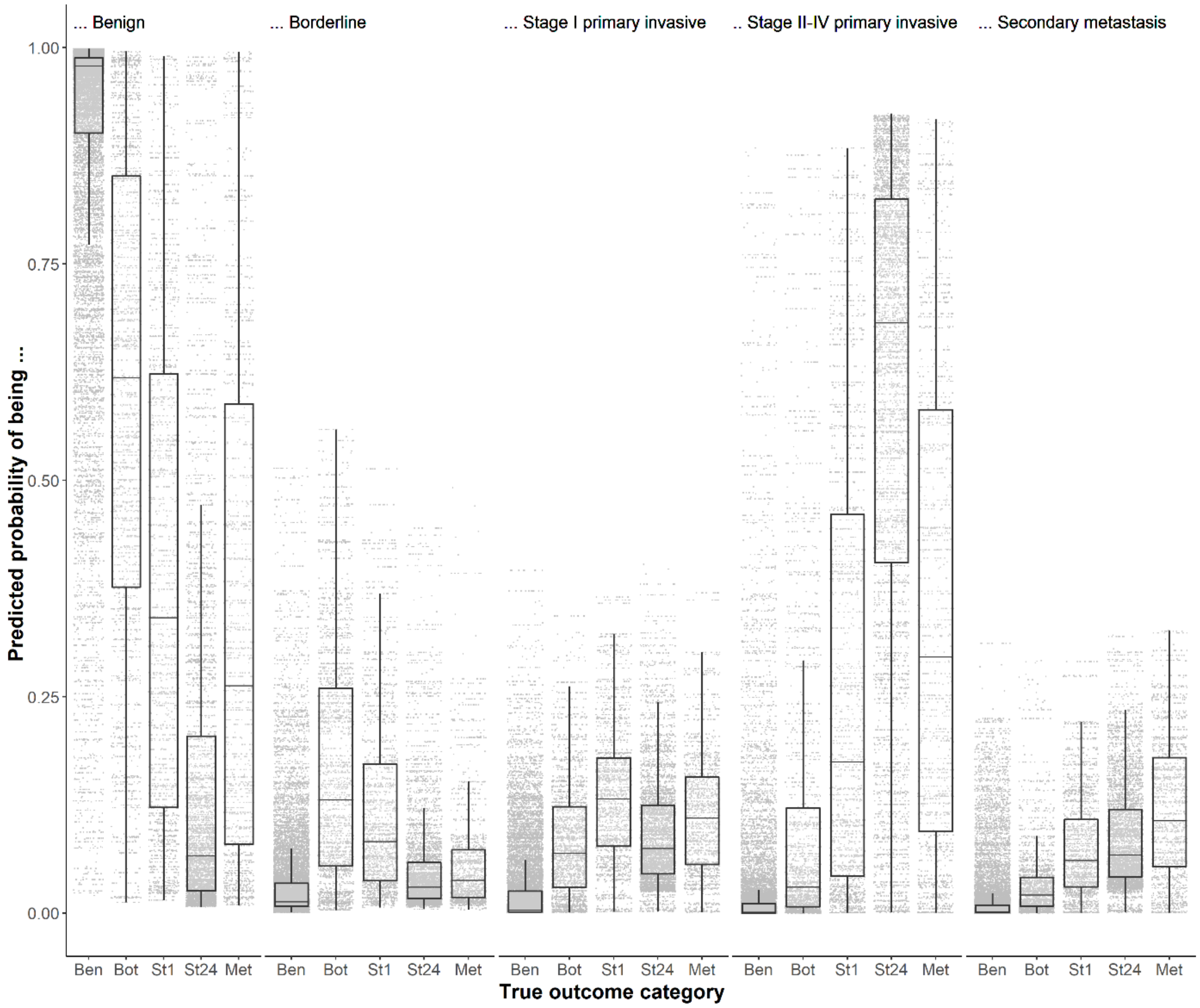

Abbreviations: Ben, benign; Bot, borderline; St1, stage I primary invasive; St24, stage II-IV primary invasive, Met, secondary metastatic.

**Figure S10. Box plots of estimated probabilities for support vector machine with CA125 on validation data.**

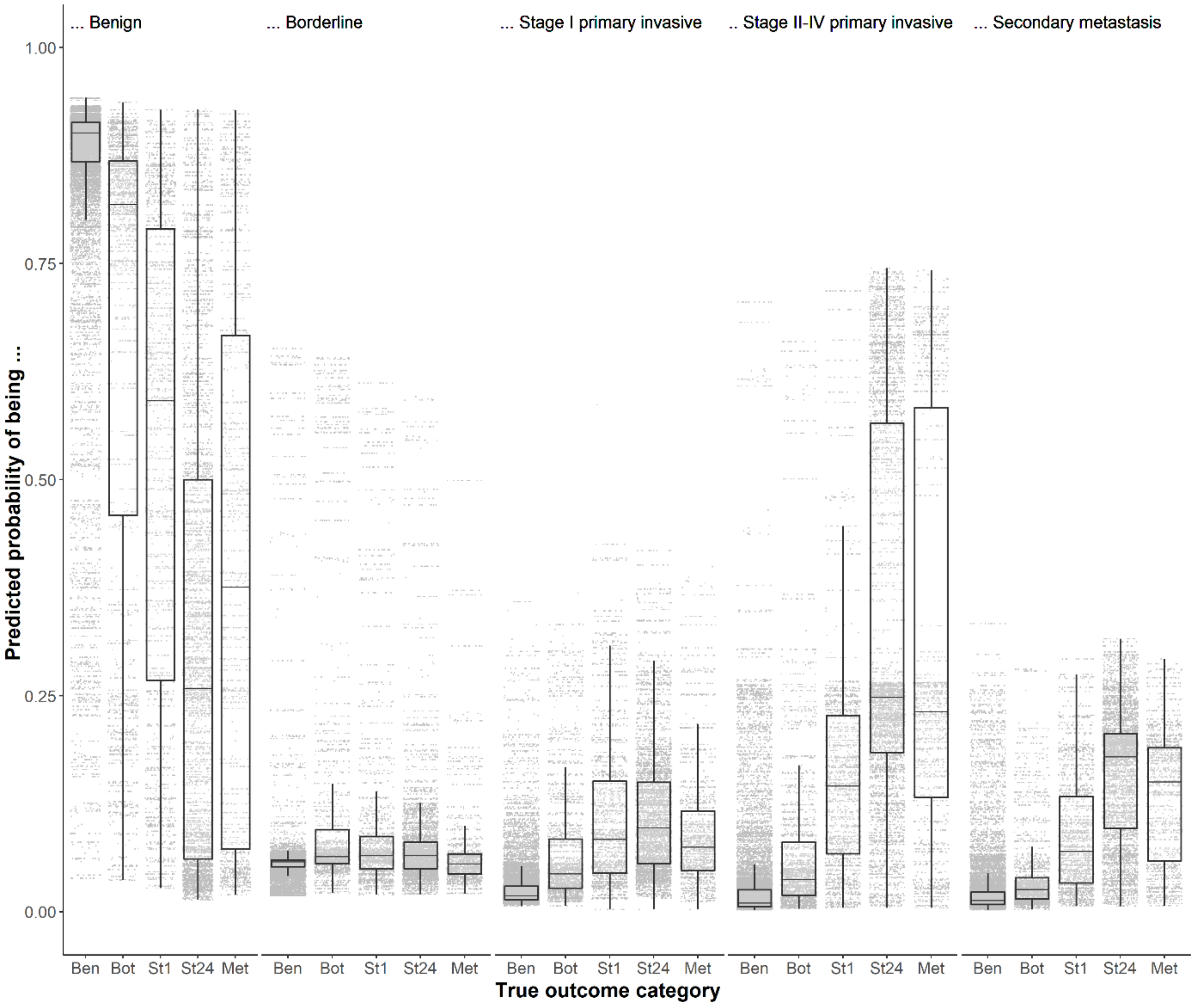

Abbreviations: Ben, benign; Bot, borderline; St1, stage I primary invasive; St24, stage II-IV primary invasive, Met, secondary metastatic.

**Figure S11. Flexible calibration curves for models without CA125 on validation data.**

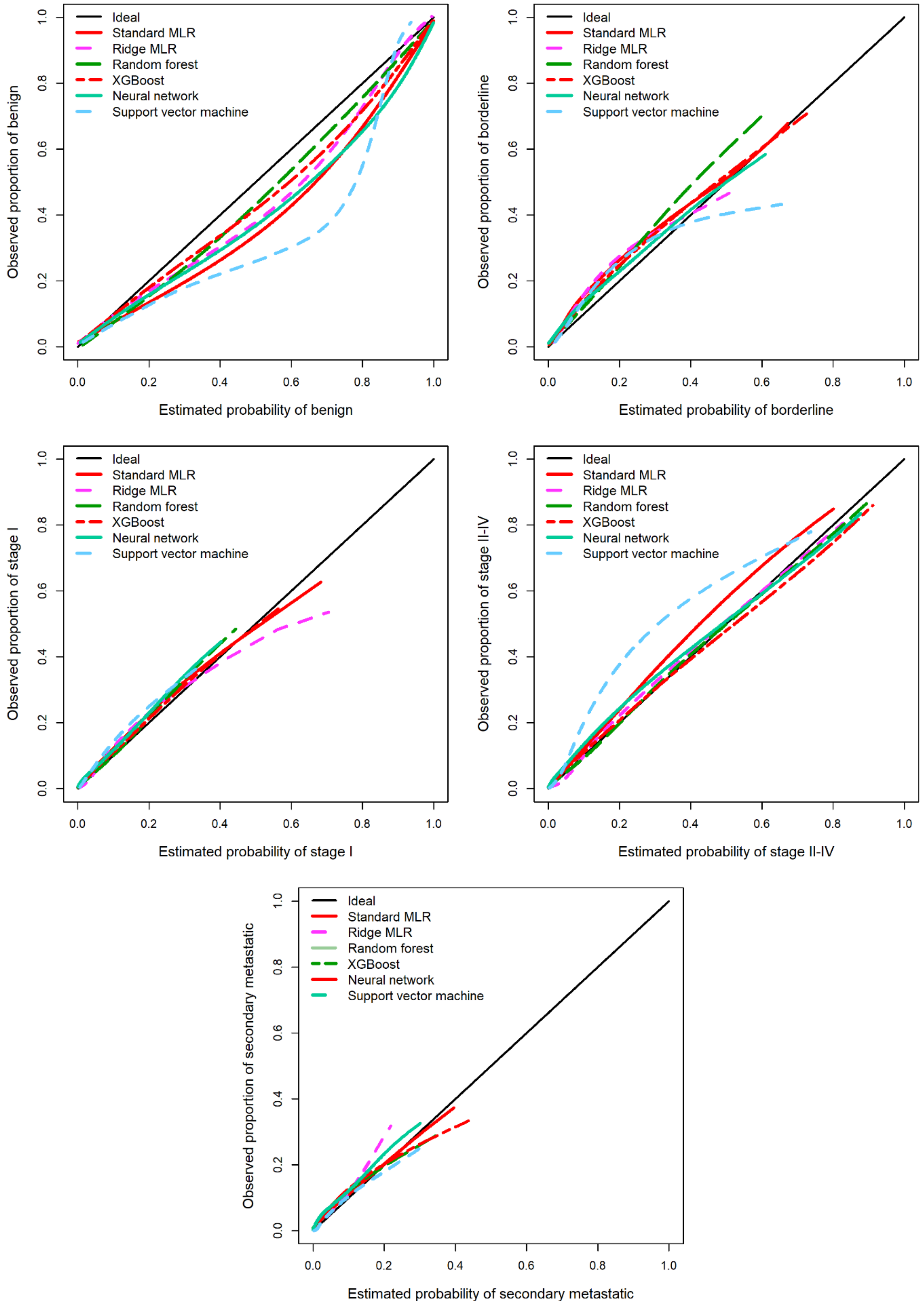

Abbreviations: MLR, multinomial logistic regression; XGBoost, extreme gradient boosting.

**Figure S12. Box plots of estimated probabilities for standard MLR without CA125 on validation data.**

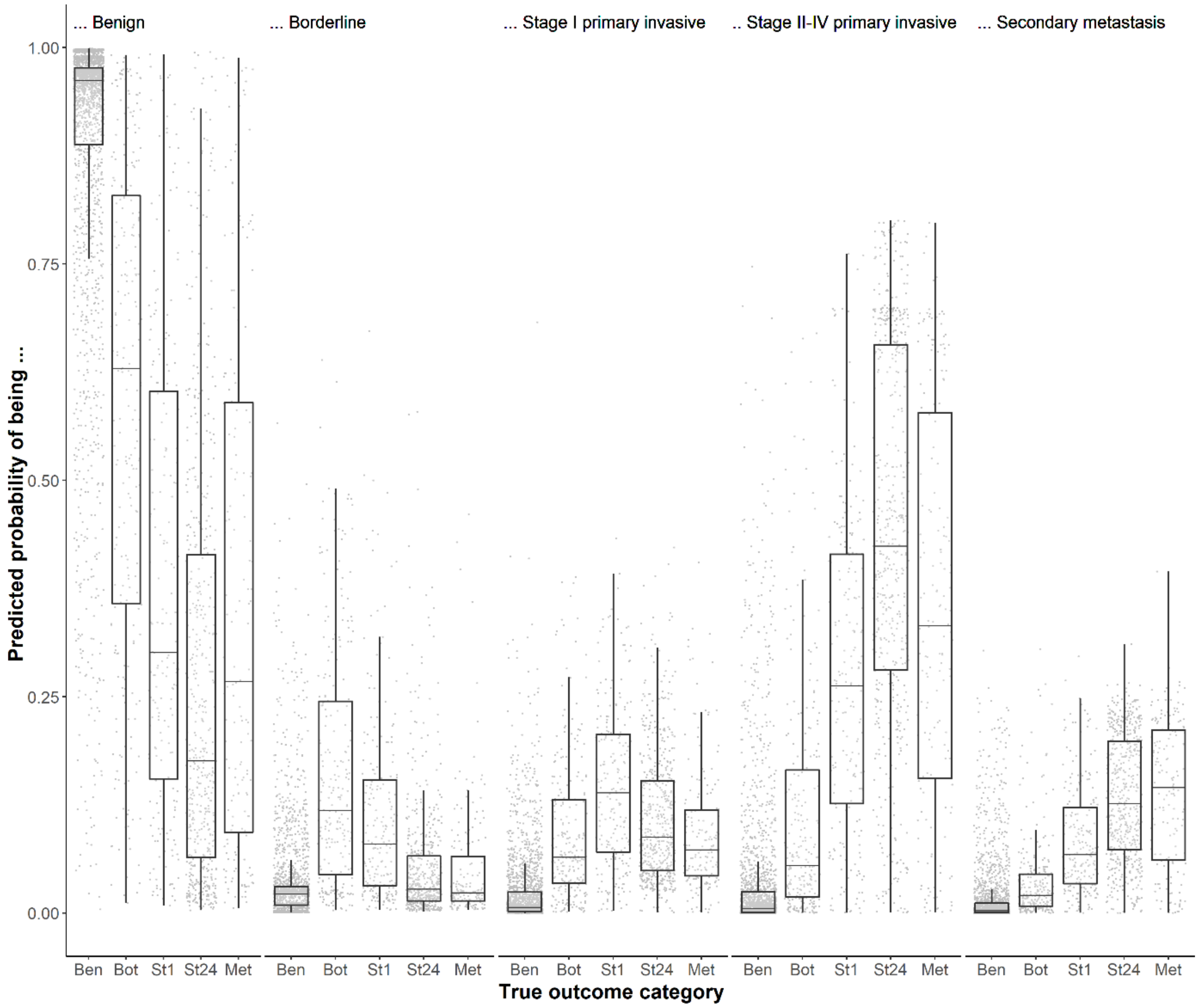

Abbreviations: MLR, multinomial logistic regression; Ben, benign; Bot, borderline; St1, stage I primary invasive; St24, stage II-IV primary invasive, Met, secondary metastatic.

**Figure S13. Box plots of estimated probabilities for ridge MLR without CA125 on validation data.**

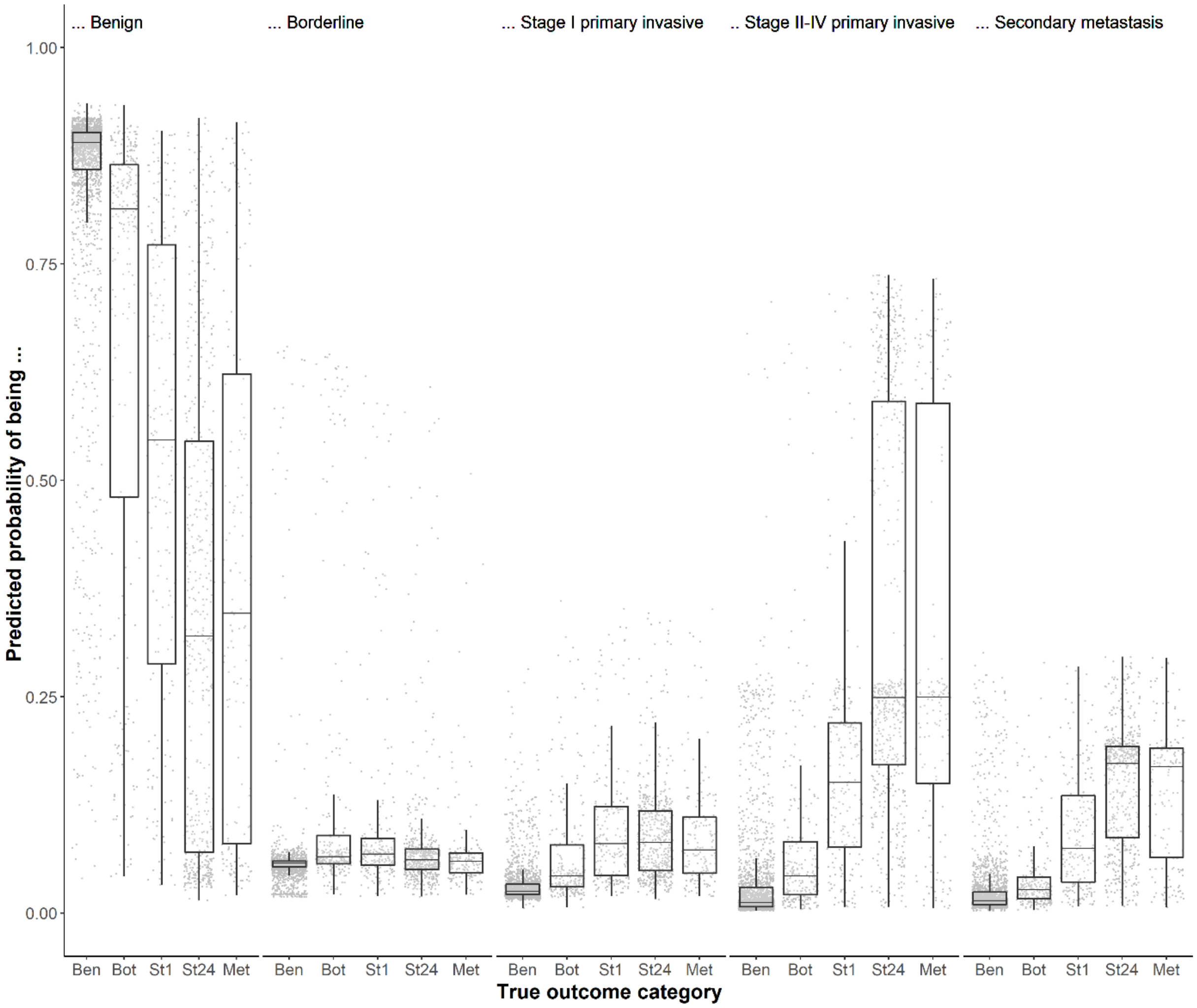

Abbreviations: MLR, multinomial logistic regression; Ben, benign; Bot, borderline; St1, stage I primary invasive; St24, stage II-IV primary invasive, Met, secondary metastatic.

**Figure S14. Box plots of estimated probabilities for random forest without CA125 on validation data.**

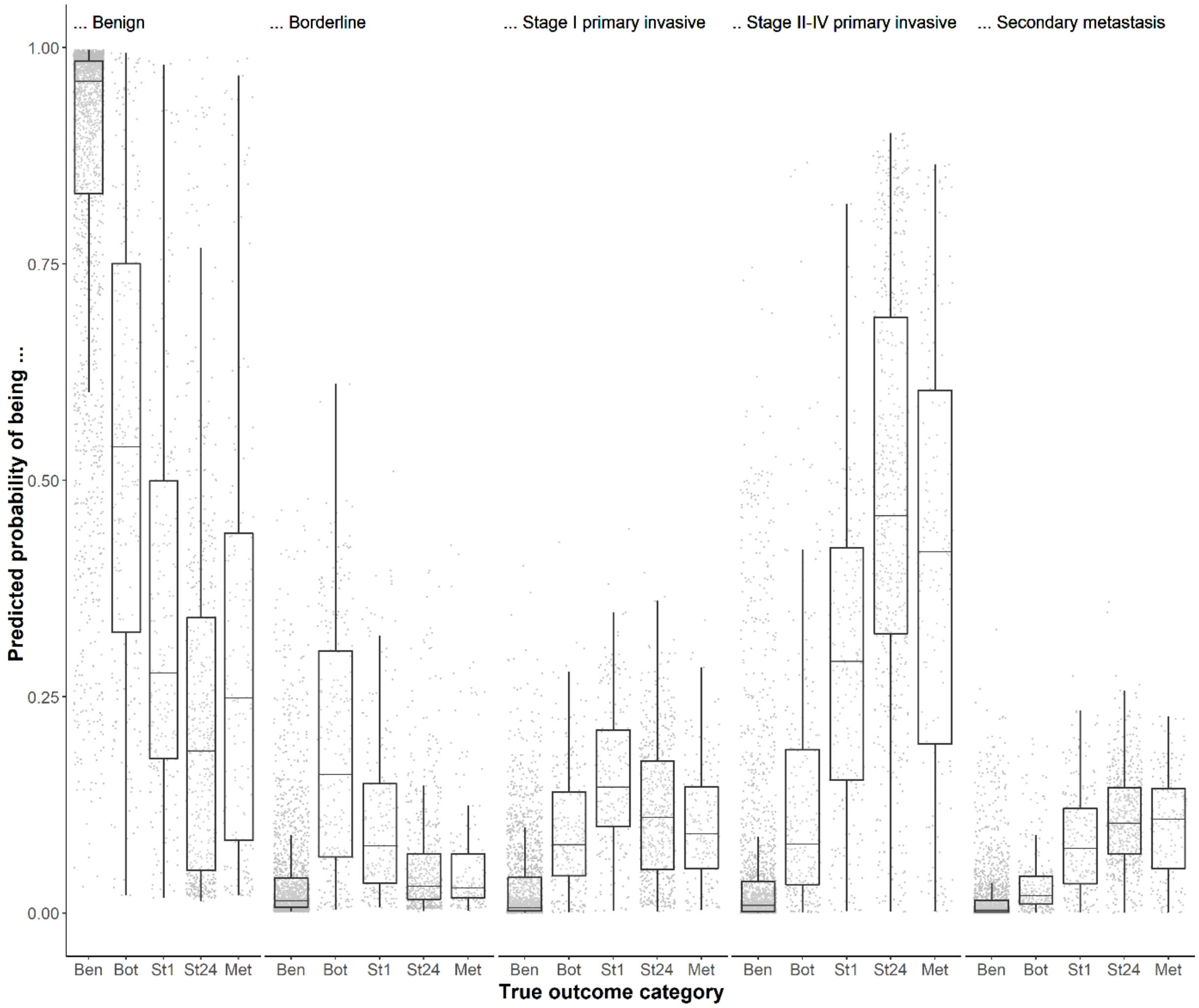

Abbreviations: Ben, benign; Bot, borderline; St1, stage I primary invasive; St24, stage II-IV primary invasive, Met, secondary metastatic.

**Figure S15. Box plots of estimated probabilities for extreme gradient boosting (XGBoost) without CA125 on validation data.**

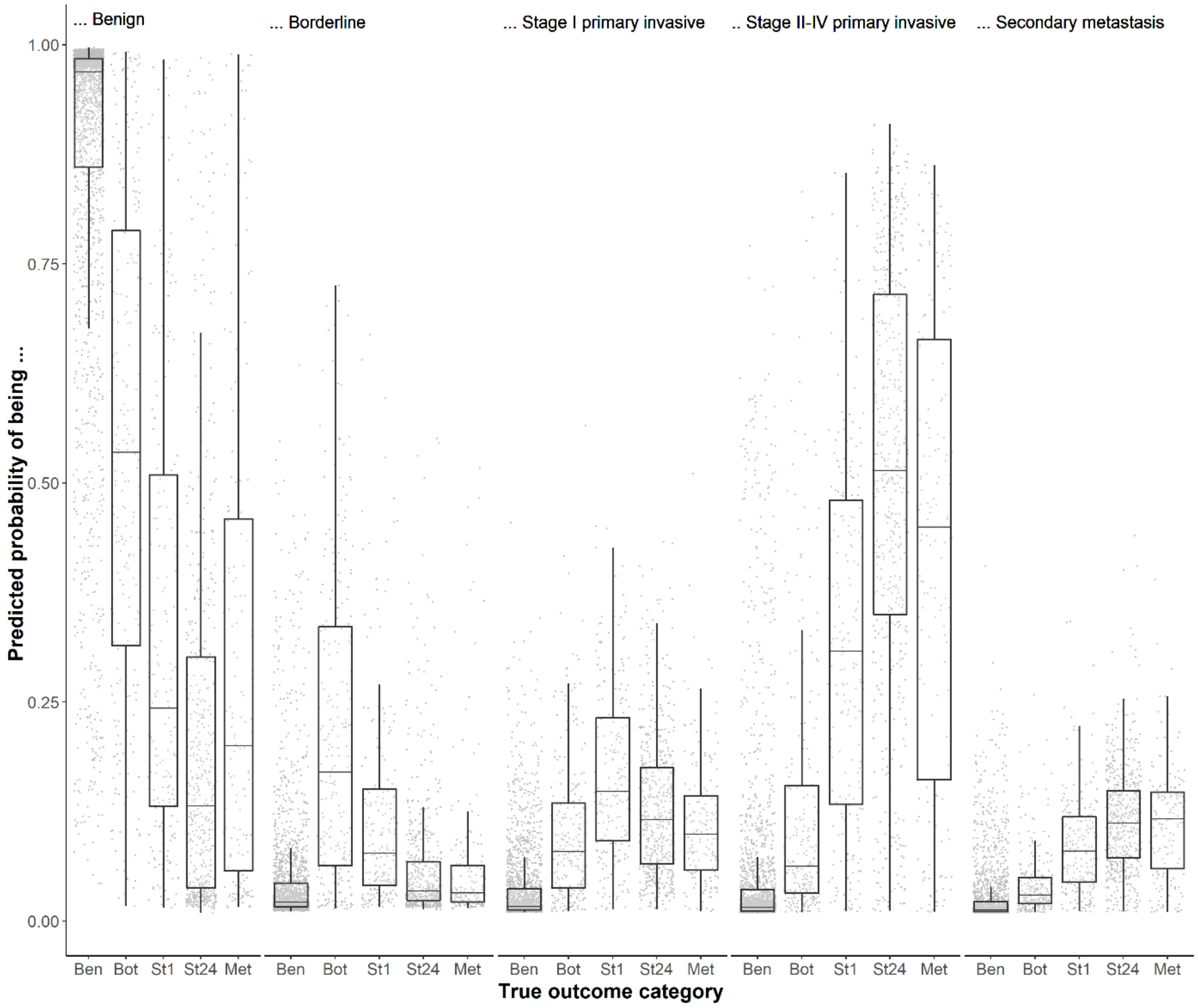

Abbreviations: Ben, benign; Bot, borderline; St1, stage I primary invasive; St24, stage II-IV primary invasive, Met, secondary metastatic.

**Figure S16. Box plots of estimated probabilities for neural network without CA125 on validation data.**

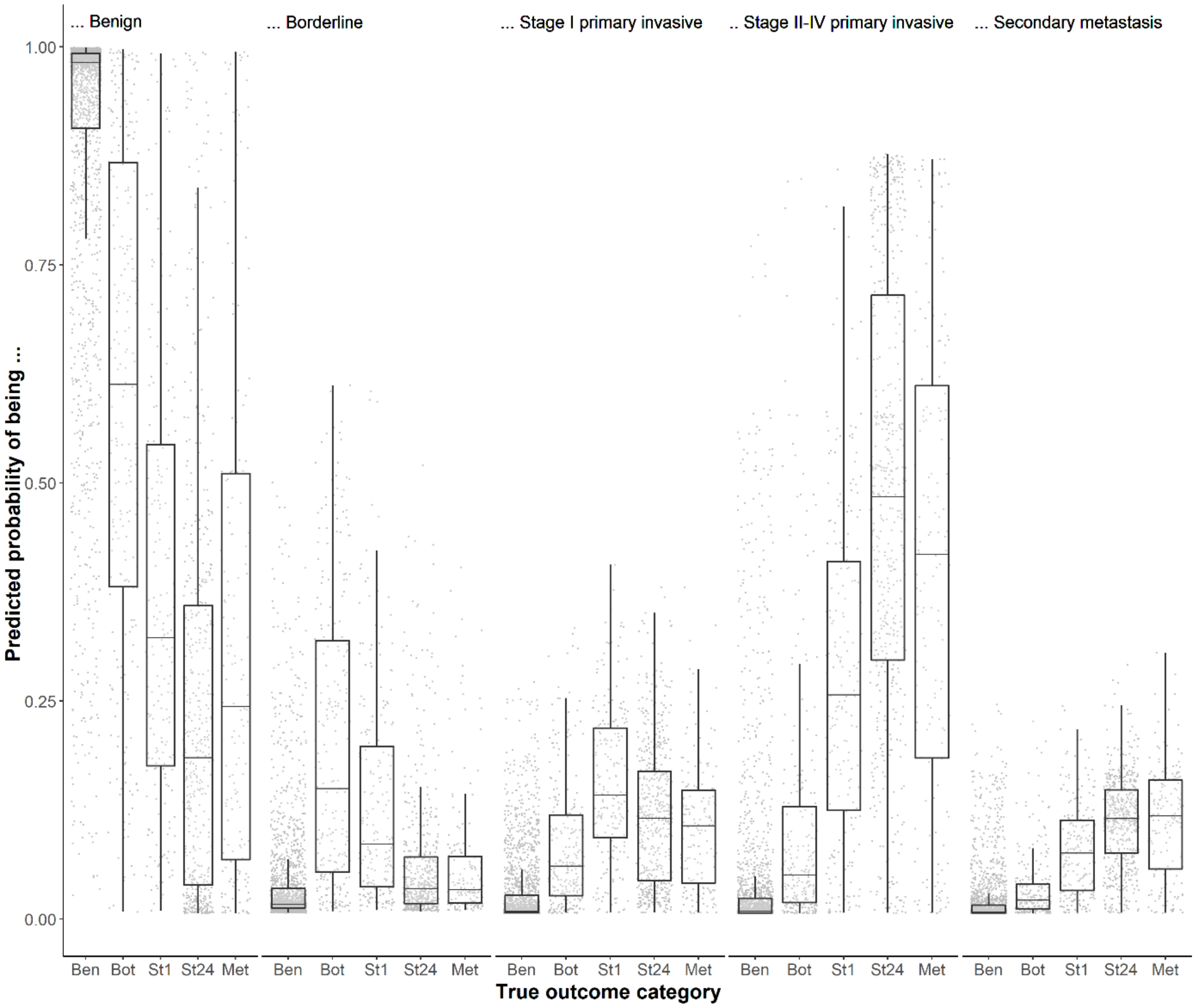

Abbreviations: Ben, benign; Bot, borderline; St1, stage I primary invasive; St24, stage II-IV primary invasive, Met, secondary metastatic.

**Figure S17. Box plots of estimated probabilities for support vector machine without CA125 on validation data.**

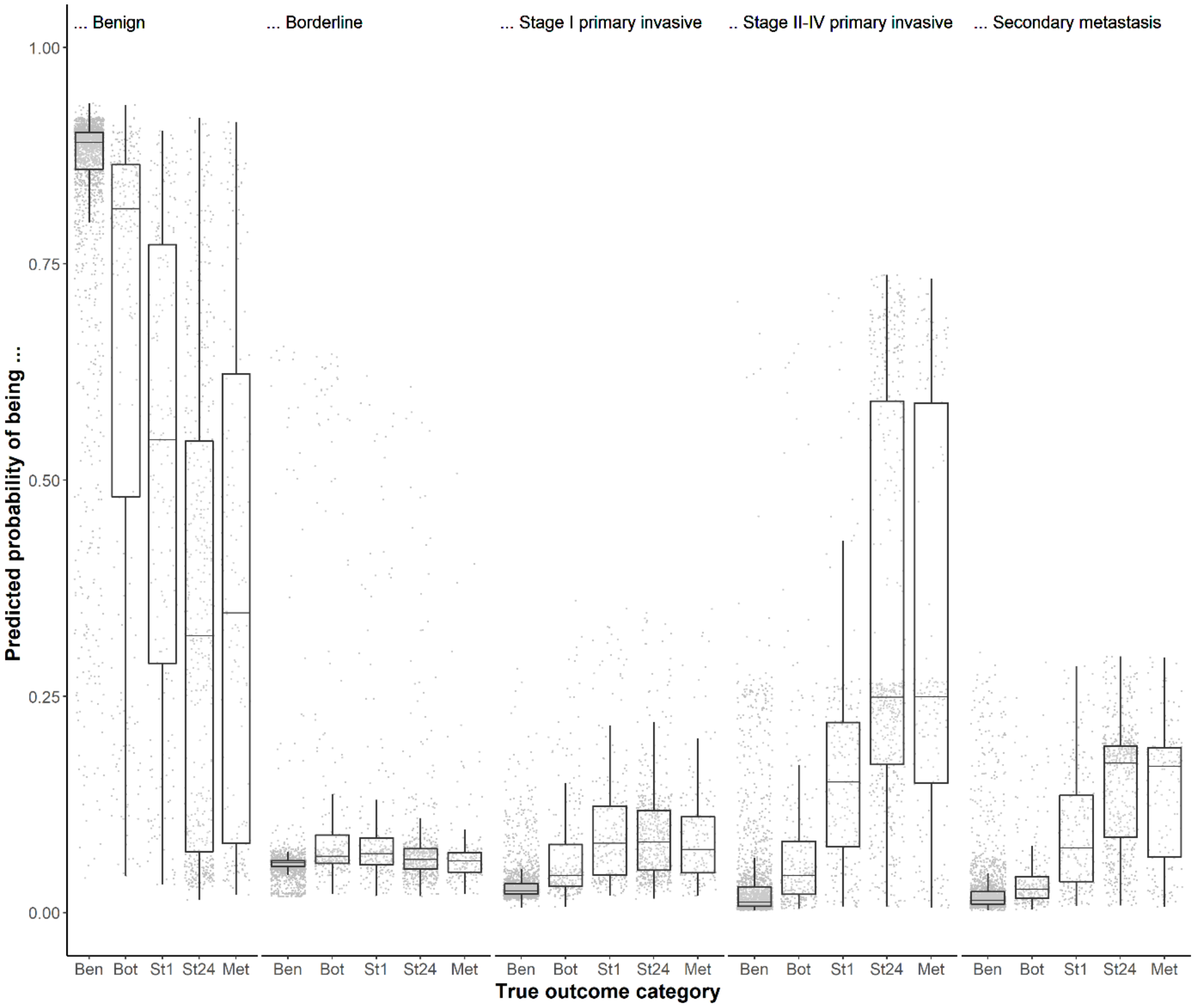

Abbreviations: Ben, benign; Bot, borderline; St1, stage I primary invasive; St24, stage II-IV primary invasive, Met, secondary metastatic.

**Figure S18. Decision curves for models with CA125 on external validation data.**

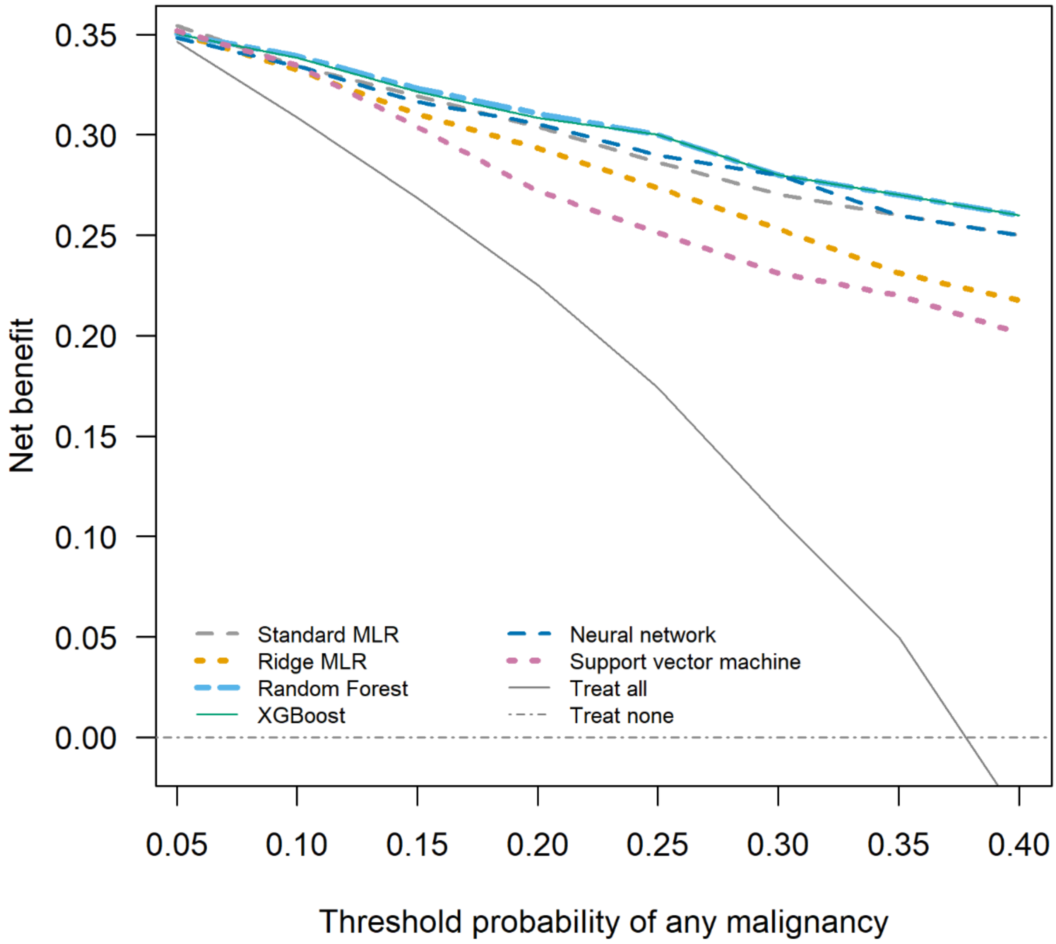

Abbreviations: MLR, multinomial logistic regression; XGBoost, extreme gradient boosting.

**Figure S19. Decision curves for models without CA125 on external validation data.**

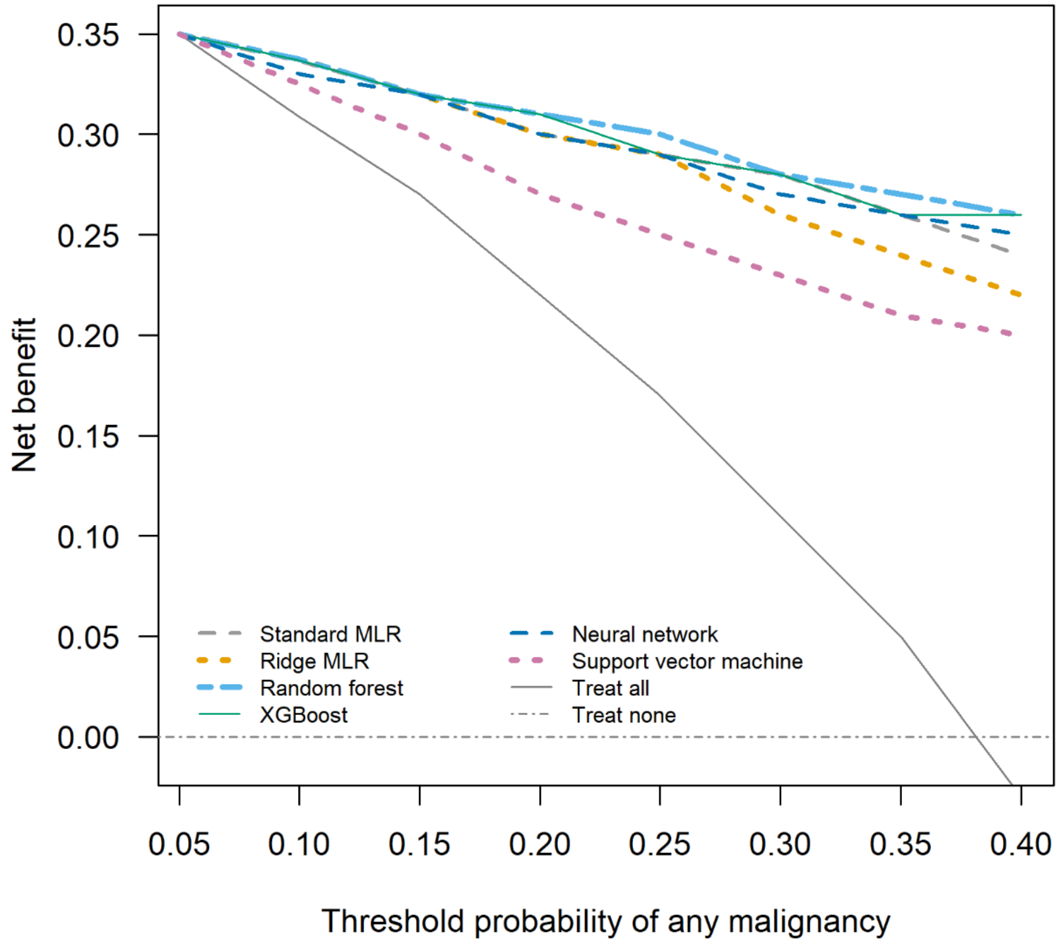

Abbreviations: MLR, multinomial logistic regression; XGBoost, extreme gradient boosting.

**Figure S20. Differences between the highest and lowest estimated probability for each outcome across the six models with CA125 (panel A) and the six models without CA125 (panel B) for patients in the external validation dataset. Each dot denotes the difference between the highest and the lowest estimated probability for one patient. This means that each patient is shown five times in each panel, once for each outcome category. For example, at the far left, the difference between the highest and lowest estimated probability for a benign tumor is shown for all 3199 patients in the dataset. The box represents the interquartile range which contains the middle 50% of the differences. The line inside the box indicates the median. Whiskers correspond to the 5th and 95th percentile.**

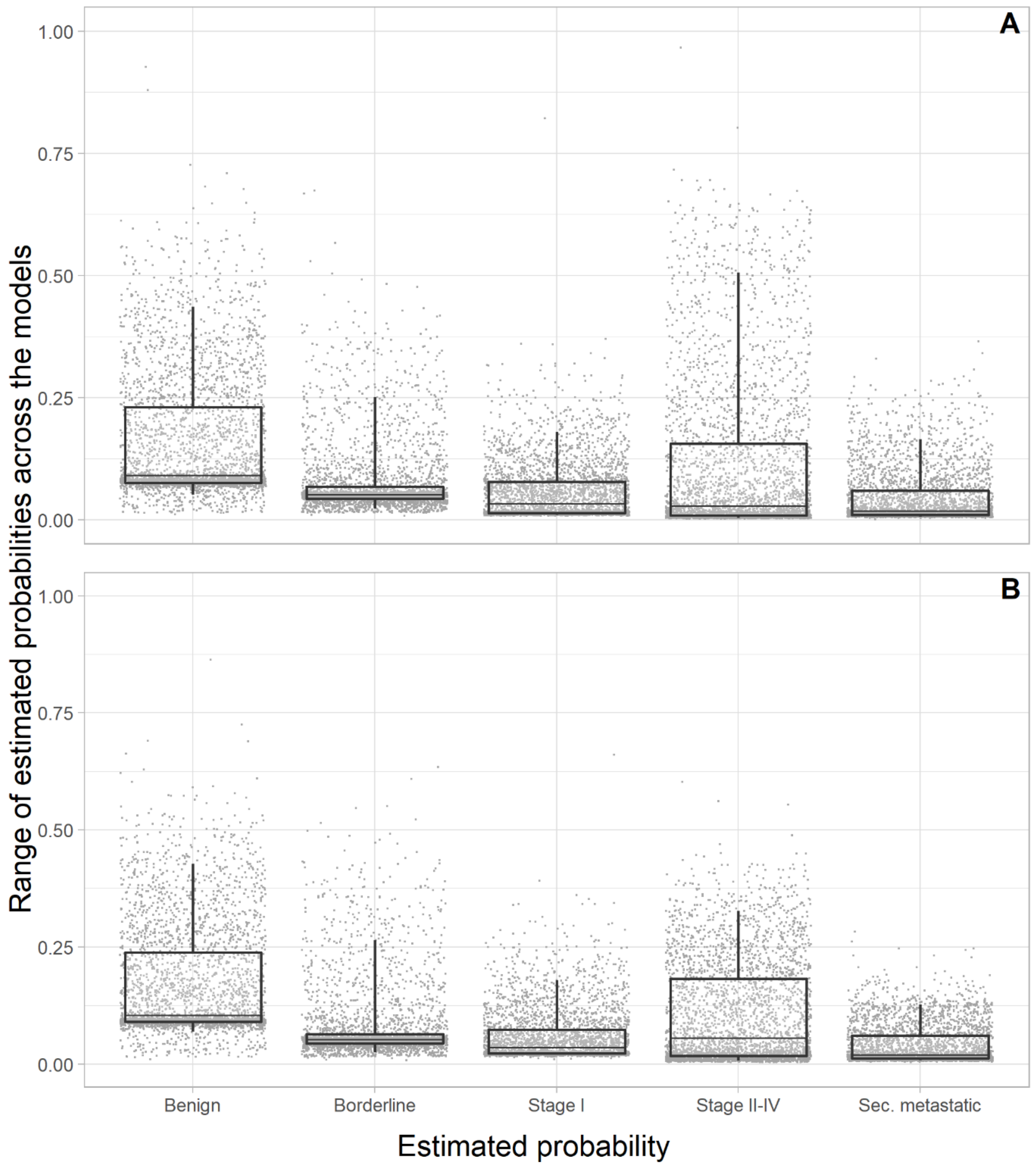

**Figure S21. Scatter plots of the estimated probability of a benign tumor for each pair of models without CA125 on validation data.**

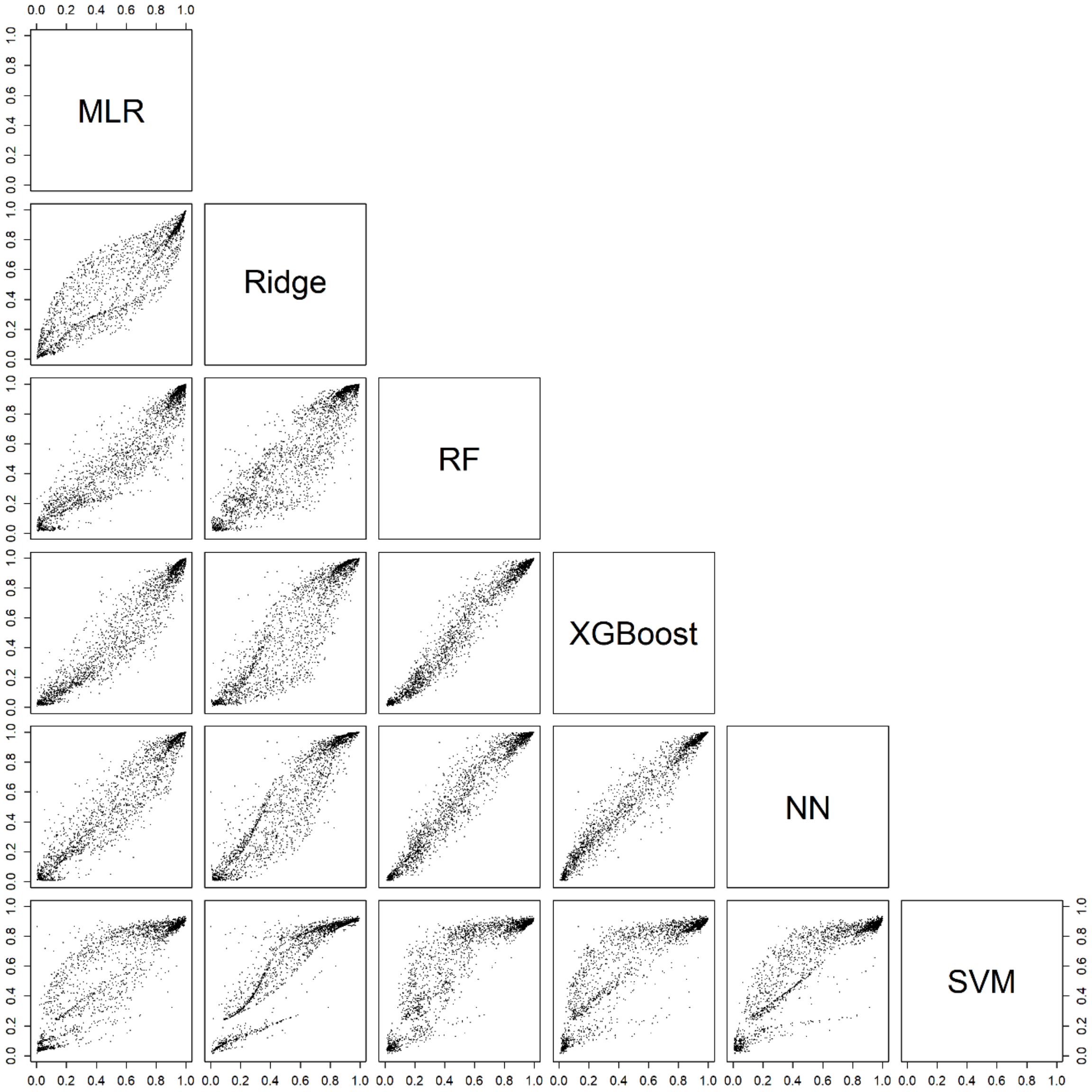

Abbreviations: MLR, multinomial logistic regression; RF, random forest; XGBoost, extreme gradient boosting; NN, neural network; SVM, support vector machine.

**Figure S22. Scatter plots of the estimated risk of a borderline tumor for each pair of models without CA125 on validation data.**

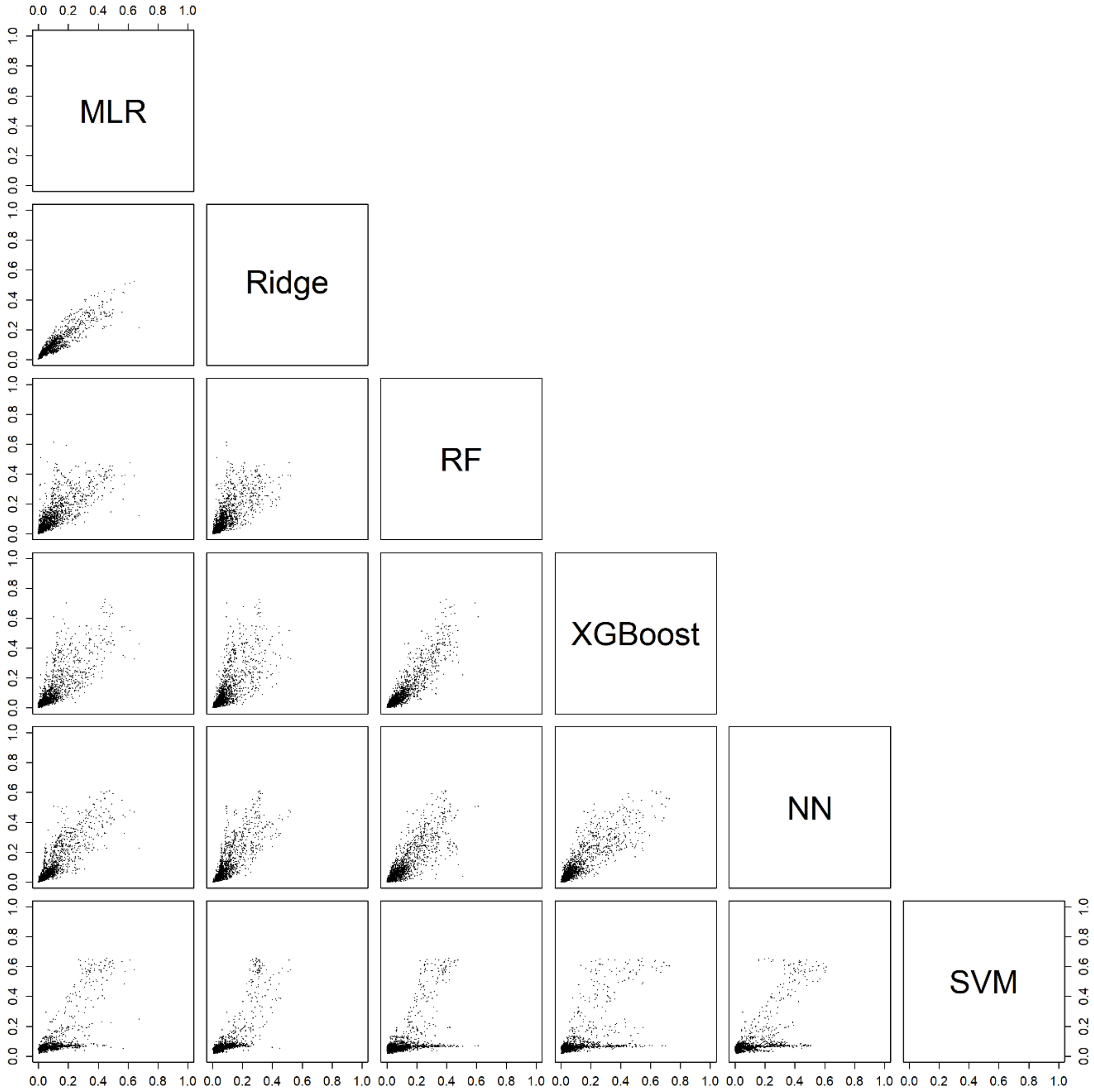

Abbreviations: MLR, multinomial logistic regression; RF, random forest; XGBoost, extreme gradient boosting; NN, neural network; SVM, support vector machine.

**Figure S23. Scatter plots of the estimated risk of a stage I primary invasive tumor for each pair of models without CA125 on validation data.**

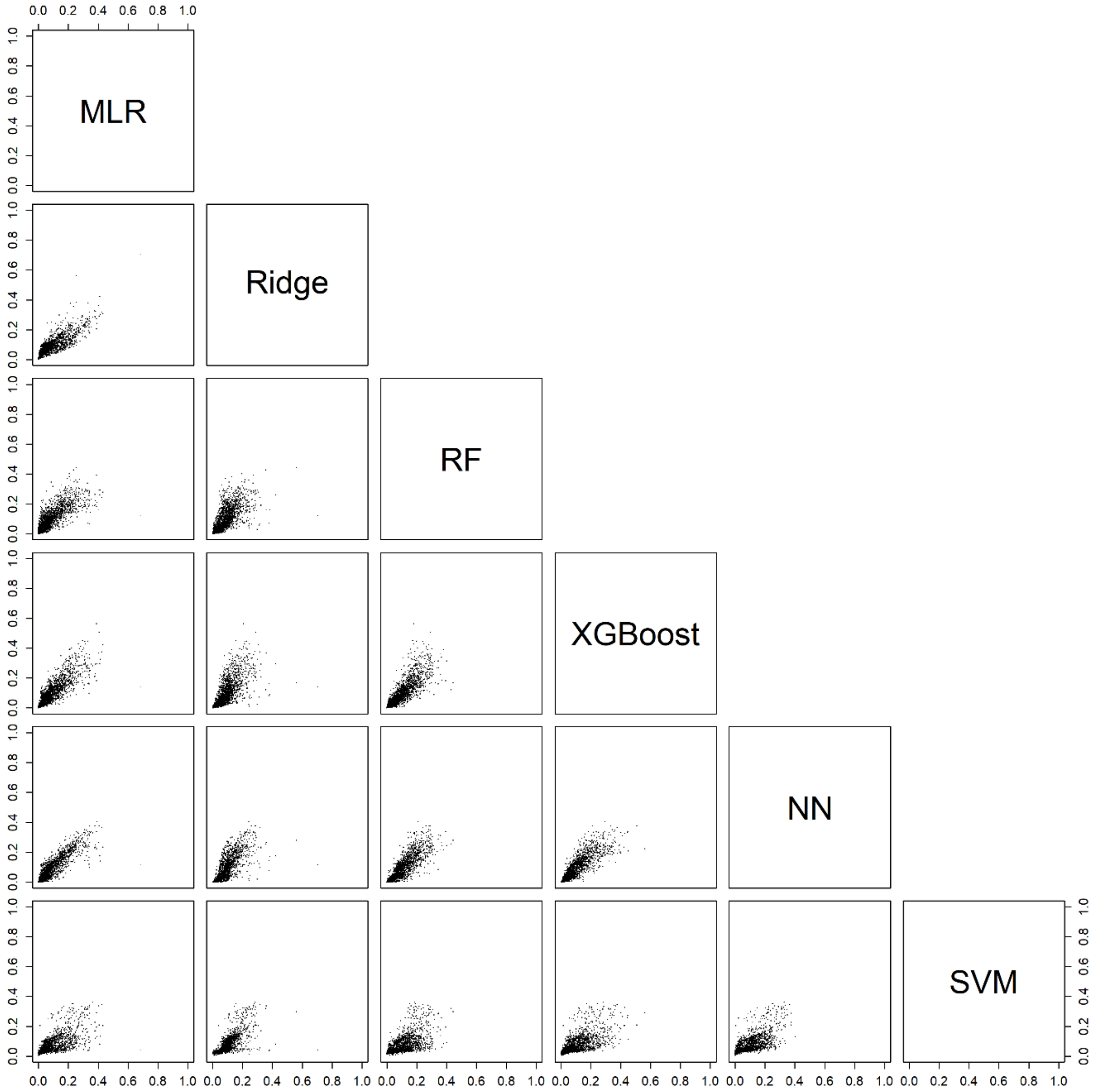

Abbreviations: MLR, multinomial logistic regression; RF, random forest; XGBoost, extreme gradient boosting; NN, neural network; SVM, support vector machine.

**Figure S24. Scatter plots of the estimated risk of a stage II-IV primary invasive tumor for each pair of models without CA125 on validation data.**

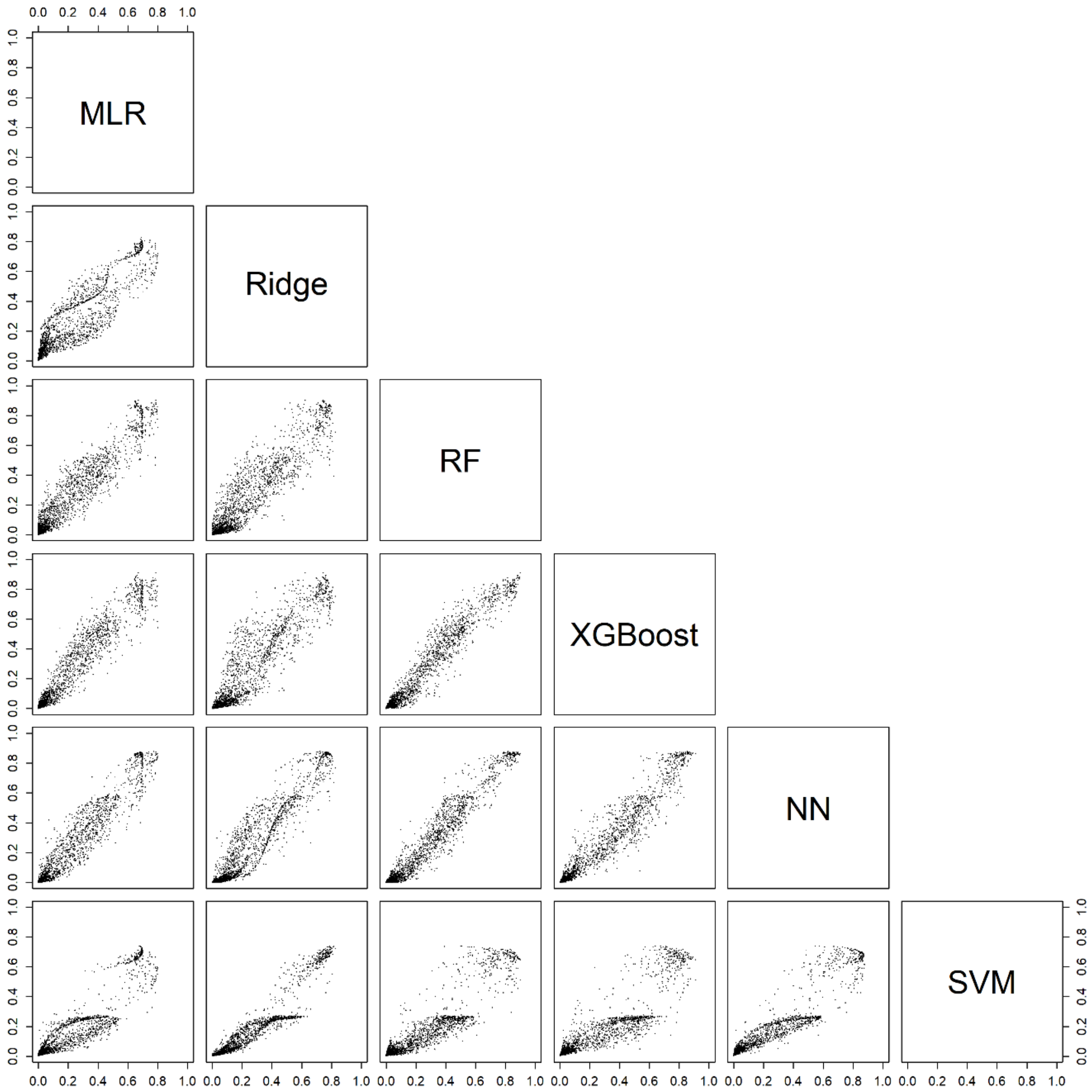

Abbreviations: MLR, multinomial logistic regression; RF, random forest; XGBoost, extreme gradient boosting; NN, neural network; SVM, support vector machine.

**Figure S25. Scatter plots of the estimated risk of a secondary metastatic tumor for each pair of models without CA125 on validation data.**

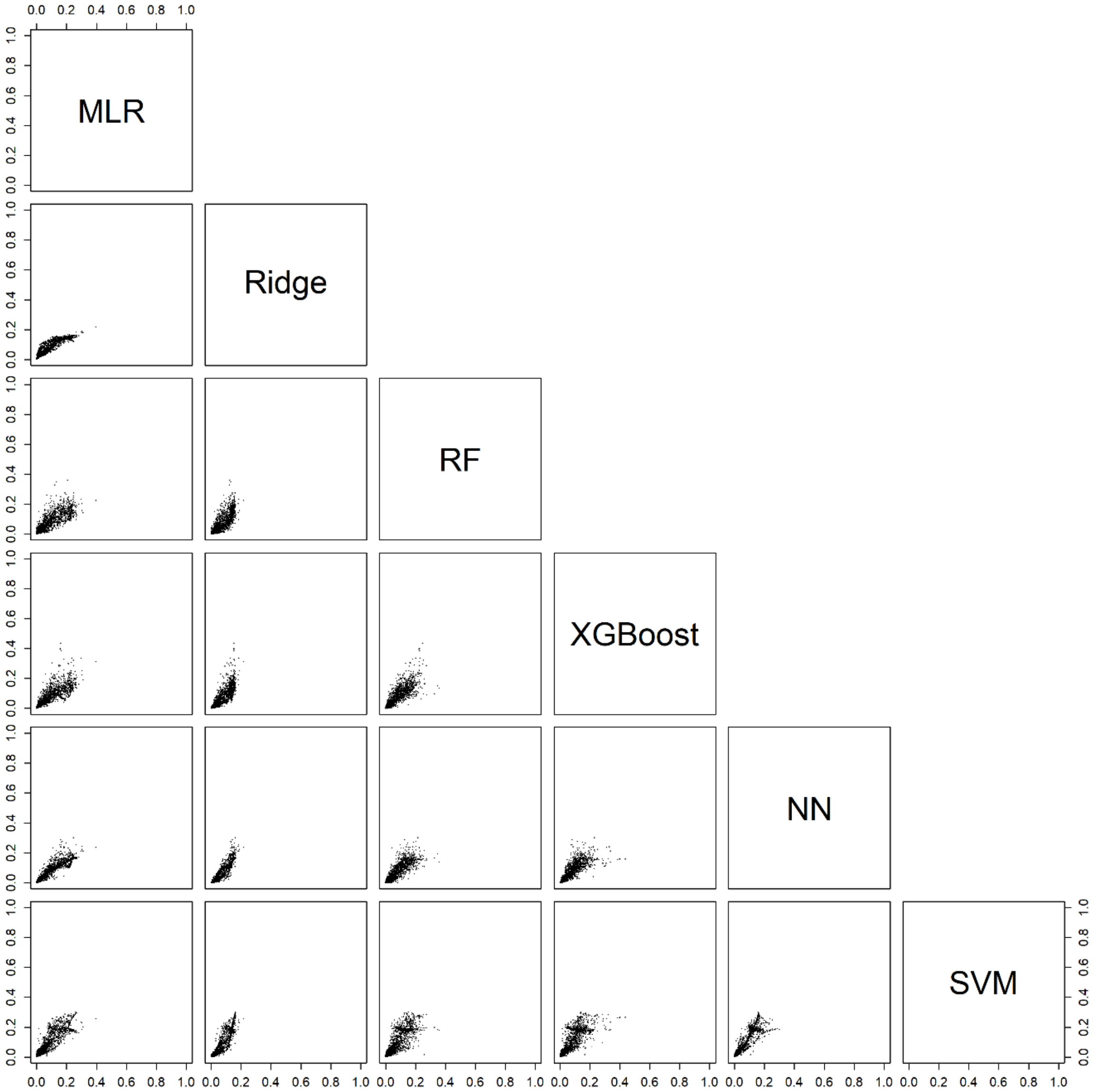

Abbreviations: MLR, multinomial logistic regression; RF, random forest; XGBoost, extreme gradient boosting; NN, neural network; SVM, support vector machine.
